# Sleep chart of biological aging clocks in middle and late life

**DOI:** 10.1101/2025.08.08.25333313

**Authors:** The MULTI Consortium, Cliodhna Kate O’Toole, Zhiyuan Song, Filippos Anagnostakis, Zhijian Yang, Ye Ella Tian, Michael R. Duggan, Chunrui Zou, Yue Leng, Yi Cai, Wenjia Bai, Cynthia H.Y. Fu, Michael S. Rafii, Paul Aisen, Gao Wang, Philip L. De Jager, Jian Zeng, Hamilton Se-Hwee Oh, Xia Zhou, Keenan A. Walker, Daniel W. Belsky, Andrew Zalesky, Eleanor M. Simonsick, Susan M. Resnick, Luigi Ferrucci, Christos Davatzikos, Junhao Wen

## Abstract

Optimal sleep plays a vital role in promoting healthy aging and enhancing longevity. This study proposes a Sleep Chart to assess the relationship between self-reported sleep duration and 23 biological aging clocks across 17 organ systems or tissues and 3 omics data types (imaging^1^, proteomics^2^, and metabolomics^3^). First, a systemic, U-shaped pattern shows that both short (<6 hours) and long (>8 hours) sleep duration are linked to elevated biological age gaps (BAGs) across 9 brain and body systems and 3 omics types. The lowest BAGs are achieved between 6.4 and 7.8 hours of sleep duration, and vary by organ and sex in the UK Biobank (ages 37–84 years). Furthermore, short and long sleep duration, compared to a normal sleep duration ([6-8] hours), are consistently linked to increased risk of systemic diseases beyond the brain and all-cause mortality, with evidence from genetic correlations and time to incident disease predictions, such as migraine, depression, and diabetes. Finally, short and long sleep duration are associated with late-life depression via distinct pathways: long sleep may contribute indirectly through biological aging processes, while short sleep shows a more direct link. Although our Mendelian randomization does not show strong causal effects from disease to sleep disturbances, it does not fully rule out the possibility that sleep disturbances may, in part, reflect underlying disease burden. Our findings suggest that the U-shaped relationship is likely driven by modifiable sleep disturbances rather than genetic predisposition, highlighting the potential of sleep optimization to support healthy aging, lower disease risk, and extend longevity. An interactive web portal is available to explore the Sleep Chart at: https://labs-laboratory.com/sleepchart.

## Main

Sleep is a fundamental biological process essential for physical restoration, cognitive functioning, and overall health^4^. Increasing evidence underscores its association with aging^5^, disease susceptibility^6^, and longevity^7^. Both insufficient and excessive sleep duration have been linked to a wide range of adverse health outcomes, including cardiometabolic disease^8^, cognitive decline^9^, and psychiatric disorders^10^, such as late-life depression (LLD^11^). Importantly, sleep is likely modifiable^12^, making it a potential target for promoting healthy aging and reducing the burden of age-related diseases across the lifespan.

In parallel, the field of aging research has seen rapid progress through the development of biological aging clocks derived from imaging and multi-omics data, like magnetic resonance imaging (MRIBAG^1,13^), plasma proteomics (ProtBAG^2,14^), and metabolomics (MetBAG^3^). These clocks aim to quantify the biological age of individuals across organ systems and molecular layers, enabling a more granular understanding of aging beyond chronological or calendar age. Organ-specific biological age gaps (BAGs) derived from these clocks have been used as intuitive and personalized biomarkers to quantify biological aging and have shown great predictive value for disease morbidity, cognition, and mortality risk^15,16^. This multi-organ, multi-omics aging clock framework offers a promising framework to model human health and disease in a systems-level and personalized manner.

Prior studies have demonstrated a nonlinear, U-shaped relationship between sleep duration and several phenotype-based aging clocks^17,18,19,20^, suggesting that both short (e.g., <6 h) and long (e.g., >8 h) sleep may accelerate neurobiological aging. However, it remains unclear whether this relationship generalizes beyond the brain to body systems (i.e., multi-organ^21,22,23,24^) and omics layers (i.e., multi-omics^2^), and is similar in males and females. Do similar U-shaped patterns emerge in the structural, functional, and molecular hallmarks of aging in organs and tissues beyond the brain? Are the observed U-shaped associations and empirically derived sample minimum values of the BAG-sleep relationships consistent across different sexes and organ systems? To address these gaps, we leveraged large-scale population biobanks, combining both individual-level and summary-level data, consolidated through the MULTI Consortium^21,3^ (**Method 1** and **Supplementary Table 1**), to comprehensively map how sleep duration associates with biological aging across multiple systems and to elucidate the pathways through which disrupted sleep may drive age-related diseases and mortality. Our analysis focused on the UK Biobank’s (UKBB) questionnaire-derived sleep duration (Field ID: 1160). Self-reported measures are less objective than actigraphy or polysomnography, capturing different yet complementary aspects of sleep biology, with only moderate correlations between modalities^25,26^. The large sample size (∼500,000) allows robust identification of nonlinear associations using generalized additive models (**GAM; Method 2**).

This study addresses several scientific and clinical questions. First, we examined the nonlinear, U-shaped associations between sleep duration and 23 multi-organ BAGs derived from multi-omics data (**Fig. 1**). We reinforced this pattern using *in vivo* imaging markers (**Extended Data Figure 1**) and circulating molecular phenotypes, including plasma proteomics and metabolomics **(Extended Data Figures 2-3**). Secondly, we assessed whether abnormal sleep duration patterns (i.e., short or long sleep duration) were adversely associated with all-cause mortality and systemic disease endpoints (DE) beyond the brain (**Fig. 2–3**). Thirdly, we examined whether short and long sleep duration are differentially associated with two distinct subtypes of LLD^11^ through separate mediational pathways (**Fig. 4**). Finally, we explored whether sleep disturbances act as modifiable risk factors for disease (or genetic predisposition), consequences of disease burden, or reflect a potential bidirectional relationship (**Fig. 4**, **Extended Data Figure 4** and **Supplementary Note 4**). All results, code, and summary statistics are publicly available at the Sleep Chart portal: https://labs-laboratory.com/sleepchart.

**Figure 1:**
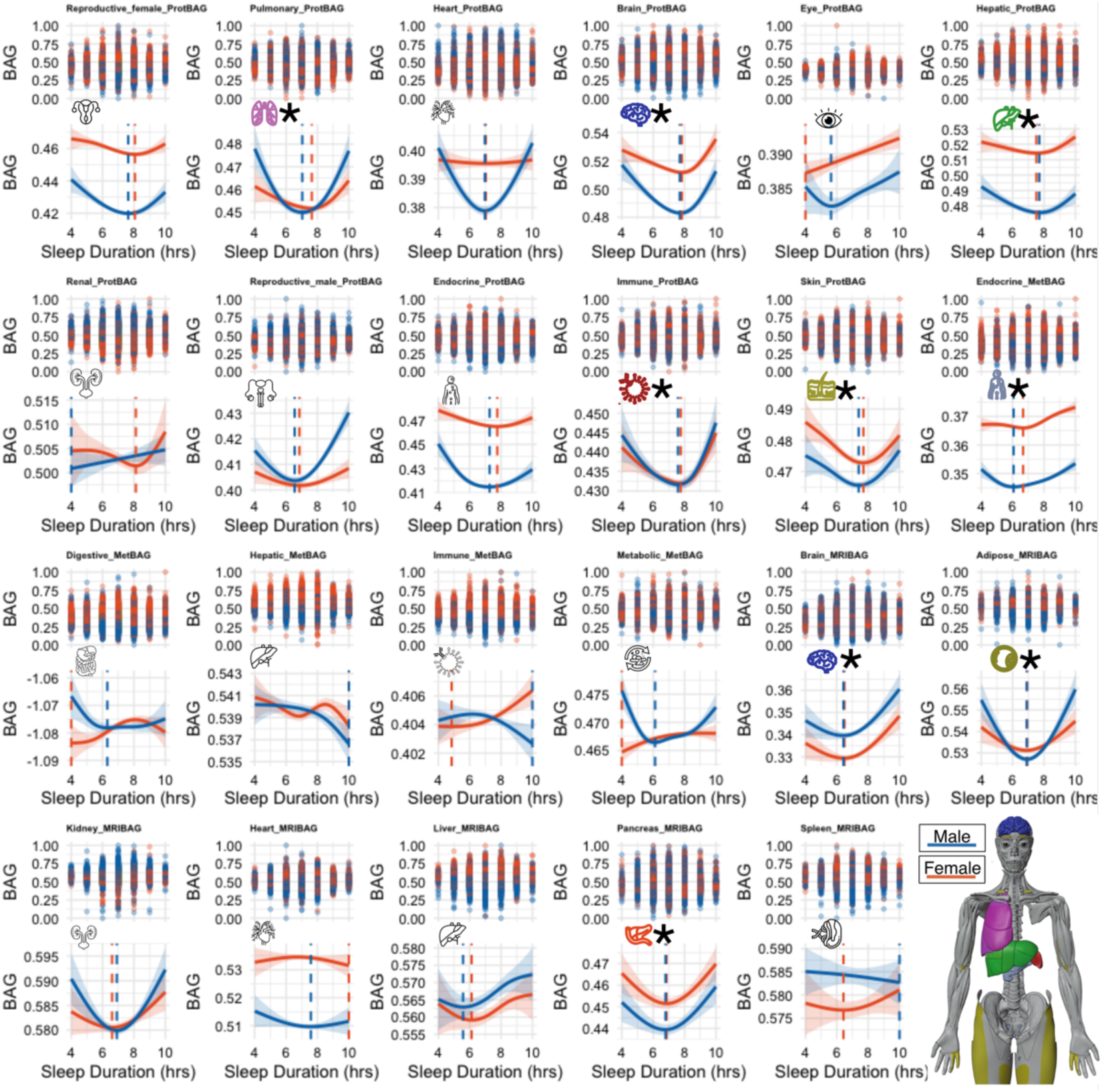
Sleep Chart delineates U-shaped patterns between sleep duration and biological aging clocks. Sleep duration (x-axis) exhibits nonlinear U-shaped relationships with 9 out of 23 BAGs across 17 organs and 3 omics types (y-axis; units are normalized years): 11 ProtBAGs for plasma proteomics, 5 MetBAGs for plasma metabolomics, and 7 MRIBAGs for *in vivo* MRI data. For each BAG, we fitted a generalized additive model (GAM) with cubic regression splines to assess the nonlinear association between sleep duration and BAG, stratified by sex and sex-sleep interaction term. The solid curves depict estimated BAG, while shaded bands represent the 95% confidence interval (CI). Significant signals (two-sided P-value<0.05/23) are shown with colored icons and the * symbol. Sample minimum values (in hours) of sleep duration, minimizing the BAG load, are displayed separately for males and females. Sample sizes of each sleep-BAG association are presented in **Supplementary Table 2**.

**Figure 2:**
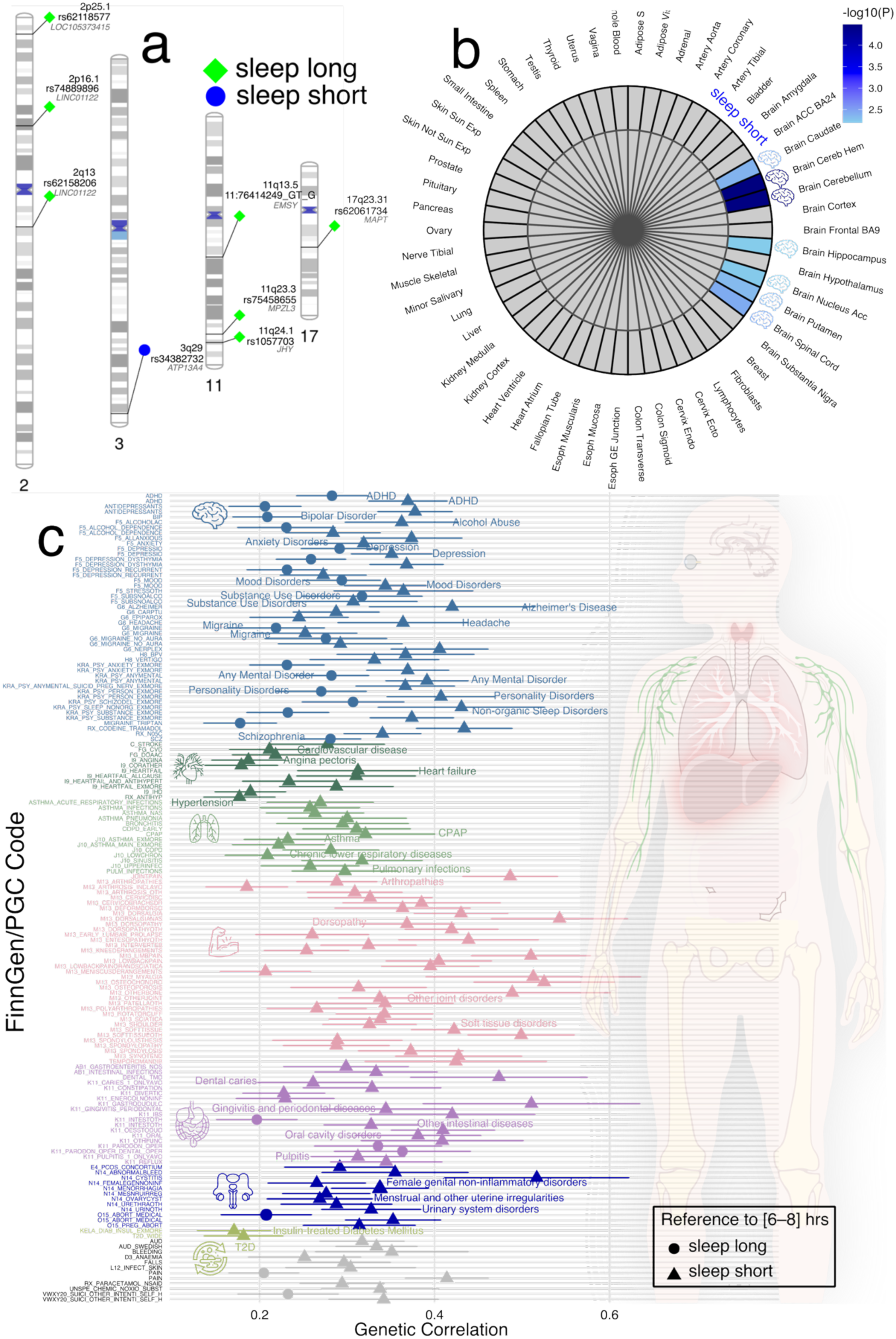
Genetic evidence for a systemic landscape of abnormal sleep duration patterns compared to normal sleep duration. **a**) Top lead SNP, nearest gene, and cytogenetic regions associated with short ([4–6) hours hours) and long sleep duration ((8–10] hours) compared to a normal sleep duration ([6-8] hours), with significant genomic loci identified using the genome-wide significance threshold (two-sided P-value<5×10^−8^). **b**) MAGMA gene property analysis of tissue-specific expression across 54 GTEx v8 tissues. Statistical significance was determined using FDR-adjusted P-values (<0.05). **c**) Genetic correlation estimates via LDSC between the two abnormal sleep duration patterns and 527 disease endpoints from FinnGen and Psychiatric Genomics Consortium (PGC) (two-sided P-value<0.05/527). An online interactive webpage is available at https://labs-laboratory.com/sleepchart/sleep_gc.html to facilitate visualization. Sample sizes of the FinnGen and PGC DEs are presented in **Supplementary Table 7**.

**Figure 3:**
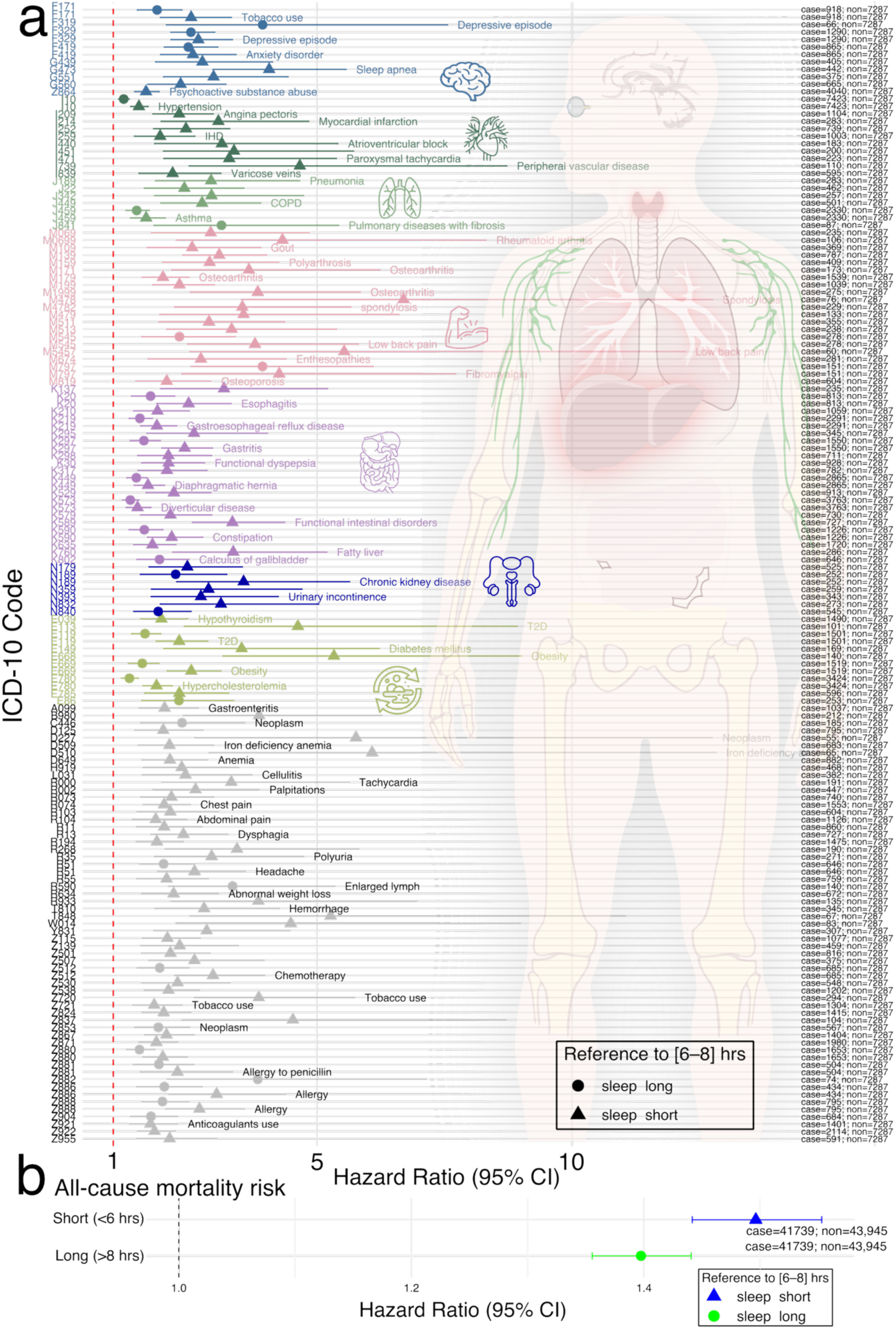
Clinical evidence for a systemic involvement of abnormal sleep duration patterns compared to normal sleep duration. **a**) Using normal sleep duration, [6-8] hours, as the reference, we treated sleep duration patterns as categorical variables ([4–6) hours for short sleep, and (8–10] hours for long sleep) to estimate their association with 726 comorbidity-free disease endpoints defined by ICD-10 codes (limited to diseases with more than 50 cases). After applying Bonferroni correction for multiple comparisons (two-sided P-value< 0.05/726), we highlight the significant associations and annotate representative systemic diseases across various organ systems. **b**) Short and long sleep duration are associated with a higher risk of all-cause mortality. An online interactive webpage is available at https://labs-laboratory.com/sleepchart/sleep_cox.html to facilitate visualization.

**Figure 4:**
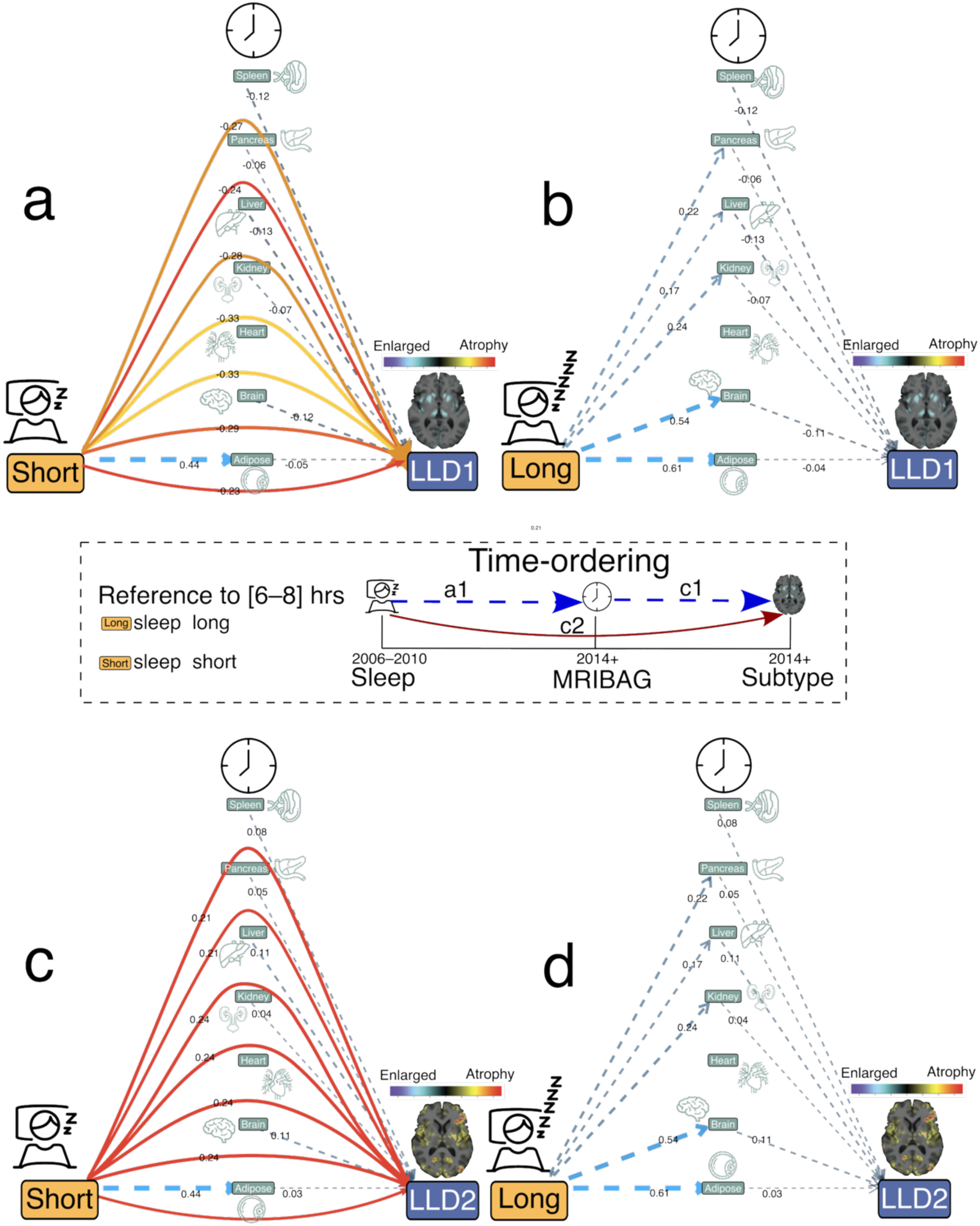
7 MRIBAGs mediate the effects of disturbed sleep duration patterns on 2 late-life depression subtypes. We tested whether 7 MRIBAGs mediate the effect of abnormal sleep duration on two AI-derived subtypes of late-life depression (LLD1 and LLD2^11^, using only the first visit of MRI). Participants were grouped into short (4-6 hours), normal ([6–8] hours; as reference), and long (8-10 hours) sleep categories: sleep duration→ MRIBAG→LLD1/2. For each LLD subtype and each contrast (short vs. normal, long vs. normal), we fit structural equation models (SEMs) using the *lavaan* package in R. The model included a direct path from the binary-coded sleep category to the LLD subtype (depicted as solid lines with warm colors) and indirect paths mediated through the MRIBAG (shown as dotted lines with cool colors). Only statistically significant pathways (two-sided P-value<0.05/7) and estimated path coefficients are displayed. All models were adjusted for relevant covariates, including age, sex, body mass index, and systolic and diastolic blood pressure, among others. Standardized estimates were obtained using 1,000 nonparametric bootstrap resamples. **a**) Mediational models for short sleep duration→MRIBAG→LLD1. **b**) Mediational models for long sleep duration→MRIBAG→LLD1. **c**) Mediational models for short sleep duration→MRIBAG→LLD2. **d**) Mediational models for long sleep duration→MRIBAG→LLD2. Our primary analyses were based on the temporal ordering of data collection, with sleep duration assessed at baseline and both MRIBAG and LLD subtypes measured at the second visit. Sample sizes of the analyses are presented in **Supplementary Table 8**.

## Results

### The U-shaped relationship between sleep duration and 23 multi-organ, multi-omics biological aging clocks

Using GAMs with the UKBB data, we assessed the nonlinear, U-shaped relationship between sleep duration and 23 multi-organ, multi-omics BAGs (P<0.05/23; **Fig. 1**). We quantified nonlinearity in the sleep–BAG relationship using the effective degrees of freedom (EDF) of the smooth term, a measure of curve complexity (**Method 2-3**).

We confirmed that 9 out of the 23 BAGs exhibited statistically significant nonlinear associations (P<0.05/23). Among the 9 ProtBAGs, the brain ProtBAG showed the strongest U-shaped association with sleep duration (EDF=3.61; P_1_<1×10^−20^). At the population mean level, females exhibited significantly higher brain ProtBAG values than males (P_2_<1×10^−20^). The interaction between sex and sleep duration approached nominal significance (P_3_=0.06), indicating a potential trend toward sex-specific associations. We then estimated the sample minimum of the BAG-sleep curves based on the peak of the smoothed spline curves for the brain ProtBAG, identifying 7.82 hours for females and 7.70 hours for males. Similar U-shaped relationships were also evident for the pulmonary (EDF=3.21; P_1_=1.09×10^−4^), hepatic (EDF=3.13; P_1_=1.36×10^−3^), immune (EDF=3.47; P_1_=1.78×10^−5^), and skin (EDF=3.21; P_1_=3.12×10^−6^) ProtBAGs. Among the 5 MetBAGs, the endocrine MetBAG showed a significant U-shaped relationship (EDF=1.04; P_1_=3.97×10^−5^), and the mean population value was different between females and males (P_2_<1×10^−20^; P_3_=5.72×10^−7^), with the estimated sample minimum values being 6.67 hours for females and 6.06 hours for males. Finally, among the 7 MRIBAGs, the brain MRIBAG showed the most significant U-shaped relationship (EDF=1.94; P_1_=3.85×10^−7^), and the mean population value was different between females and males (P_2_<1×10^−20^; P_3_=0.84). The optimal sleep time was 6.48 hours for females and 6.42 hours for males. Significant signals were also observed for the adipose (EDF=1.93; P_1_=2.84×10^−4^) and pancreas (EDF=1.95; P_1_=9.95×10^−6^) MRIBAGs. Detailed statistics, including the P value, sample size, chosen family distribution, and EDF, are presented in **Supplementary Table 2** and **Supplementary File 1.**

Several sensitivity analyses were performed. **Supplementary Figure 1** displays Quantile-Quantile (QQ) plots and residual vs. fitted (RVF) plots for the best model of each BAG. **Supplementary Figure 2-16** and **Supplementary Table 3** present a full set of analyses to scrutinize the robustness of the U-shaped pattern, including a potential replication of the U-shaped relationship in two independent datasets^27,28^, though the sample sizes (*N*=385 and 573) are much smaller and the populations are much older than UKBB. We also observed this U-shaped pattern between sleep duration and 720 organ-specific imaging-derived phenotypes (IDP; **Extended Data Figure 1**), 342 organ-enriched plasma proteins (**Extended Data Figure 2** and **Supplementary Figure 17**), and 107 organ-associated plasma metabolites (**Extended Data Figure 3**, **Method 4-6**, and **Supplementary File 2-4**). A detailed discussion of these results and their implications in sleep-related hypotheses is presented in **Supplementary Note 1**.

Overall, across the nine significant proteomic, metabolomic, and MRI-based BAGs, sample minimum values for sleep duration ranged from 6.5 to 7.8 hours for females and 6.4 to 7.7 hours for males. Based on this, we defined normal sleep duration as 6 to 8 hours [6-8] for subsequent genetic and time to incident disease predictive analyses. Although our definition is data-driven by the sample minimum values range observed, it is important to note that the definition of normal sleep varies between studies and is often associated with cultural and environmental factors^29^.

To summarize, we observed robust U-shaped associations between sleep duration and multi-organ BAGs, extending beyond the brain to organ systems such as the pulmonary, hepatic, and immune systems. This U-shaped pattern was subsequently confirmed in *in vivo* IDPs, plasma proteins, and metabolites. Given strong evidence of the U-shaped relationship across widespread biomarkers at both phenotypic and molecular levels, we classified sleep duration into short (<6 hours), long (>8 hours), and normal ([6–8] hours) categories for all downstream analyses.

### Long and short sleep duration are associated with systemic diseases beyond the brain and all-cause mortality

In UKBB, our genome-wide association study (GWAS; **Method 7**) identified 8 genetic loci (P<5×10^−8^) associated with abnormal sleep duration patterns (**Fig. 2a**). A locus at 3q29 (top lead SNP: rs34382732) was associated with short sleep duration, whereas long sleep duration was linked to multiple loci, highlighting potential genetic heterogeneity between short and long sleep patterns. We denoted the genomic loci using their top lead SNPs defined by FUMA (**Supplementary Note 2**), considering linkage disequilibrium; the genomic loci are presented in **Supplementary Table 4**. The Manhattan plot and QQ plot, as well as the LDSC^30^ intercept, are presented in **Supplementary Figure 18**. We then conducted gene-property analyses^31^ to assess tissue-specific expression patterns of genetic signals associated with short and long sleep duration, using gene expression data from 54 human tissues provided by the Genotype-Tissue Expression Program (GTEx) v8^32^. Notably, only the genetic architecture of short sleep duration demonstrated significant enrichment (P_FDR_<0.05) in several brain regions, including the cerebellum, caudate, and hippocampus (**Fig. 2b**), but not long sleep duration.

Given the relatively smaller number of genomic loci in our GWASs, despite the large sample size (N > 300,000), we hypothesize that the U-shaped association between sleep duration and BAG is primarily environmentally driven and modifiable. As such, we hypothesized that this U-shaped pattern observed at the BAG level would be flattened and attenuated when examining BAG polygenic risk scores (PRS) derived in our previous studies^1,13,2,3^. Our results supported our hypothesis (**Extended Data Figure 4**). Notably, the endocrine MetBAG showed a J-shaped association, with higher genetic risk in individuals reporting shorter sleep duration, while the metabolic MetBAG demonstrated a linear inverse relationship, indicating lower PRS with longer sleep. Additionally, genetic correlation analyses revealed only a significant positive correlation between short sleep duration and endocrine MetBAG, and between long sleep duration and pancreas MRIBAG (P < 0.05/23) (**Supplementary Table 5**). These findings imply that while most sleep-BAG relationships are not genetically mediated, certain organ systems, particularly endocrine and metabolic pathways, may share partial genetic underpinnings with habitual sleep duration.

Using summary-level data from our GWAS and FinnGen and PGC, genetic correlation analyses revealed 153 positive associations between abnormal sleep duration patterns and systemic diseases across multiple organ systems (P<0.05/527), dominated by short sleep duration (**Fig. 2c**). For short sleep duration, we observed robust correlations with cardiovascular diseases, including ischemic heart disease (*g_c_*=0.19±0.04; P=2.52×10^−6^), heart failure (*g_c_*=0.31±0.06; P=5.80×10^−6^), and coronary atherosclerosis (*g_c_*=0.18±0.04; P=1.51×10^−5^), metabolic disorders, such as type 2 diabetes (*g_c_*=0.18±0.04; P=5.33×10^−5^) and insulin-requiring diabetes (*g_c_*=0.17±0.04; P=4.45×10^−5^), and musculoskeletal conditions, like low back pain (*g_c_*=0.40±0.06; P=1.13×10^−12^), osteoarthritis (*g_c_*=0.19±0.05; P=7.07×10^−5^) and soft tissue disorders (*g_c_*=0.50±0.06; P=2.04×10^−16^). Neurological and psychiatric associations included migraine (*g_c_*=0.25±0.06; P=1.91×10^−5^), depression (*g_c_*=0.37±0.04; P=1.57×10^−18^), anxiety (*g_c_*=0.32±0.06; P=1.83×10^−7^), substance use disorders (*g_c_*=0.37±0.05; P=3.73×10^−15^), and suicidality (*g_c_*=0.34±0.06; P=4.28×10^−8^), indicating broad involvement of the central nervous system. Pulmonary and infectious conditions, such as asthma (*g_c_*=0.22±0.05; P=2.50×10^−5^), bronchitis (*g_c_*=0.42±0.12; P=5.00×10^−4^), and chronic obstructive pulmonary disease (*g_c_*=0.28±0.05; P=2.74×10^−7^), were also genetically correlated, as were gastrointestinal and hepatic diseases, including reflux (*g_c_*=0.34±0.06; P=6.09×10^−8^), diverticulosis (*g_c_*=0.23±0.04; P=1.14×10^−6^), and irritable bowel syndrome (*g_c_*=0.32±0.09; P=3.00×10^−4^). In contrast, long sleep duration showed a more focused genetic correlation profile, predominantly involving brain-related phenotypes, such as major depressive disorder (*g_c_*=0.29±0.04; P=2.57×10^−11^), schizophrenia (*g_c_*=0.28±0.03; P=3.47×10^−16^), bipolar disorder (*g_c_*=0.21±0.03; P=1.09×10^−7^), alcohol dependence (*g_c_*=0.23±0.05; P=3.26×10^−5^), ADHD (*g_c_*=0.28±0.04; P=2.24×10^−12^), and migraine (*g_c_*=0.28±0.07; P=7.09×10^−5^), suggesting potential compensatory or indirect neuropsychiatric mechanisms (further elucidated in **Fig. 4**). **Supplementary File 5a** presents detailed statistics for our genetic correlation analysis.

Taken together, these findings support that short and long sleep duration have distinct genetic architectures from each other (**Fig. 2a**). While both show genetic correlations with various systemic diseases, short sleep duration exhibits broader systemic associations, whereas long sleep duration is more specifically linked to brain-related traits (**Fig. 2c**). **Supplementary Note 3**, **Supplementary Figure 19-21**, and **Supplementary File 5b-c** present a discussion comparing our GWAS and genetic relevance with previous studies^33,34,35,36,37^.

Leveraging ICD code-based clinical diagnosis and echoing the genetic correlation results, we further assessed the relationships between abnormal sleep duration patterns and the time to incident disease prediction, as well as all-cause mortality, using a survival analysis framework in UKBB (**Method 8**). We identified 153 significant associations between sleep duration patterns (both short and long) and DEs (P<0.05/726), after correcting for multiple comparisons across endpoints with at least 50 cases (**Fig. 3a**), with a notable predominance of associations linked to short sleep duration (1.20<hazard ratio<6.69). These findings span multiple organ systems, supporting the systemic and direct association with disturbed sleep. Within the brain-related disorders, short sleep was significantly associated with depressive episodes (F329), anxiety disorders (ICD code: F419), and primary insomnia (G473), reinforcing the well-established link between sleep and mental health. In the metabolic domain, we observed associations with obesity (E669), type 2 diabetes (E119), and hyperlipidemia (E780), aligning with prior evidence that short sleep disrupts metabolic homeostasis. Cardiovascular outcomes such as essential hypertension (I10), ischemic heart disease (I209, I252), and arrhythmias (I471, I440) were also enriched, suggesting increased cardiovascular risk among individuals with short sleep. Pulmonary conditions, including chronic obstructive pulmonary disease (J449) and asthma (J459), were linked to both short and long sleep, though more strongly with short sleep. Finally, a cluster of digestive disorders, including gastritis and duodenitis (K297), gastroesophageal reflux disease (K219), and functional intestinal disorders (K590), was significantly predicted by both long and short sleep duration. These findings underscore the broad biological associations of both insufficient and prolonged sleep, highlighting distinct mechanistic pathways by which short and long sleep duration relate to systemic DEs (further elucidated in **Fig. 4**). We also performed a sensitivity analysis for time to incident disease prediction using [7-9] hours as normal sleep duration (**Supplementary Figure 22**), as suggested by the National Sleep Foundation^38^ for young/older adults.

In predicting all-cause mortality, we found that both short (hazard ratio=1.50 [1.44, 1.55]; P<1×10^−20^) and long sleep duration (hazard ratio=1.40 [1.36, 1.44]; P<1×10^−20^) patterns were positively associated with increased risk of death from any cause (**Fig. 3b**). **Supplementary File 6** presents detailed statistics for all survival analyses. **Supplementary Note 4**, **Supplementary File 7**, and **Extended Data Figure 5** present results of examining the associations between two sleep disorders (insomnia and hypersomnia) and the DEs, using the TriNetX dataset, identified in **Fig. 3a**, as well as all-cause mortality.

### Distinct mediating pathways from short or long sleep duration to disease endpoints in late-life depression via biological aging burden

Given that short sleep duration showed more significant associations with systemic DEs in our genetic correlation (**Fig. 3a**) and survival analysis (**Fig. 3b**) than long sleep duration, we hypothesize that long sleep may be linked to disease risk through more complex or indirect pathways, potentially serving as a marker of underlying health conditions or reflecting mediational, compensatory physiological responses rather than acting as a direct risk factor (**Method 9**).

To evaluate this hypothesis in UKBB, we performed structural equation modeling (SEM) using sleep duration patterns (short or long) as exposures, the 7 MRIBAGs as mediators, and two MRI-derived subtypes of late-life depression (LLD1 and LLD2^11^) as outcomes. We selected LLD as the outcome because it is closely linked to sleep disturbances (**Fig. 2-3**) and aging, making it a compelling model to explore potential mediating pathways. In the pathway linking short sleep to LLD1, characterized by preserved subcortical brain volume, we observed strong direct effects (c2 ranging from −0.33 to −0.24) across 6 organ systems; refer to **Method 9** for the definition of c1, c2, and a1 coefficients. Among the mediators, only the adipose MRIBAG showed a significant indirect effect (a1=0.44±0.13; c1=-0.05±0.005), suggesting a specific role for adipose aging in mediating this relationship. In contrast, for long sleep, the associations with LLD1 were predominantly mediated through organ-specific MRIBAGs, particularly the brain (a1=0.54±0.08; c1=-0.11±0.002) and adipose (a1=0.61±0.09; c1=-0.04±0.005) pathways. Notably, brain MRIBAG alone accounted for 62% of the total effect via mediation, followed by the adipose and liver MRIBAGs (24%), highlighting a more indirect and organ-mediated link between long sleep and late-life depression. Similar patterns were observed for LLD2, which were characterized by diffuse patterns of cortical atrophy in LLD (**Fig. 4**). All statistics for the mediation analyses are presented in **Supplementary File 8**.

Our mediation analysis treated sleep duration as a modifiable risk factor, based on the temporal sequence of data collection: sleep data were recorded at baseline, while brain MRI occurred during follow-up. As detailed in **Supplementary Table 6**, we conducted sensitivity analyses by alternately specifying brain MRIBAG and LLD1/2 as mediators and outcomes. However, due to the limitations of the data’s time-ordering, we cannot fully exclude the possibility that sleep disturbances result from underlying disease burden. To test this, we conducted Mendelian randomization analyses using 5 different estimators (**Supplementary Note 5** and **Supplementary File 9**) to test the reverse causal pathway, from 525 DEs using FinnGen and Psychiatric Genomics Consortium (PGC) data, to the 2 binary sleep traits. These analyses did not support a widespread causal effect of disease on sleep disturbances, reinforcing the interpretation of sleep disturbances as potential risk factors. We nevertheless acknowledge the possibility of bidirectional effects. For example, Pasman et al.^39^ found bidirectional causality between major depressive disorder and insomnia, but not sleep duration. In addition, some previous studies modeled sleep duration as a continuous trait using linear Mendelian randomization, which does not capture potential non-linear relationships that methods such as fractional polynomial Mendelian randomization can address^40^. To probe this further, we conducted additional sensitivity analyses focused on depression-related endpoints in our MR results. Specifically, F5_DEPRESSION_DYSTHYMIA (number of cases: 48,222) in relation to long sleep duration was not statistically significant (P-value>0.05). The sensitivity analyses results are presented in **Extended Data Figure 6**, including the per-SNP F statistics, horizontal pleiotropy and heterogeneity test based on MR-Egger, and horizontal pleiotropy from MR-PRESSO and fractional polynomial MR.

## Discussion

In this study, we reveal a robust and systemic U-shaped association between sleep duration and biological aging across multiple organ systems and omics layers. By leveraging large-scale, multimodal data from the MULTI Consortium, we demonstrate that both short (<6 hours) and long (>8 hours) sleep duration are consistently linked to elevated biological aging burden across 7 organ systems and 3 omics types. Importantly, our findings extend prior work on phenotype-based aging clocks by showing that this nonlinear sleep-aging relationship generalizes across the body and is evident not only in structural and functional imaging features but also at the molecular level. This is the first study, to our knowledge, that reveals a broad agreement between sleep duration and multi-organ, multi-omics aging clocks, and links these signatures to systemic disease outcomes and mortality risk. Our results underscore the systemic biological adverse associations of disturbed sleep and provide a compelling framework for more targeted and thoughtful attention to sleep disturbance as a potential signal of emerging health issues and a partner in the quest to promote healthy aging, reduce disease risk, and extend lifespan.

### U-shaped pattern between sleep duration and biological aging clocks beyond the brain

A key contribution of this study lies in uncovering consistent U-shaped associations between sleep duration and organ-specific biological aging clocks across diverse organs, tissues, and omics data types. At the molecular level, we observed that both short and long sleep duration are associated with accelerated pulmonary, brain, hepatic, immune, and skin ProtBAGs, suggesting that proteomic signatures of aging in these systems are particularly sensitive to sleep perturbations. These findings support sleep’s systemic implications on immune-inflammatory processes^41,42,43^, metabolic detoxification^44,45^, and neurodegenerative pathways^46^. Notably, endocrine MetBAG also exhibited a U-shaped pattern, highlighting the metabolic cost of sleep imbalance, possibly via dysregulation of hormonal homeostasis and glucose metabolism^47,48^. At the imaging level, U-shaped patterns in brain MRIBAG, adipose MRIBAG, and pancreas MRIBAG further reinforce the idea that both central and peripheral organs experience structural and functional alterations under sleep extremes^49,50,51^.

In addition to the overarching U-shaped pattern, our results reveal organ- and omics-specific variability in the sample BAG minimum values of the BAG-sleep relationship. While the overall sample minimum values centered between 6 and 8 hours, consistent with prior epidemiological recommendations^7^, the precise inflection point varied across tissues and omics types. For instance, brain ProtBAG showed the lowest aging burden around 7.7 hours of sleep, whereas brain MRIBAG reached its inflection point closer to 6.5 hours. These findings suggest that molecular aging in the brain may require longer restorative sleep than its structural counterpart, potentially reflecting the different timescales and mechanisms by which proteomic versus imaging phenotypes capture sleep-related damage or resilience^50,52^. Alternatively, this can be due to a reverse causality, where longer sleep reflects brain aging rather than causes it, pulling the right side of the U-shaped curve upward, and making the apparent sample BAG minimum values appear shorter for the brain MRIBAG (e.g., 6.5 hours). In addition, UKBB’s MRI sub-study over-represents healthier and more highly educated participants; this may attenuate or distort the right limb of the observed U-shape independent of reverse causality. In contrast, the proteomic and metabolomic BAGs are derived from broader, potentially more representative samples spanning wider health states, which could partly explain their stronger or more symmetric associations. To summarize, the differing sample minimum values of sleep durations across organ-specific clocks likely reflect the heterogeneous physiological demands and recovery processes of each organ system. For example, organs such as the brain may be more sensitive to sleep deprivation and circadian disruption, whereas metabolic or peripheral organs (e.g., liver, pancreas) may exhibit delayed or compensatory responses, resulting in distinct sleep–aging optima. Moreover, clear sex differences emerged in the relationship between sleep and biological aging. Several organs displayed significant BAG differences, BAG-by-sex interactions, divergence in the population mean, and sample minimum values of sleep time. For example, males exhibited higher brain MRIBAG, whereas females exhibited higher brain ProtBAG between sexes. This divergence likely reflects that these clocks index distinct layers of biology. The brain MRIBAG is driven by macro-structural MRI features (i.e., regional volumes) that are shaped by lifelong neurodevelopmental trajectories, hormonal, and sex-differential factors that have been repeatedly linked to sex differences in brain aging and neurodegeneration. In contrast, brain ProtBAG is derived from circulating brain-enriched proteins, which may be more sensitive to systemic inflammatory/immune signaling, endocrine regulation, blood–brain barrier permeability, and glia- and vasculature-related processes that influence the release, transport, or clearance of brain-linked proteins in plasma. These differences may reflect sex-specific hormonal regulation, immune responses, activity patterns, or metabolic demands, as well as socially unfavorable factors for females, that likely relate to how sleep duration modulates aging trajectories across organ systems – a concept supported by growing literature on sex-specific sleep physiology and aging biology^53,54,55^.

Together, these results reveal that deviations from sample minimum values of sleep duration are not only associated with the brain but also peripheral tissues and systems critical to cardiometabolic and immune regulation, emphasizing the multi-organ biological burden imposed by disrupted sleep.

### Evidence from molecular factors and imaging-derived phenotypes reinforces the U-shaped pattern of sleep duration

Our findings provide converging evidence from imaging, proteomics, and metabolomics that support the hypothesis that sleep duration exerts nonlinear effects across multiple organ systems and molecular domains. These patterns reflect the systemic nature of sleep physiology and are consistent with prior research showing that both sleep deprivation and prolonged sleep are associated with adverse outcomes across diverse biological pathways.

At the imaging level, U-shaped associations between sleep duration and organ-specific IDPs, particularly in the brain, adipose tissue, and pancreas, echo prior findings that link both short and long sleep with structural brain alterations and metabolic dysregulation (**Extended Data Figure 1**). For example, previous neuroimaging studies have shown that hippocampal atrophy is associated with both insufficient and excessive sleep, possibly reflecting neurotoxic or neurodegenerative processes triggered by circadian misalignment, impaired glymphatic clearance, or inflammatory load^49,50^. Similarly, adipose tissue dysfunction and pancreatic abnormalities underlie cardiometabolic conditions such as insulin resistance and obesity, both of which have been associated with extremes of sleep^56,57^.

At the molecular level, the U-shaped relationships observed in organ-enriched plasma proteins^58^ align with several established biological hypotheses of sleep regulation (**Extended Data Figure 2**). For instance, immune-enriched proteins involved in neutrophil chemotaxis, complement activation, and leukocyte signaling were significantly altered in individuals with suboptimal sleep. This supports the immune activation hypothesis, positing that sleep disturbances lead to, or reflect, systemic inflammation, impairing tissue repair, neurogenesis, and metabolic homeostasis^59^. Similarly, hepatic-enriched proteins involved in coagulation and innate defense pathways reinforce the liver-sleep axis, wherein disrupted sleep contributes to liver stress, oxidative damage, and altered lipid metabolism^60^. Complementing these proteomic findings, the metabolomics results revealed alterations in metabolites involved in fatty acid metabolism, ketone body synthesis, and amino acid catabolism (**Extended Data Figure 3**). For example, the observed enrichment of pathways related to neurotransmitter transport and synaptic remodeling, such as tyrosine metabolism and Na⁺/Cl⁻-dependent transporter activity, supports the neurochemical plasticity hypothesis, suggesting that sleep modulates central nervous system function via fine-tuned molecular and ionic gradients essential for neural signaling^61,62,63^.

Altogether, our multi-layered findings reinforce the notion that sleep duration is not only a behavioral or neurological variable but a deeply embedded systemic modulator. The evidence from both structural-functional imaging and circulating biomolecules converges to support the role of sleep in maintaining organ integrity, metabolic balance, and immune equilibrium. These results further reinforce sleep as an important process with far-reaching molecular and physiological implications across the body.

### Genetic and clinical evidence suggest that abnormal sleep duration is associated with systemic disease endpoints and mortality

Our integrated genetic and clinical analyses consistently demonstrate that disturbed sleep duration patterns, both short and long, are robustly linked to a wide array of systemic DEs and increased mortality risk. Genetic correlation analyses revealed significant positive associations between abnormal sleep duration patterns and over 150 disease phenotypes across cardiovascular, metabolic, musculoskeletal, psychiatric, neurological, pulmonary, and gastrointestinal systems, with short sleep duration exhibiting particularly widespread effects. These patterns were mirrored in our clinical survival analyses, where short and long sleep duration were both significantly associated with elevated hazard ratios for a broad spectrum of diseases and all-cause mortality. Importantly, the genetic and clinical evidence together suggest that short and long sleep duration may contribute to disease via distinct biological pathways: short sleep duration appears to exert more direct effects across multiple organ systems, potentially reflecting heightened physiological stress and systemic dysregulation, whereas long sleep duration shows a more focused association with neuropsychiatric and brain-related phenotypes, potentially reflecting compensatory mechanisms or underlying latent pathologies. These findings reinforce the role of sleep as a potentially modifiable risk factor in health and aging management and underscore the need for mechanistically informed strategies tailored to sleep disruption, as we further illustrated in LLD.

### Distinct mechanistic insights into short sleep duration and long sleep duration patterns in late-life depression

While LLD has been conceptualized as a brain-centric disorder, our findings suggest a more nuanced framework in which both short and long sleep duration link to LLD vulnerability via distinct mechanistic pathways involving multiple organ systems. Notably, short sleep duration appears to exert more direct effects on LLD. This pattern may reflect acute physiological stressors associated with sleep deprivation, including heightened sympathetic activity, immune dysregulation, and hypothalamic-pituitary-adrenal axis activation, all of which can be linked to mood regulation independently of brain atrophy. These findings are consistent with prior studies linking short sleep to systemic inflammation, metabolic strain, and emotional dysregulation^59,64^, pointing to a fast-acting, body-to-brain pathway^65^.

In contrast, long sleep duration appears to be associated with LLD predominantly through indirect pathways, especially via accelerated aging in the brain and adipose tissue. The strong mediation effects observed for brain and adipose MRIBAGs suggest that long sleep may not be a direct risk factor per se, but rather a marker of underlying physiological compensations or subclinical disease processes, potentially including neurodegeneration, energy imbalance, or immune exhaustion, that gradually erode mental health resilience. The finding also parallels a cultural illustration from Moshfegh’s “*My Year of Rest and Relaxation”*, where an unnamed narrator methodically increases prescription medication to achieve a year-long sleep^66^. Such patterns support emerging views of long sleep as a complex, sometimes maladaptive response to latent morbidity rather than a simple behavioral choice^67,68^. The pronounced mediational roles of the brain and other body aging clocks are particularly striking, indicating that central and systemic aging may jointly drive mood vulnerability in individuals with prolonged sleep. Importantly, the mediation of long sleep’s effect on depression was not exclusive to the brain, despite LLD’s traditional classification as a neuropsychiatric disorder. This broader multi-organ involvement suggests that depression in late life might arise from distributed physiological aging, wherein systemic decline in tissues like adipose, liver, or cardiovascular systems contributes to or exacerbates neural vulnerability. This is in line with recent multidimensional models of depression that integrate metabolic dysfunction, systemic inflammation, and neuroplasticity deficits as co-occurring drivers of depressive phenotypes^69,70^. Importantly, we used Mendelian randomization to partially rule out the possibility of reverse causality, whereby sleep disturbances may be caused by disease burden. While a previous study by Austin-Zimmerman^34^ reported a bidirectional relationship between long sleep duration and major depressive disorder, their analysis relied on a lenient P-value threshold (P-value<1×10^−5^) for selecting instrumental variables, which may cause weak instrument bias. We attempted to replicate their findings using the same PGC exposure GWAS by Wray et al.^71^ (45,396 cases, removing UKBB samples) using a genome-wide P-value threshold (P-value<5×10^−8^). After LD clumping, no independent genome-wide significant instruments remained. To further investigate this, we downloaded the most recent major depressive disorder (MDD) GWAS from FinnGen (59,333 cases). Overall, although the IVW (P = 0.003) estimator suggested a positive causal effect of genetic liability, the significant MR-Egger intercept (P=0.047), MR-PRESSO global pleiotropy (P=0.002), and high heterogeneity (e.g., P=0.0009) indicate pleiotropy-induced biases. Pleiotropy-aware methods showed attenuated effects and were sensitive to pleiotropy assumptions (balanced vs. directional pleiotropy), including MR-Egger for null/negative (P=0.24), MRMix for near-null (P=0.41), and MR-RAPS for nominally positive (P=0.019). Therefore, while suggestive, the evidence for a causal link between MDD and long sleep duration remains inconclusive and should be interpreted cautiously (**Extended Data Figure 7**).

Together, these findings highlight that while short sleep may acutely disturb mood through direct neuroimmune and neuroendocrine perturbations, long sleep may reflect an organ-mediated pathway, linking chronic subclinical aging processes to eventual neuropsychiatric decline. This distinction reinforces the need for tailored sleep interventions and underscores the importance of viewing LLD as a systemic, not merely cerebral, manifestation of aging.

### Limitations

Several limitations warrant consideration. First, although our findings revealed U-shaped associations between sleep duration and various phenotypes, these patterns require external validation in independent cohorts, potentially on the same scale as UKBB, and additional consideration of harmonization of resources from different studies and protocols. Second, the reliance on self-reported, questionnaire-based sleep duration measurement may introduce recall bias or misclassification, and future studies incorporating objective measures, such as polysomnography, are essential to better understand underlying mechanisms. Third, the cross-sectional design of this study limits our ability to determine causality or the direction of effect; although our current analysis positions sleep disturbance as a modifiable risk factor, longitudinal follow-up is needed to clarify whether sleep disturbance is a modifiable risk factor or a consequence of disease burden. Furthermore, proteomic and metabolomic signals fluctuate with time, illness, medication, and diet, so single snapshots can misclassify biology. Longitudinal sampling may yield more reliable estimates and separate transient noise from persistent, risk-relevant biology. Fourth, our analyses were restricted to individuals of predominantly European ancestry, which limits generalizability. Studies in more ethnically diverse and underrepresented populations are needed to ensure broader applicability. Fifth, despite adjusting for a wide array of covariates, residual confounding and the potential for reverse causality, particularly for long sleep as a marker of subclinical illness, cannot be fully excluded. In addition, circadian misalignment and sleep fragmentation were not directly assessed, which may influence the observed associations between sleep duration and organ-specific biological aging. Sixth, future research needs to extend the Sleep chart across the lifespan to better capture dynamic patterns of sleep duration beyond adults. Finally, while our analyses support the interpretation of sleep disturbance as a modifiable risk factor and provide no evidence for reverse causality based on Mendelian randomization, further investigation using longitudinal data is warranted to rigorously assess this relationship.

## Methods

### Method 1: The MULTI Consortium

The MULTI Consortium is an ongoing initiative to integrate and consolidate existing multi-organ data and multi-omics data, including imaging, genetics, and proteomics. Building on existing consortia and studies, MULTI aims to curate and harmonize the data to model human aging and disease at scale across the lifespan. Refer to **Supplementary Table 1** for comprehensive information, including the complete list of data analyzed and their respective sample characteristics. Participants provided written informed consent to the corresponding studies. The MULTI Consortium is approved by the Institutional Review Board at Columbia University (AAAV6751).

### UK Biobank

UKBB^72^ is a population-based research initiative comprising around 500,000 individuals from the United Kingdom between 2006 and 2010. Ethical approval for the UKBB study has been secured, and information about the ethics committee can be found here: https://www.ukbiobank.ac.uk/learn-more-about-uk-biobank/governance/ethics-advisory-committee. The main sleep data used in this study were sleep duration (Field ID: 1160) based on a self-reported questionnaire collected from all 500,000 participants at baseline. The 7 brain MRIBAGs were derived from multi-organ MRI data at the second visit, 11 ProtBAGs, and 5 MetBAGs were derived using plasma proteomics and metabolomics at baseline. Finally, we also included individual plasma proteins and metabolites in our ProWAS and MetWAS, as detailed below.

For the primary variable of interest in UKBB, sleep duration (Data-Field: 1160) was assessed using an ACE touchscreen questionnaire asking: “About how many hours sleep do you get in every 24 hours? (please include naps).” Participants entered a numeric value, which underwent basic quality control: responses of less than 1 hour or more than 23 hours were rejected, and values below 3 hours or above 12 hours triggered a confirmation prompt. If participants clicked the Help button, they were instructed that, if their sleep duration varied substantially, they should report the average number of hours slept in a 24-hour day over the past 4 weeks. For this variable, the value −1 indicates “Do not know” and the value −3 indicates “Prefer not to answer, which were excluded in the current work. For the multi-organ IDPs, we used multi-organ MRI data from 7 organ systems and tissues (Category ID: 100003), including the brain, heart, liver, pancreas, spleen, adipose, and kidney. Specifically, the MUSE atlas-derived brain IDPs from the T1-weighted MRI^13^ were used for the brain MRIBAG generation. We also used neural networks to analyze the raw cardiac MRI images in our previous study^21^ and returned them to the UKBB to extract heart-specific IDPs (Category ID: 157), which were used to derive the heart MRIBAG. For the other organs’ IDPs, we used the pre-derived features from the UKBB (Category ID: 105). For the plasma proteomics data, we downloaded the original data (Category ID: 1838), which were analyzed and made available to the community by the UK Biobank Pharma Proteomics Project (UKB-PPP^73^). The initial quality control procedures were described in the original work^74^; we conducted additional quality-check steps as outlined in **Method 5**. We also imputed missing normalized protein expression (NPX) values and defined the organ-specific proteins using the HPA platform (https://www.proteinatlas.org/), as detailed in our previous work. For the plasma metabolomics data, we downloaded the original data (Category ID: 220), which were analyzed and made available to the community by Nightingale Health Plc in the UKBB. Additional quality check analyses were performed as detailed in **Method 6**. **Supplementary Table 1** provides a detailed list of subsample characteristics corresponding to each data pattern.

### FinnGen

The FinnGen^75^ study is a large-scale genomics initiative that has analyzed over 500,000 Finnish biobank samples and correlated genetic variation with health data to understand disease mechanisms and predispositions. The project is a collaboration between research organizations and biobanks within Finland and international industry partners. For the benefit of research, FinnGen generously made its GWAS findings accessible to the wider scientific community (https://www.finngen.fi/en/access_results). This research utilized the publicly released GWAS summary statistics (version R9), which became available on May 11, 2022, after harmonization by the consortium. No individual data were used in the current study.

FinnGen published the R9 version of GWAS summary statistics via REGENIE software (v2.2.4)^76^, covering 2272 DEs, including 2269 binary traits and 3 quantitative traits. The GWAS model included covariates such as age, sex, the initial 10 genetic principal components, and the genotyping batch. Genotype imputation was referenced on the population-specific SISu v4.0 panel. We included GWAS summary statistics for 521 FinnGen DEs in our analyses.

### Psychiatric Genomics Consortium

PGC^77^ is an international collaboration of researchers studying the genetic basis of psychiatric disorders. PGC aims to identify and understand the genetic factors contributing to various psychiatric disorders such as schizophrenia, bipolar disorder, major depressive disorder, and others. The GWAS summary statistics were acquired from the PGC website (https://pgc.unc.edu/for-researchers/download-results/), underwent quality checks, and were harmonized to ensure seamless integration into our analysis. No individual data were used from PGC. Each study detailed its specific GWAS models and methodologies, and the consortium consolidated the release of GWAS summary statistics derived from individual studies. In the current study, we included summary data for 6 brain diseases (**Supplementary Table 7**).

### TriNetX

To evaluate real-world clinical outcomes associated with sleep traits, we leveraged the TriNetX^78^ database (https://trinetx.com/), a global federated health research platform that provides access to de-identified electronic medical records from over 70 healthcare organizations, encompassing more than 90 million patients. The TriNetX platform integrates clinical data, including diagnoses, medications, procedures, and laboratory results, enabling large-scale observational analyses. We used this resource to assess associations between insomnia and hypersomnia and systemic disease outcomes across organ systems identified in UKBB (**Fig. 3**).

### Baltimore Longitudinal Study of Aging

The main goal of the BLSA is to understand the normal aging process. Tracking physiological and cognitive changes over time aims to identify risk factors for age-related diseases, study patterns of decline, and discover predictors of healthy aging. BLSA^79,80^ brain MRI, self-reported, and actigraphy-derived sleep duration (*N*=385) were used to compare and replicate the U-shaped pattern observed in the main analysis for the brain MRIBAG. For self-reported sleep duration, sleep duration is assessed using a standardized questionnaire item asking: “On average, in the past month, how many hours of sleep did you get each night?” Participants select from ordered categorical response options reflecting typical nightly sleep duration: more than 7 hours, more than 6 up to 7 hours, more than 5 up to 6 hours, or 5 hours or fewer. This measure captures habitual sleep duration over the prior month and represents participants’ perceived average nightly sleep. We also included participants who underwent wrist actigraphy. They were instructed to wear an Actiwatch-2 wrist actigraphy (Philips-Respironics, Bend, OR) on their nondominant wrist for seven consecutive 24-hour periods. The devices continuously recorded activity counts and ambient light levels, and participants were asked to press the event marker each time they intended to go to sleep (“lights out”) and when they got up to start their day^81^.

### The Multi-Ethnic Study of Atherosclerosis

The Multi-Ethnic Study of Atherosclerosis (MESA^27,28,82^) is a medical research initiative involving over 6,000 men and women from six U.S. communities. For this analysis, we included 573 participants with available brain MRI data, self-reported sleep duration measures, and relevant covariates such as age, sex, and BMI. We leveraged the MESA cohort to attempt replication of the U-shaped association observed in UKBB for the brain MRIBAG. Self-reported sleep duration is obtained from the Exam 5 Sleep Questionnaire, in which participants report their habitual bedtimes and wake times separately for weekdays and weekends. The questionnaire includes items asking for bedtime and waketime in 24-hour format, from which MESA derives weekday sleep duration and weekend sleep duration expressed in hours or hours-and-minutes.

### Method 2: GAM models the relationship between sleep duration and organ aging clocks, IDPs, proteins, and metabolites

To model the nonlinear association between sleep duration and the 23 multi-organ BAGs and any other phenotype, we implemented GAMs using the *mgcv* package in R. This approach enabled flexible modeling of complex relationships, whether linear, flat, sigmoidal, or U-shaped, without requiring any prior assumptions about the underlying shape of the fitted curves. We adjusted for key demographic and physiological covariates (i.e., age, sex, weight, height, waist circumference, BMI, assessment center, diastole, systolic blood pressure, time differences for data collection (for MRIBAGs), and disease status). Participants reporting extremely short durations or those with missing sleep duration were excluded. The analysis was restricted to those reporting 4-10 hours of sleep to reduce the influence of outliers. For each BAG, we fitted GAM with cubic regression splines (*bs=’cr’*) to smooth the nonlinear association between sleep duration and the BAG, considering sex and sex-sleep interaction term.

Model selection was conducted by systematically evaluating combinations of smoothing dimensions (k = 3, 5, 10, 15, 20) and distribution families (Gaussian, t, and Gamma). For each candidate model, the estimated EDF and smoothing parameters were optimized internally via penalized maximum likelihood estimation. The optimal model was defined as the one yielding the lowest Akaike Information Criterion, indicating the best balance between model fit and complexity, while ensuring that the EDF did not approach the upper limit of the specified k, thereby avoiding overfitting. We tested *i*) the main effect (sleep duration; P_1_), *ii*) sex difference in population mean (P_2_), and *iii*) sex-sleep interaction terms (P_3_) on each BAG. The solid curves in **Fig. 1** depict estimated BAG, while shaded bands represent the 95% confidence interval (CI). **Supplementary Figure 1** provides diagnostic plots of model fit for the optimal model. The raw BAG values were normalized to the range (0, 1) to allow for the application of different family distributions in the model.

### Method 3: 23 Multi-organ aging clocks

In our previous work, we processed raw brain MRI data from the UK Biobank to extract 119 gray matter (GM) regions of interest (ROIs) from T1-weighted images, which were used to compute the brain MRIBAG. For the heart MRIBAG, we utilized 80 cardiac MRI-derived traits from Bai et al.^21^, which we had incorporated in an earlier study. Additionally, five abdominal MRIBAGs were derived from abdominal MRI data (Category ID = 105) across multiple studies^83,84,85,86,87,88^, yielding a total of 7 MRIBAGs^1^. We also developed 11 ProtBAGs using UKBB plasma proteomics data^2^, along with 5 MetBAGs^3^ based on plasma metabolomics profiles. All 23 BAGs were developed using a nested cross-validation framework, adhering to best practices in machine learning to minimize overfitting and prevent data leakage^89,2^.

In our previous studies^1,2,3^, we described in detail the population definition and cross-validation procedures used for model training, which we summarize here. Applying a coherent machine learning framework, we assessed the performance of age-prediction models. Hyperparameter tuning was performed through nested, repeated holdout cross-validation with 50 repetitions (80% training/validation and 20% testing). Specifically, we performed a grid search for fine-tuning model-specific hyperparameters. The within-distribution, holdout test dataset was held out to unbiasedly evaluate model performance (different from the 20% test dataset from the cross-validation).

To rigorously train the age-prediction models, we first defined participants without any pathologies based on ICD code and clinical history as CN. We further split the CN into the following datasets:

- CN within-distribution, holdout test dataset: 500 participants were randomly drawn from the CN population. Within-distribution, holdout test datasets are ideal for objectively evaluating machine learning model performance, especially in studies with large sample sizes, such as the UK Biobank.
- CN training/validation dataset: 80% of the remaining CN population was used for the inner loop 10-fold cross-validation for hyperparameter selection.
- CN cross-validated test dataset: 20% of the remaining CN population was used for the outer-loop 50 repetitions.
- Patient dataset: All patients who have at least one ICD-10-based diagnosis or clinical history.

The CN training/validation/test datasets were used for model development and were employed with a nested cross-validation procedure for all machine learning models (LASSO regression and support vector regressor, elastic net, and neural network), whereas the within-distribution, holdout test set provided an unbiased assessment of model performance. Model evaluation metrics included MAE and Pearson’s *r*. Age bias correction was applied using the approach outlined by Beheshti et al.^90^.

### Method 4: IDP-wide associations

We assessed the association between sleep duration and 720 image-derived phenotypes (IDPs) covering 8 organ systems and tissues using UKBB *in vivo* imaging data. For each IDP, we fitted the same GAM as in **Method 2** to capture the nonlinear associations with sleep duration. These models included age, sex, body mass index, height, weight, waist circumference, assessment center, blood pressure, and disease status as covariates. Additionally, organ-specific covariates were incorporated, such as brain positioning in the scanner (lateral, transverse, and longitudinal), head motion, and intracranial volume for the brain IDPs. Outlier values in the IDP outcome variables, defined as ±4 standard deviations from the mean, were removed to minimize the influence of extreme values. For each IDP, we extracted effect estimates and tested for the main effects of sleep, sex differences, and sex-specific interactions. Predicted curves and 95% confidence intervals were generated separately for males and females. When significant (P<0.05/720), sex-specific turning points (sample minimum values of sleep duration) were also identified from the fitted curves. Descriptions of these 720 IDPs are provided in our previous study^1,16^ and in **Supplementary File 2**.

### Method 5: Proteome-wide associations

Our analysis focused on the first instance of the proteomics data (“instance” = 0). We then integrated Olink files containing coding information, batch numbers, assay details, and limit of detection (LOD) data (Category ID: 1839) by matching them to the proteomics dataset ID. Finally, we excluded Normalized Protein eXpression (NPX) values that fell below the protein-specific LOD. Descriptions of these proteins are provided in our previous study^2^ and in **Supplementary File 3**.

We conducted ProWASs by linking sleep duration to 2923 unique plasma proteins measured using the same GAM model. The GAM was adjusted for common covariates, including age, sex, weight, height, waist circumference, BMI, assessment center, disease status, diastolic and systolic blood pressure, protein batch number, limit of detection, and the first 40 genetic principal components. Multiple testing corrections were applied using Bonferroni adjustment (P < 0.05/2923). Given the substantial correlation structure among proteomic measures, we acknowledge this choice is conservative; we retain Bonferroni correction in the main text to minimize false positives. To identify and exclude extreme outliers, we defined an upper threshold as the mean plus 4 times the standard deviation for each protein. We mainly focus on the 342 organ-enriched proteins defined in our previous study^2^.

### Method 6: Metabolome-wide associations

The original data (Category ID: 220) were *i*) calibrated absolute concentrations (or ratios) and not raw NMR spectra, and *ii*) before release, had already been subject to quality control procedures by Nightingale Health Plc^91^. Following the additional procedures described in Ritchie et al.^92^, we performed additional quality check steps to remove a range of unwanted technical variations, including shipping batch, 96-well plate, well position, aliquoting robot, and aliquot tip. We focused our analysis on the first instance of the metabolomics data (“instance”=0). The analysis included 327 metabolites (comprising both small molecules and lipoprotein measures), of which 107 were non-derived metabolites, and the remainder were composite metabolites, across 274,247 participants. Descriptions of these metabolites are provided in our previous study^3^ and in **Supplementary File 4**.

We conducted MetWASs by linking sleep duration to 327 plasma metabolites. The GAM controlled common covariates, including age, sex, weight, height, waist circumference, BMI, assessment center, disease status, diastolic and systolic blood pressure, and the first 40 genetic principal components. Multiple testing corrections were applied using Bonferroni adjustment (P < 0.05/327). To identify and exclude extreme outliers, we defined an upper threshold as the mean plus 4 times the standard deviation for each metabolite. We mainly focus on the 107 organ-associated metabolites defined in our previous study^3^.

### Method 7: Genetic analyses

We used the genotype and imputed genotype data from UKBB for all genetic analyses. Our quality check pipeline focused on European ancestry in UKBB (6,477,810 SNPs passing quality checks). We summarize our genetic quality check steps. First, we excluded related individuals (up to 2^nd^-degree) from the complete UKBB sample using the KING software for family relationship inference^93^. We then removed duplicated variants from all 22 autosomal chromosomes. Individuals whose genetically identified sex did not match their self-acknowledged sex were removed. Other exclusion criteria were: *i*) individuals with more than 3% of missing genotypes; *ii*) variants with minor allele frequency (MAF; dosage mode) of less than 1%; *iii*) variants with larger than 3% missing genotyping rate; *iv*) variants that failed the Hardy-Weinberg test at 1×10^−10^. To further adjust for population stratification,^94^ we derived the first 40 genetic principal components using the FlashPCA software^95^. Details of the genetic quality check protocol are described elsewhere^96,13,97,11,98^.

#### (a) GWAS

Given the large sample sizes for both short vs. normal (16,872 short and 300,420 normal) and long vs. normal (25,049 long and 300,420 normal) sleep duration comparisons, we employed REGENIE^76^ for GWAS, as it is well-suited for large-scale genetic analyses due to its computational efficiency and ability to control for population structure and relatedness, outperforming alternatives like PLINK and fastGWA in these settings. Our GWAS adjusted common covariates, including age, disease status, age-squared, sex, interactions of age with sex, BMI, waist circumference, standing height, weight, systolic/diastolic blood pressure, and the first 40 genetic principal components. We applied a genome-wide significance threshold (5×10^−8^) to annotate the significant independent genomic loci.

##### Annotation of genomic loci

For all GWASs, genomic loci were annotated using FUMA^99^. For genomic loci annotation, FUMA initially identified lead SNPs (correlation *r*^2^ ≤ 0.1, distance < 250 kilobases) and assigned them to non-overlapping genomic loci. The lead SNP with the lowest P-value (i.e., the top lead SNP) represented the genomic locus. Further details on the definitions of top lead SNP, lead SNP, independent significant SNP, and candidate SNP can be found in **Supplementary Note 2**.

#### (b) MAGMA Tissue Expression Analysis

MAGMA gene-property analysis^31^ was conducted using gene expression data from the GTEx (v8) to investigate tissue-specific associations with genetic variants. Unlike differential gene expression enrichment tests focusing solely on prioritized genes, MAGMA leverages the full distribution of SNP p-values across the genome, providing a more comprehensive assessment of how genetic signals relate to gene expression patterns in various tissues.

#### (d) Genetic correlation

We estimated the genetic correlation (*g_c_*) using the LDSC^30^ software between the two abnormal sleep duration patterns and 527 DEs from the FinnGen and PGC datasets. We employed precomputed LD scores from the 1000 Genomes of European ancestry, maintaining default settings for other parameters in LDSC. It’s worth noting that LDSC corrects for sample overlap, ensuring an unbiased genetic correlation estimate^100^. Statistical significance was determined using Bonferroni correction.

### Method 8: Survival analyses for risk of disease endpoints and all-cause mortality

In the UKBB, we evaluated the predictive value of abnormal sleep duration patterns, specifically short sleep (<6 hours) and long sleep (>8 hours), through two sets of analyses: *i*) survival analysis to assess the risk of time to incident single disease outcomes defined by ICD-10 codes, and *ii*) survival analysis to estimate the risk of all-cause mortality.

#### (a) Survival analysis for ICD-based single disease endpoint

we employed a Cox proportional hazard model while adjusting for covariates to test the associations of short/long sleep duration, compared to normal sleep duration ([6-8] hours), with the time to incident of ICD-based single disease entities. Notably, we excluded individuals with any prior disease diagnosis (except the disease of interest) to ensure the analysis was restricted to a disease-free population. The covariates, age, sex, body mass index, height, weight, waist circumference, and blood pressure, were included as additional right-side variables in the model. To train the model, the “time” variable was determined by calculating the difference between the date of diagnosis of the disease for cases (or the censoring date for non-cases) and the date attending the assessment center (Field ID: 53). Participants who were diagnosed for a specific disease of interest after enrolling in the study were classified as cases; non-cases were defined by participants without any disease diagnoses.

#### (b) Survival analysis for mortality risk

we employed a Cox proportional hazard model while adjusting for covariates(i.e., age and sex) to test the associations of short/long sleep duration patterns with all-cause mortality. The covariates, age, sex, body mass index, height, weight, waist circumference, and blood pressure, were included as additional right-side variables in the model. The hazard ratio (HR), exp(*β_R_*), was calculated and reported as the effect size measure that indicates the influence of each biomarker on the risk of mortality. To train the model, the “time” variable was determined by calculating the difference between the date of death (Field ID: 40000) for cases (or the censoring date for non-cases) and the date attending the assessment center (Field ID: 53). Participants who passed away after enrolling in the study were classified as cases.

### Method 9: Structural equation modeling for mediation analysis

Using UKBB data, we used SEM^16^ to examine whether organ-specific MRIBAGs, measured at the second imaging visit (2014+), serve as mediators in the relationship between sleep duration, assessed at the baseline visit (2007-2010), and two distinct subtypes of late-life depression (LLD1 and LLD2), also evaluated at the second visit. Sleep duration was categorized into binary groups reflecting short (<6 hours) and long sleep (>8 hours) patterns, with respect to the normal sleep duration ([6-8] hours).

For each MRIBAG, we specified a mediation model (sleep duration → MRIBAG → LLD) that included: *i*) a direct path from sleep duration to LLD subtype (c2), and *ii*) an indirect path from sleep duration to MRIBAG (a1), and from MRIBAG to LLD subtype (c1), with the product term (a1×c1) representing the mediated (indirect) effect. Models were adjusted for relevant covariates, including age at assessment, sex, weight, standing height, waist circumference, BMI, diastolic blood pressure, and systolic blood pressure. To assess the robustness of causal directionality considering the time-ordering of events, we also tested reversed models (sleep duration → LLD1/2 → brain MRIBAG) to scrutinize potential inverse mediation. Significance was determined using a Bonferroni-corrected threshold (P < 0.05/7) to account for multiple comparisons across the 7 organ systems. All model estimates, including direct, indirect, total effects, and proportion mediated, are reported in the Supplementary Materials.

### Method 10: Mendelian randomization tests whether disease endpoints are causally linked to short or long sleep duration relative to normal sleep duration

We conducted two-sample Mendelian randomization linking 525 DEs (as exposures) from FinnGen and PGC to short and long sleep duration as outcomes; however, limited statistical power prevented testing the reverse direction (sleep duration → DEs), which was partially tested by the mediation analysis.

We used a two-sample Mendelian randomization approach implemented in the *TwoSampleMR* package^101^ to infer the causal relationships. We employed five distinct Mendelian randomization methods, including the inverse variance weighted (IVW) method, Egger, weighted median, simple mode, and weighted mode estimators. The STROBE-MR Statement^102^ guided our analyses to increase transparency and reproducibility, encompassing the selection of exposure and outcome variables, reporting statistics, and implementing sensitivity checks to identify potential violations of underlying assumptions. First, we performed an unbiased quality check on the GWAS summary statistics. Notably, the absence of population overlapping bias^103^ was confirmed, given that FinnGen and UKBB participants largely represent populations of European ancestry without explicit overlap with UKBB. PGC GWAS summary data were ensured to exclude UKBB participants. Furthermore, all consortia’s GWAS summary statistics were based on or lifted to GRCh37. Subsequently, we selected the effective exposure variables by assessing the statistical power of the exposure GWAS summary statistics in terms of instrumental variables (IVs), ensuring that the number of IVs exceeded 7. Crucially, the function “*clump_data*” was applied to the exposure GWAS data, considering LD. The function “*harmonise_data*” was then used to harmonize the GWAS summary statistics of the exposure and outcome variables. Bonferroni correction was applied to all tested traits based on the number of effective DEs (*N*=179).

Finally, we conducted multiple sensitivity analyses. First, we conducted a heterogeneity test to scrutinize potential violations of the IV’s assumptions. To assess horizontal pleiotropy, which indicates the IV’s exclusivity assumption^104^, we utilized a funnel plot, single-SNP Mendelian randomization methods, and the Egger estimator. Furthermore, we performed a leave-one-out analysis, systematically excluding one instrument (SNP/IV) at a time, to gauge the sensitivity of the results to individual SNPs. Of note, our Mendelian randomization analyses rely on the standard assumption of linear genetic effects on sleep duration, providing an average causal effect per unit increment in the exposure. As such, we conducted the sleep GWAS using binary traits (long sleep vs. normal sleep and short sleep vs. normal sleep), rather than treating sleep duration as a continuous variable. Nevertheless, the MR estimates do not directly represent the nonlinear U-shaped sleep-BAG relationships observed in our phenotypic analyses and should be interpreted as complementary to, rather than a direct mirror of, the observational sleep–BAG associations.

## Data Availability

The GWAS summary statistics for long and short sleep duration are publicly available via the Sleep Chart portal (https://labs-laboratory.com/sleepchart). The GWAS summary statistics for the 23 multi-organ BAGs are publicly accessible via the MEDICINE portal (https://labs-laboratory.com/medicine/). All GWAS summary statistics are also available at Synapse (https://www.synapse.org/Synapse:syn64923248/wiki/630992^105^ and https://www.synapse.org/Synapse:syn68602065/wiki/633329^106^). Our study used data generated by the Human Protein Atlas (https://www.proteinatlas.org) and MetaboAnalyst (https://www.metaboanalyst.ca/MetaboAnalyst/ModuleView.xhtml). GWAS summary data for the DEs were downloaded from the official websites of FinnGen (R9: https://www.finngen.fi/en/access_results) and PGC (https://pgc.unc.edu/for-researchers/download-results/). Individual data from UKBB can be requested with proper registration at https://www.ukbiobank.ac.uk/. The data used to replicate the main signal in UKBB comes from TriNetX (https://trinetx.com/), which requires a valid registration and request. We also shared the original PDF for the 4 main figures and the R script for the GAM modeling at: https://zenodo.org/records/17409426.

## Code Availability

The software and resources used in this study are all publicly available:

- MLNI^107^ (v0.1.2): https://github.com/anbai106/mlni, BAG generation;
- MGCV (v1.9.1) with R v4.2.2: https://cran.r-project.org/web/packages/mgcv/index.html, nonlinear GAM modeling; the R script to reproduce the U-shaped GAM modeling is available in the online supplementary section.
- GAMLSS (v5.4.22) with R v4.2.2: https://www.gamlss.com/; replicate the 9 sleep-BAG signals identified by the GAM modeling.
- Lifelines (v0.27.8): https://lifelines.readthedocs.io/en/latest/, survival analysis;
- Lavaan (v0.6.19): https://lavaan.ugent.be/, mediation analysis;
- String (v12.0): https://string-db.org/, protein set enrichment analyses and network analysis;
- MetaboAnalyst (v6.0): https://www.metaboanalyst.ca/, metabolite enrichment analysis and network analysis;
- REGENIE (v4.1): https://rgcgithub.github.io/regenie/, binary GWAS;
- LDSC (git version aa33296): https://github.com/bulik/ldsc, genetic correlation
- FUMA (v1.8.0): https://fuma.ctglab.nl/, Gene mapping, genomic locus annotation

## Authors’ contributions

Dr. Wen has full access to all the study data and is responsible for its integrity and accuracy.

*Study concept and design*: W.J.

*Drafting of the manuscript*: W.J.

*Critical revision of the manuscript for important intellectual content*: All authors

*Analysis*: W.J. ran all analyses. W.J., O.K.C., and S.Z. developed the Sleep Chart portal. A.F. ran analyses for TriNetX.

## Ethics Statement

All data used in this study were obtained from previously approved research cohorts and biobanks. The MULTI Consortium has been approved by the Institutional Review Board at Columbia University (IRB protocol: AAAV6751). Individual contributing studies received approval from their respective institutional review boards. All research was performed in accordance with relevant guidelines and regulations, and written informed consent was obtained from all participants in each study.

## Additional Information

Correspondence and requests for materials should be addressed to Dr. Junhao Wen.

## Acknowledgments

This research is supported through funding from the NIH-supported MULTI Consortium (W.J.; grant number: RF1AG092412). The MULTI consortium (J.W.; UK Biobank Application Number: 647044) aims to integrate multi-organ imaging and multi-omics data to advance our understanding of human aging and disease mechanisms. This study used the UK Biobank resource under Application Numbers 647044 (W.J.) and 35148 (D.C.; NIA grant number: RF1 AG054409). D.R.M., W.A.K., S.M.E., R.M.S., and F.L. are supported by the Intramural Research Program of the NIH, NIA. We want to express our sincere gratitude to the UK Biobank team for their invaluable contribution to advancing clinical research in our field (https://www.ukbiobank.ac.uk/). We want to acknowledge the participants and investigators of the FinnGen study and the PGC consortium, and we thank FinnGen (https://www.finngen.fi/en) and PGC (https://pgc.unc.edu/) for their generosity in sharing the GWAS summary statistics with the scientific community. We thank the BLSA participants and staff for their participation and continued dedication. The BLSA protocol was approved by the Institutional Review Board of the National Institute of Environmental Health Science, National Institutes of Health (03AG0325). We acknowledge the Multi-Ethnic Study of Atherosclerosis (MESA) and the MESA SHARe project, conducted and supported by the National Heart, Lung, and Blood Institute (NHLBI) in collaboration with MESA investigators, under contracts 75N92025D00022, 75N92020D00001, HHSN268201500003I, N01-HC-95159, 75N92025D00026, 75N92020D00005, N01-HC-95160, 75N92020D00002, N01-HC-95161, 75N92025D00024, 75N92020D00003, N01-HC-95162, 75N92025D00027, 75N92020D00006, N01-HC-95163, 75N92025D00025, 75N92020D00004, N01-HC-95164, 75N92025D00028, 75N92020D00007, N01-HC-95165, N01-HC-95166, N01-HC-95167, N01-HC-95168, and N01-HC-95169, with additional support from NHLBI grant R01HL127659 and NCATS grants UL1-TR-000040, UL1-TR-001079, and UL1-TR-001420. The MESA Sleep Ancillary Study was funded by NIH/NHLBI grant R01HL098433. The National Sleep Research Resource was supported by NHLBI (R24HL114473, 75N92019R002).

## Extended figures

**Extended Data Figure 1:**
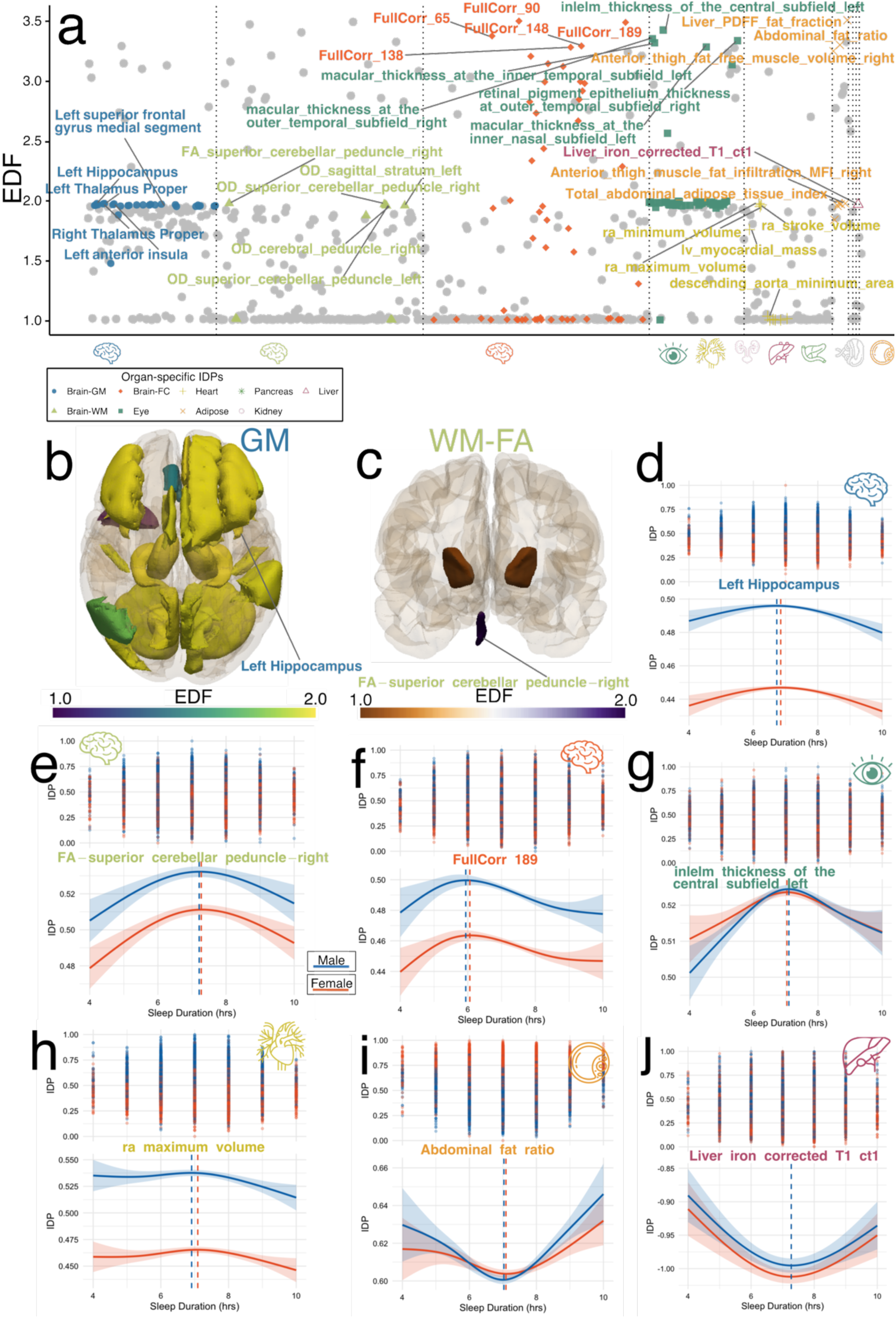
Sleep Chart delineates U-shaped associations between sleep duration and 720 organ-specific imaging-derived phenotypes. **a**) Sleep duration was associated with 720 organ-specific imaging-derived phenotypes (IDP; x-axis) using the GAM model. Non-gray colored IDPs indicate significant associations after Bonferroni correction for multiple comparisons across all available IDPs (P<0.05/720). The y-axis shows the effective degrees of freedom (EDF), quantifying the degree of nonlinearity for the U-shaped patterns, with representative proteins annotated for each organ. **b**) The EDF brain map displays the EDF values for significant signals associated with brain IDPs derived from T1-weighted MRI. **c**) The EDF brain map displays the EDF values for significant signals associated with brain IDPs derived from fractional anisotropy (FA) using diffusion MRI. **d-j**) Representative IDPs from various organ systems and tissues illustrate U-shaped associations with sleep duration.

**Extended Data Figure 2:**
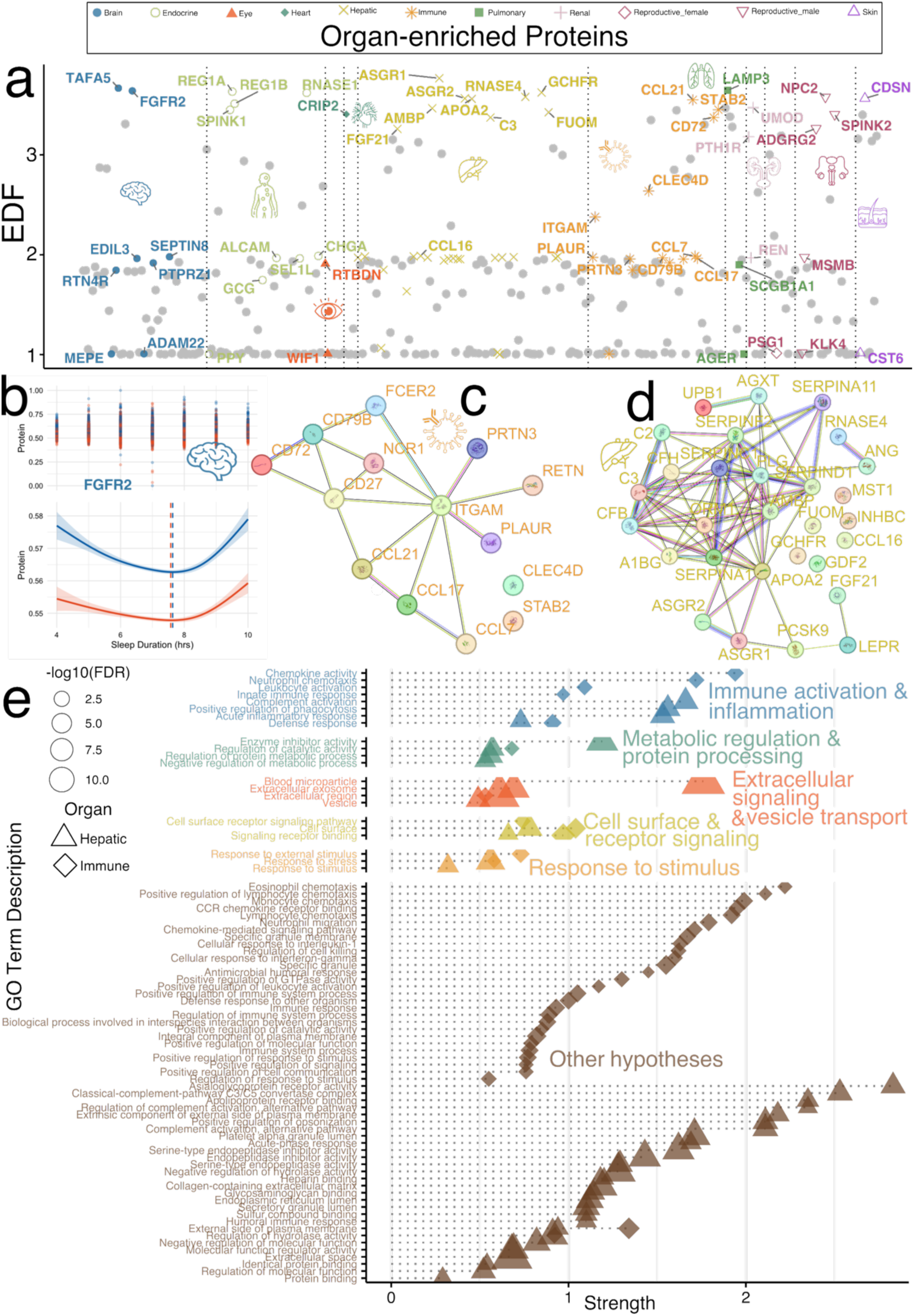
Sleep Chart delineates U-shaped associations between sleep duration and 342 organ-enriched proteins. **a**) Sleep duration was associated with 342 organ-enriched plasma proteins (x-axis) using the GAM model. These organ/tissue-enriched proteins were defined in our previous study^2^ via resources from the Human Protein Atlas (https://www.proteinatlas.org/) project. Non-gray colored proteins indicate proteins with significant associations after Bonferroni correction for multiple comparisons across all available proteins (P<0.05/2923 for all available proteins). The y-axis shows the effective degrees of freedom (EDF), quantifying the degree of nonlinearity (e.g., U-shaped associations), with representative proteins annotated for each organ. **b**) Representative U-shaped relationship between the FGFR2 protein and sleep duration. **c-d**) Protein-protein interaction (PPI) network was conducted using STRING v12.0 (https://string-db.org/) to examine the functional relationships among 14 immune-enriched and 29 hepatic-enriched proteins. STRING integrates evidence from experimental data, computational predictions, and literature text mining, enabling the PPI network to reveal how these proteins may functionally interact within and across biological pathways, potentially offering insights into shared mechanisms and tissue-specific processes. **e**) We performed protein set enrichment analysis (PSEA) using STRING for organs with more than 9 significant proteins (i.e., the hepatic and immune) to identify over-represented Gene Ontology (GO) terms across biological processes, molecular functions, and cellular components. Icon sizes in the visualization reflect the statistical significance, represented by the −log_10_(FDR-corrected P). The strength measurement (x-axis) is a log-scale enrichment ratio that shows the magnitude of enrichment. We group distinct GO terms along the y-axis by associating each biological pathway with potential sleep-related hypotheses informed by prior literature, such as the immune activation and inflammation hypothesis^42^ or the Extracellular signaling and vesicle transport hypothesis^108^.

**Extended Data Figure 3:**
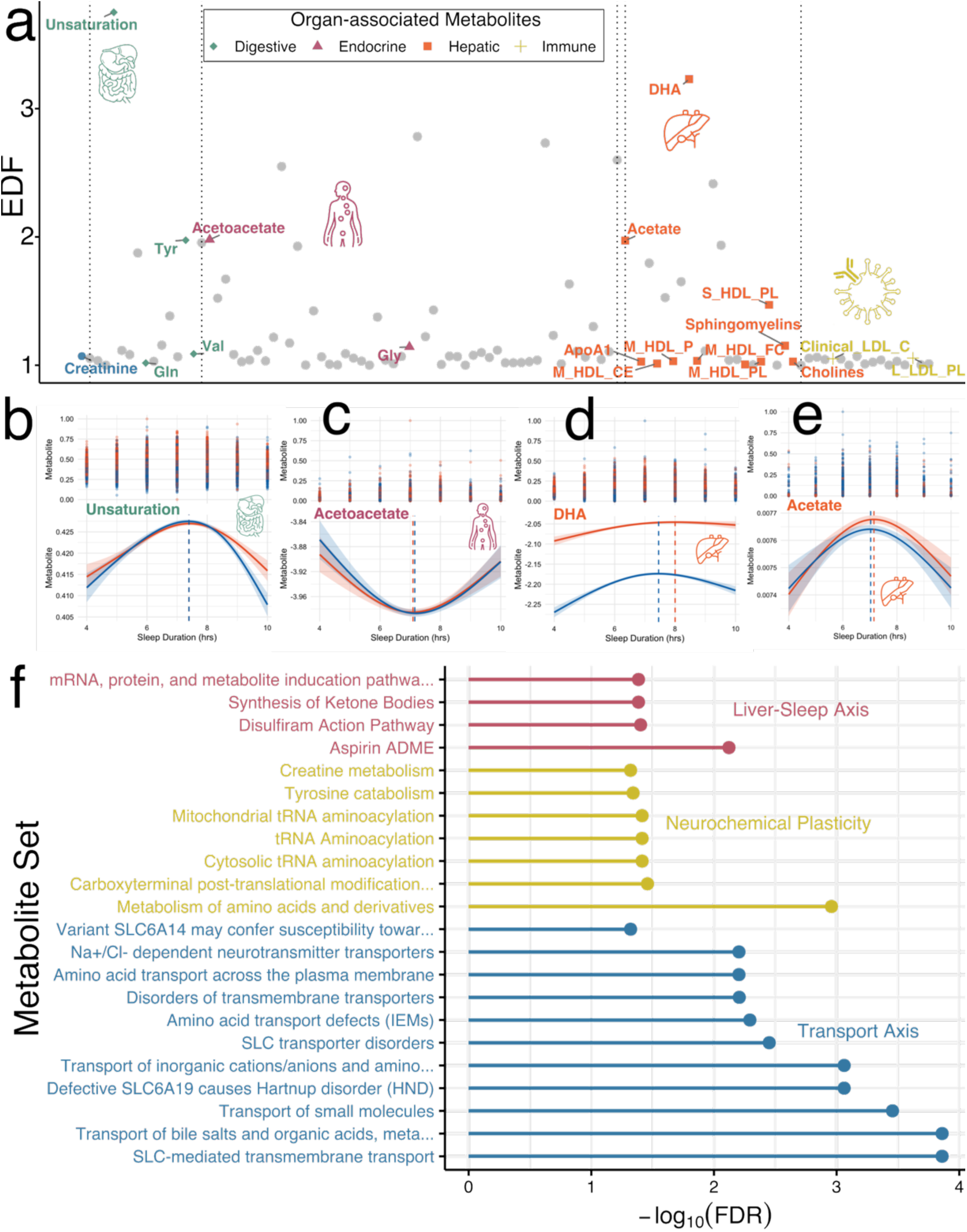
Sleep Chart delineates U-shaped associations between sleep duration and 107 organ-associated metabolites. **a**) Sleep duration was associated with 107 organ-associated plasma metabolites (x-axis) using the GAM model. These organ-associated metabolites were defined in our previous study^3^. Non-gray colored points indicate significant associations after Bonferroni correction for multiple comparisons across all available metabolites (p₁ < 0.05/327 for all available metabolites). The y-axis displays the effective degrees of freedom (EDF), quantifying the extent of nonlinearity for the U-shaped associations. We annotated representative proteins for each organ system. **b-e**) Representative U-shaped relationship between 4 metabolites or lipid subclasses and sleep duration. **f)** Metabolite set enrichment analysis (MSEA) was performed using the Over Representation Analysis (ORA) module in MetaboAnalyst (https://www.metaboanalyst.ca/), which identifies biologically meaningful patterns by assessing whether known metabolite sets (e.g., significant metabolites related to sleep duration) are overrepresented among the list of input metabolites. The analysis uses a hypergeometric test to calculate statistical significance, with multiple testing correction applied using the FDR method. Only metabolite sets with FDR < 0.05 were considered significantly enriched. Only small-molecule metabolites, excluding lipid subclasses, that could be mapped in MetaboAnalyst were included. As the background reference, we used 3,694 metabolite and lipid pathways curated in RaMP-DB (integrating KEGG via HMDB, Reactome, and WikiPathways). We organize distinct metabolite sets along the y-axis by linking them to potential sleep-related hypotheses grounded in prior literature, including the sleep-liver axis, which is closely tied to the energy hypothesis^109^ of sleep.

**Extended Data Figure 4:**
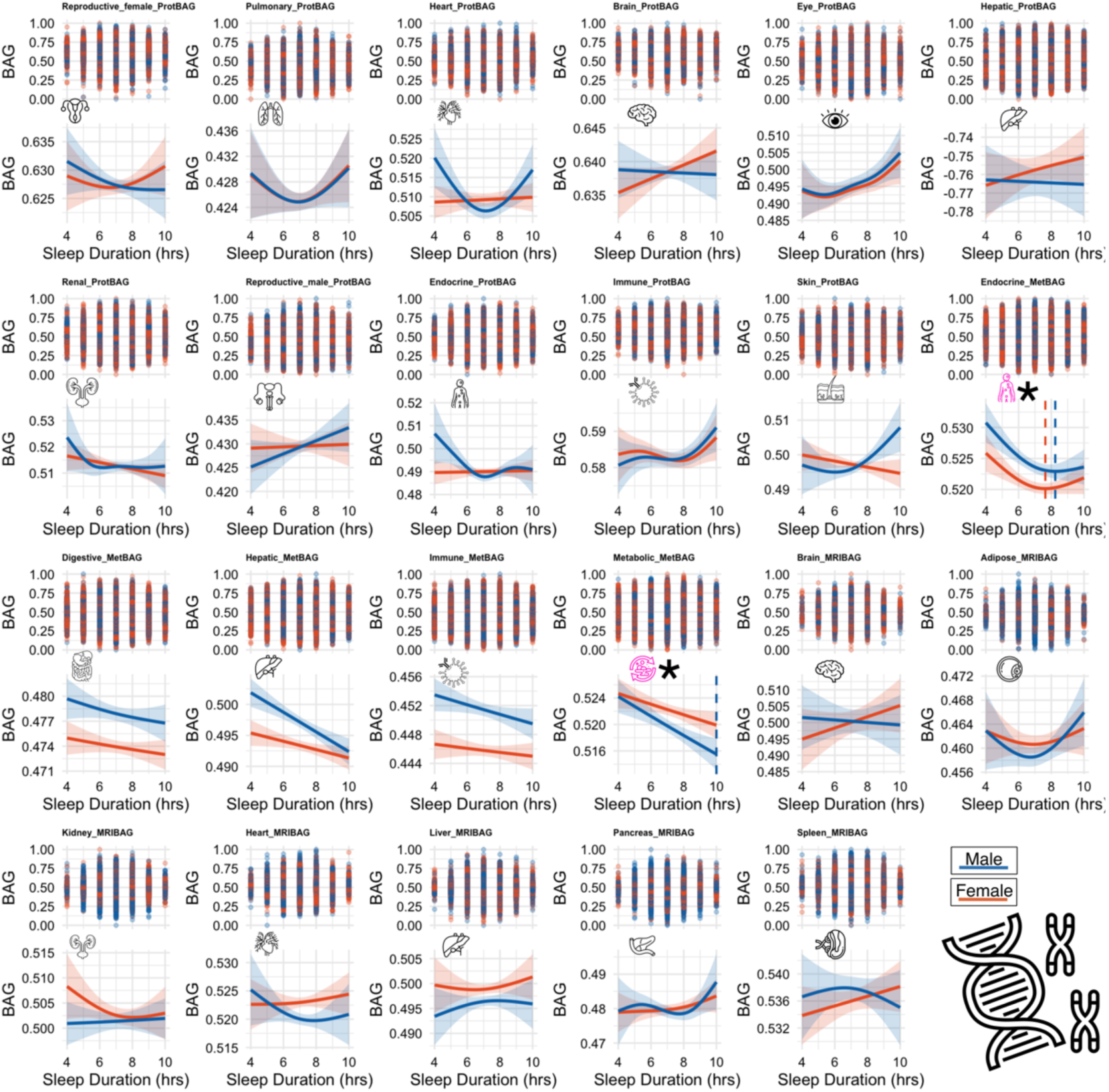
Associations between sleep duration and the polygenic risk score (PRS) of the 23 multi-organ, multi-omics BAGs. Unlike the BAGs, sleep duration (x-axis) did not exhibit a U-shaped relationship with the polygenic risk scores (PRS) of the 23 BAGs derived from common genetic variants (y-axis).

**Extended Data Figure 5:**
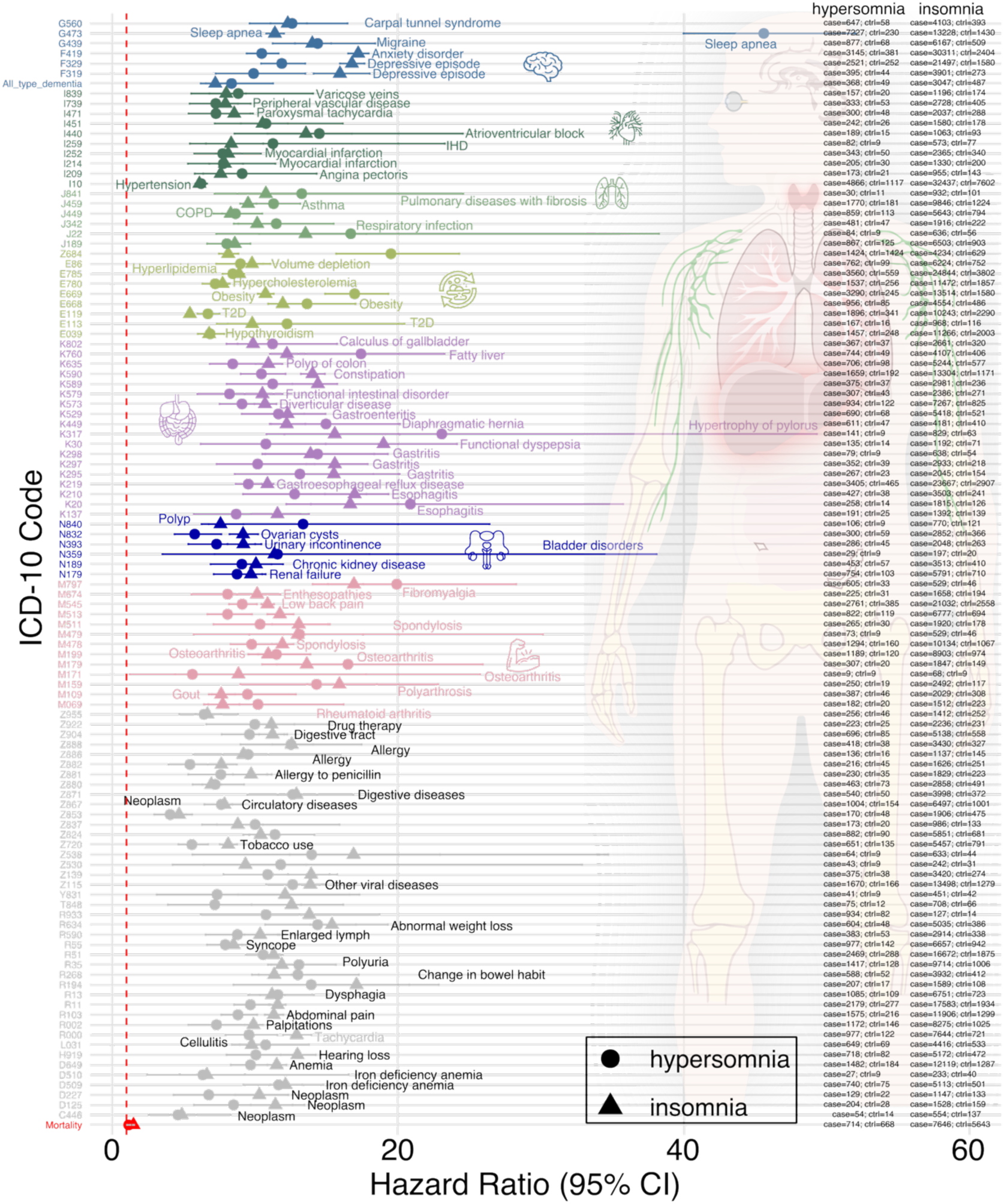
Insomnia and hypersomnia are associated with the time to incident of the 110 disease endpoints and mortality in the TriNetX database. Using insomnia and hypersomnia as proxies for short and long sleep duration, respectively, we replicated the significant associations shown in Fig. 5 within the UK Biobank cohort. Disease endpoints that remained significant after Bonferroni correction (0.05/111, accounting for 110 comorbidity-free disease endpoints and all-cause mortality) are indicated by colored shapes, while non-significant associations are shown in gray. One additional disease category was also tested for all types of dementia. Ctrl: health control.

**Extended Data Figure 6:**
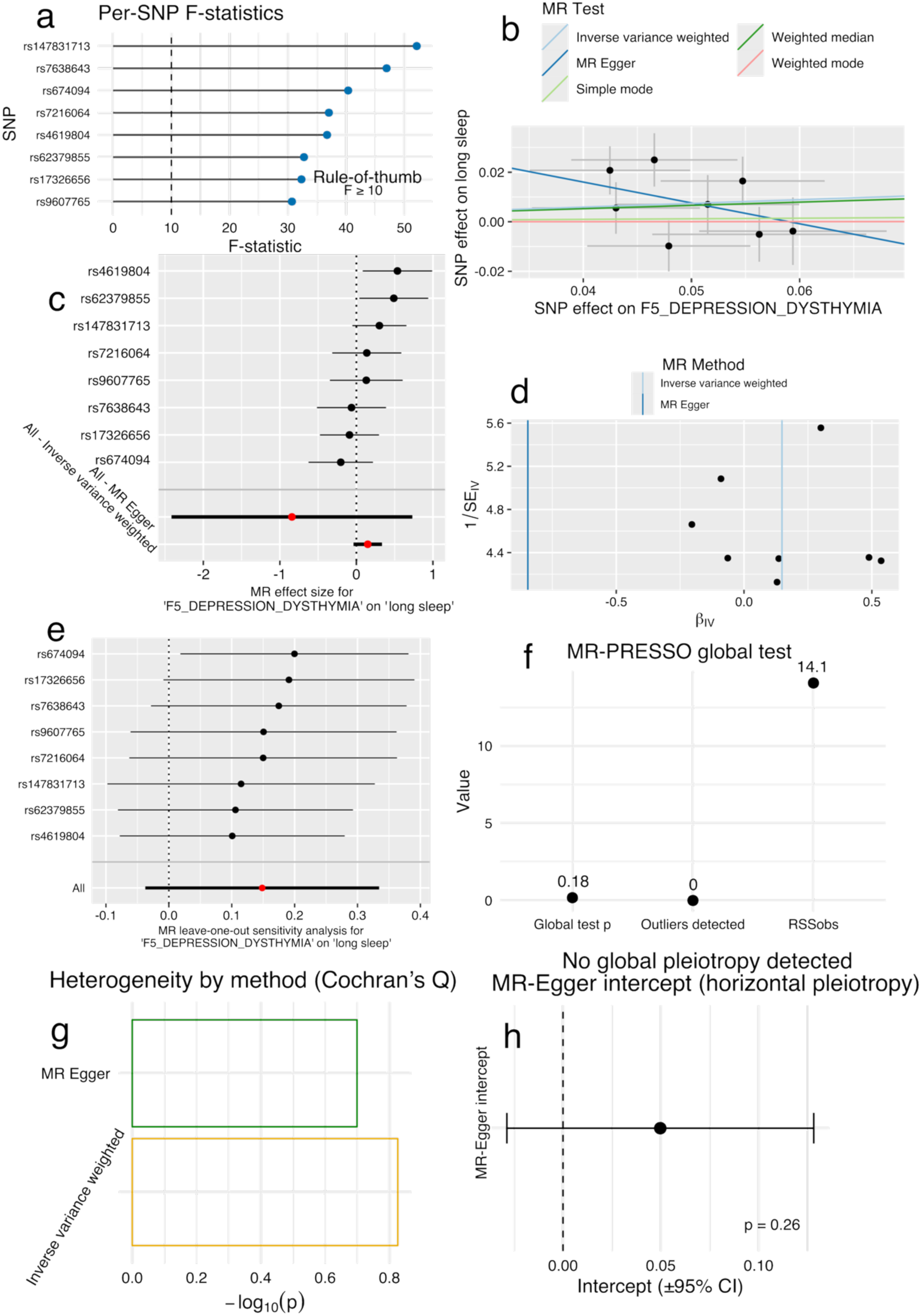
Sensitivity analysis for the causal relationship from FinnGen F5_DEPRESSION_DYSTHYMIA to long sleep duration. **a**) Lollipop plot of F-statistics for the 8 genome-wide significant depression instruments after preprocessing, considering linkage disequilibrium, ordered from weakest to strongest. All instruments exceed the commonly used rule-of-thumb threshold of F=10^110^ (min F=30.70; median F=36.88; mean F=38.62). The combined variance explained is R^2^-total = 0.00113, which corresponds to an approximate overall F-statistic of 38.66 (N=274,330; k=8), exceeding the conventional F>10 threshold that defines adequately strong instruments in univariable Mendelian randomization. **b**) Scatter plot for the Mendelian randomization effect sizes of the SNP-depression association (x-axis, log OR) and the SNP-long sleep duration associations (y-axis, log OR) with standard error bars. The slopes of the 5 lines correspond to the causal effect sizes estimated by the five MR estimators, respectively. For all five methods, no statistical significance was achieved (P-value>0.05). **c**) Forest plot for the single-SNP Mendelian randomization results. Each dot represents the effect (log OR), and the error bar displays the 95% CI using only one SNP; the red line shows the MR effect using all SNPs together for IVW and Egger estimators. **d**) Funnel plot for the relationship between the causal effect of depression on long sleep duration. Each dot represents causal effect sizes estimated using each SNP as a separate instrument against the inverse of the standard error of the causal estimate. **e**) Leave-one-SNP-out analysis of depression on long sleep duration. Each dot represents the causal effect (log OR), and the error bar displays the 95% CI by excluding that SNP from the analysis. The red line depicts the IVW estimator using all SNPs. **f**) MR-PRESSO confirms the absence of causality and a significant causal signal (P=0.16). Dots summarize the three key MR-PRESSO outputs: the global test p-value, the number of outlier instruments detected, and the observed residual sum of squares (RSSobs). Using eight SNPs, the global test was not significant (P-value=0.18), no outliers were identified (0), and RSSobs was 14.1, indicating no evidence of horizontal pleiotropy. MR-PRESSO was run with 1,000 simulations at *α*=0.05; because no outliers were detected, no outlier-corrected estimate was produced. **g**) Heterogeneity (Cochran’s Q) test indicated no strong between-instrument heterogeneity: MR-Egger Q=8.56 (df=6, p=0.200) and IVW Q=10.76 (df=7, P-value=0.149) using 8 SNPs. These P-values >0.05 suggest the causal estimates are not driven by substantial heterogeneity. **h**) The MR-Egger intercept was 0.0497 (SE=0.0400; P=0.260), indicating no evidence that directional (unbalanced) pleiotropy biases the causal estimate in this analysis.

**Extended Data Figure 7:**
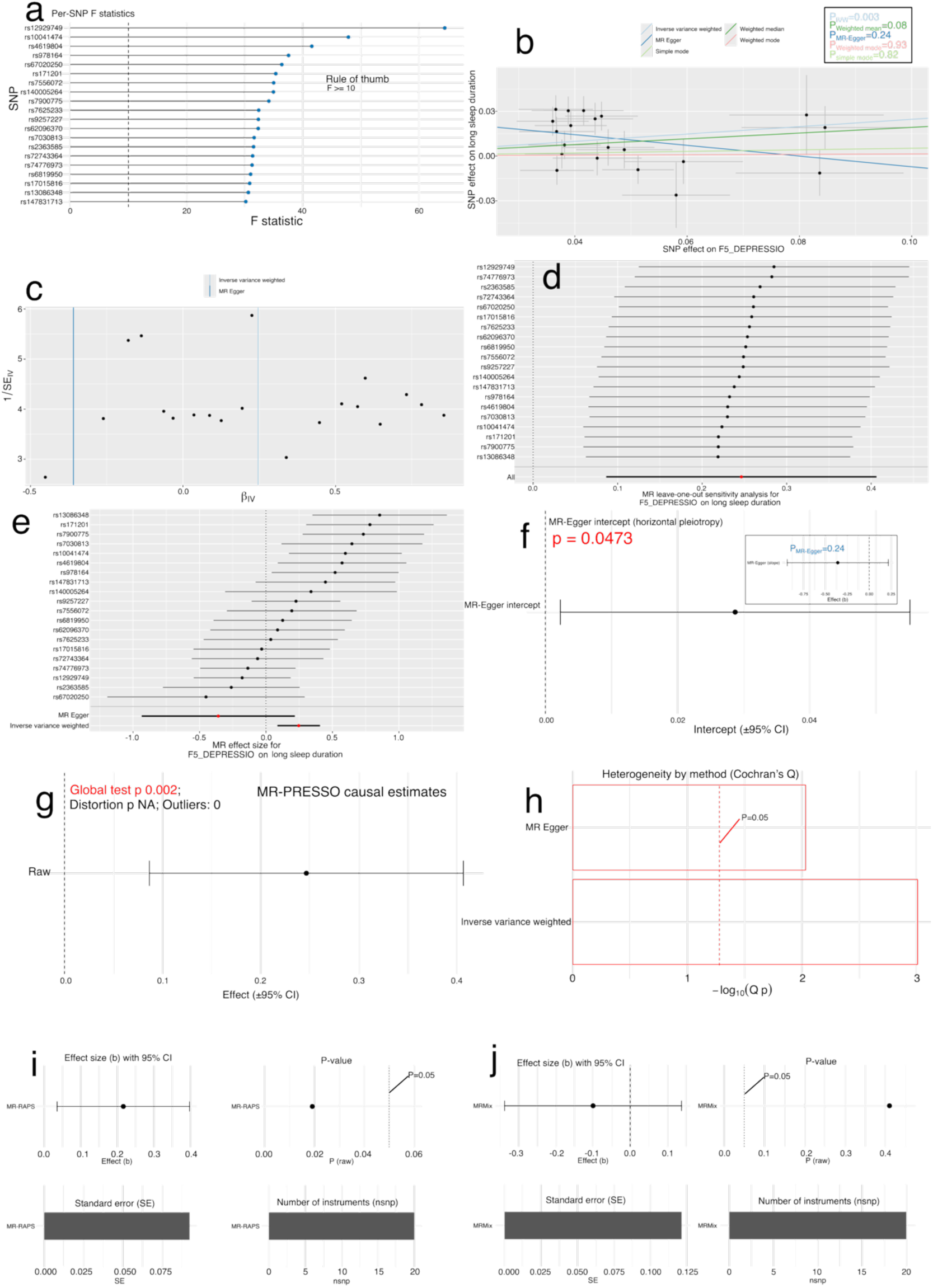
Sensitivity analysis for the causal relationship from the most recent MDD to long sleep duration in FinnGen. Mendelian randomization of major depressive disorder (MDD) on long sleep duration using the FinnGen (R12) GWAS. We downloaded the R12 FinnGen MDD GWAS with an expanded patient sample (*N*=59,333) to ensure adequate statistical power. **a)** Lollipop plot of first-stage F-statistics for the eight genome-wide significant MDD instruments after LD clumping, ordered from weakest to strongest. All instruments exceeded the conventional F > 10 threshold, suggesting sufficient instrument strength. **b)** Scatter plot of SNP-specific associations between MDD (x-axis, log OR) and long sleep duration (y-axis, log OR). The slopes of the five regression lines correspond to causal effect estimates from the inverse-variance weighted (IVW), weighted median, MR-Egger, simple-mode, and weighted-mode estimators. Among these, only the IVW method reached nominal significance (P=0.003). The MR-Egger slope was negative, contrasting with the positive slopes from all other estimators. **c)** Funnel plot showing the relationship between individual SNP causal effects and their precision, revealing heterogeneity across variants. **d)** Leave-one-SNP-out analysis of the MDD → long-sleep association; each point and 95 % CI represent the IVW estimate obtained when excluding a single SNP. **e)** Forest plot of single-SNP causal effects with corresponding 95 % confidence intervals. **f)** MR-Egger intercept plot showing a significant positive intercept (0.03 ± 0.01; P = 0.047), indicating directional pleiotropy that may bias the overall causal estimate. **g)** MR-PRESSO global test also detected horizontal pleiotropy (P = 0.002) but no outlier variants; consequently, the distortion test was not applicable. **h)** Cochran’s Q heterogeneity statistics for both IVW (P=0.0009) and MR-Egger (P=0.009) methods further confirmed substantial heterogeneity among the instruments. **i**) MR-RAPS models balanced (mean-zero) horizontal pleiotropy via an over-dispersion term and down-weights outliers using a robust loss, while profiling out weak-instrument bias. MR-RAPS point estimate remains positive but greatly attenuated versus IVW (P=0.019), indicating a signal that persists under balanced-pleiotropy/weak-IV adjustments, though with increased uncertainty. Interpretation should note that MR-RAPS does not correct for directional pleiotropy; results are therefore contingent on the balanced-pleiotropy assumption. **j**) MRMix estimates the causal effect by modeling SNP residuals as a two-component mixture, a spike for valid IVs (no direct effect) and a normal component with variance *σ²* for invalid/pleiotropic IVs. By probabilistically allocating pleiotropic SNPs to the high-variance component, MRMix limits their influence on the slope and is robust to a large fraction of invalid instruments when pleiotropic effects are mean-zero on average. Here, the estimate is near-null/slightly negative (P=0.41), with a CI spanning zero, providing no pleiotropy-robust evidence for a causal effect under the mixture model. To summarize, although IVW suggested a positive causal effect of genetic liability to MDD on long sleep (P = 0.003), multiple heterogeneity/pleiotropy diagnostics argue this is likely biased: the MR-Egger intercept is significant, the MR-PRESSO global test is positive, and Cochran’s Q indicates substantial heterogeneity. Pleiotropy-robust estimators do not corroborate the IVW signal: MR-Egger (slope) is null/negative (*β*=−0.36, P = 0.24), MRMix is near-null (*β* = −0.10, P = 0.41), and MR-RAPS, while largely attenuated and nominally positive (*β*=0.22, P = 0.019), remains sensitive to pleiotropy assumptions. Taken together, the data do not provide pleiotropy-robust evidence for a causal effect; any apparent association should be viewed as suggestive at best and interpreted with caution.

## The MULTI Consortium

Junhao Wen^1^, Cliodhna Kate O’Toole^1^, Christos Davatzikos^23^, Ye Ella Tian^3^, Andrew Zalesky^3^, Luigi Ferrucci^23^, Wenjia Bai^8^, Michael S. Rafii^12^, Paul Aisen^12^, Keenan A. Walker^7^

^1^Laboratory of AI and Biomedical Science (LABS), Columbia University, New York, NY, USA

^3^Melbourne Neuropsychiatry Centre, Department of Psychiatry, Melbourne Medical School, The University of Melbourne, Melbourne, Victoria, Australia

^7^Laboratory of Behavioral Neuroscience, National Institute on Aging, National Institutes of Health, Baltimore, MD, USA

^8^Department of Brain Sciences and Department of Computing, Imperial College London, London, UK

^12^Alzheimer’s Therapeutic Research Institute, Keck School of Medicine of the University of Southern California, San Diego, CA 92121, USA

^23^National Institute on Aging, National Institutes of Health, Baltimore, MD, USA

^24^Artificial Intelligence in Biomedical Imaging Laboratory (AIBIL), Center for AI and Data Science for Integrated Diagnostics (AI^2^D), Perelman School of Medicine, University of Pennsylvania, Philadelphia, PA, USA

