## supplement for "Sleep chart of biological aging clocks in middle and late life"

### Online Supplementary Materials

**Supplementary Note 1: U-shaped patterns between sleep duration and imaging-derived phenotypes, plasma proteins, and metabolites**
**Supplementary Note 2: The definition of genomic loci, independent significant SNP, lead SNP, candidate SNP**
**Supplementary Note 3: Comparisons between our sleep trait GWAS and genetic correlation results with previous literature**
**Supplementary Note 4: Sleep-DE signals identified in the UK Biobank in the TriNetX database**
**Supplementary Note 5: Association of binary traits of sleep duration as outcome variables in Mendelian randomization analyses with 525 disease endpoints in FinnGen and PGC**
**Supplementary Figure 1: Quantile-quantile (QQ) plots and residual vs. fitted value (RVF) plots for the 23 BAGs**
**Supplementary Figure 2: Split-sample analysis of the 23 BAGs**
**Supplementary Figure 3: Age-stratified analysis of the 23 BAGs**
**Supplementary Figure 4: Sex-stratified analysis of 23 BAGs**
**Supplementary Figure 5: Sensitivity check analysis by including age-sex interaction as an additional term**
**Supplementary Figure 6: Sensitivity check analysis by excluding participants with 3 sleep-related diseases**
**Supplementary Figure 7: Sensitivity check analysis by including smoking status as an additional covariate**
**Supplementary Figure 8: Sensitivity check analysis by including alcohol consumption status as an additional covariate**
**Supplementary Figure 9: Sensitivity check analysis by including Townsend deprivation index as an additional covariate**
**Supplementary Figure 10: Sensitivity check analysis by including computer use time as an additional covariate**
**Supplementary Figure 11: Sensitivity check analysis by testing the interaction of sleep duration and testosterone**
**Supplementary Figure 12: Sensitivity check analysis by testing the interaction of sleep duration and SHBG**
**Supplementary Figure 13: Sensitivity check analysis by testing the interaction of sleep duration and estradiol**
**Supplementary Figure 14: GAMLSS modeling of the 23 BAGs**
**Supplementary Figure 15: External validation of the U-shaped patterns using MESA self-reported sleep duration**
**Supplementary Figure 16: External validation of the U-shaped patterns based on BLSA actigraphy-derived sleep duration**
**Supplementary Figure 17: Sensitivity check of the ProWAS for protein FGFR2 of sleep duration mediated by the AST/ALT ratio**
**Supplementary Figure 18: QQ and Manhattan plots of the GWAS results**
**Supplementary Figure 19: Genetic correlation between our binary sleep GWAS with two previous GWASs from Dashti et al and Austin-Zimmerman et al**
**Supplementary Figure 20: Genetic correlation between FinnGen DEs and previous sleep GWASs from Dashti et al**

**Supplementary Figure 21: Genetic correlation between FinnGen DEs and previous sleep GWASs from Austin-Zimmerman et al**

**Supplementary Figure 22: Sensitivity analysis using 7–9 hours as the reference category for normal sleep duration in disease incidence prediction**

**Supplementary Table 1: The characteristics of participants consolidated via the MULTI study**

**Supplementary Table 2: Statistics of the GAM analysis between sleep duration and the 23 multi-organ, multi-omics biological aging clocks**

**Supplementary Table 3: Sensitivity check analyses between sleep duration and the 23 BAGs**

**Supplementary Table 4: Genomic loci identified in our GWAS of the abnormal sleep duration patterns**

**Supplementary Table 5: Genetic correlation between short/long sleep duration and 23 BAGs**

**Supplementary Table 6: Sensitivity analysis for our mediation analysis using brain MRI as the mediator**

**Supplementary Table 7: 6 PGC GWAS summary statistics**

**Supplementary Table 8: Baseline characteristics of the TriNetX database**

**Supplementary Note 1: U-shaped patterns between sleep duration and imaging-derived phenotypes, plasma proteins, and metabolites**

**Imaging-derived phenotypes from 8 organ systems and tissues show U-shaped relationships with sleep duration**

Using *in vivo* imaging-derived phenotypes (IDPs) derived from MRI data of 8 organs and tissues, we provided a granular understanding of how sleep duration impacts specific structural and functional aspects of various organs (**Extended Data Figure 1** and **Method 4**).

We identified a total of 162 statistically significant associations ($P<0.05/720$) between sleep duration and IDPs (**Extended Data Figure 1a**). Among the 27 brain IDPs derived from T1-weighted MRI (i.e., brain-GM) and diffusion MRI (i.e., brain-WM), nonlinear relationships with sleep duration were observed across brain regions (**Extended Data Figure 1b**), including the frontal, temporal, and parietal lobes, as well as subcortical structures, with the left hippocampus showing pronounced association (EDF=1.96; $P_1<1\times10^{-20}$, $P_2<1\times10^{-20}$, $P_3=1$; **Extended Data Figure 1d**). Among the 10 diffusion MRI-derived IDPs (i.e., brain-WM; **Extended Data Figure 1c** for fractional anisotropy), exemplified by the right superior cerebellar peduncle (EDF=1.97; $P_1<1\times10^{-20}$, $P_2<1\times10^{-20}$; $P_3<1\times10^{-20}$; **Extended Data Figure 1e**). Among the 54 resting-state fMRI-derived imaging phenotypes, the 189th functional connectivity measure (FullCorr_189) between two brain networks exhibited a U-shaped association with sleep duration (EDF=3.48; $P_1<1\times10^{-20}$, $P_2<1\times10^{-20}$; $P_3=1$; **Extended Data Figure 1f**).

Beyond the brain, we observed 51 significant associations, exemplified by the INL-ELM thickness of the central subfield in the left eye (EDF=3.42; $P_1<1\times10^{-20}$, $P_2<1\times10^{-20}$; $P_3=0.18$; **Extended Data Figure 1g**). Among the 10 significant heart IDPs, the largest volume of the right atrium of the heart showed a significant U-shaped pattern with sleep duration (EDF=1.97; $P_1<1\times10^{-20}$, $P_2<1\times10^{-20}$; $P_3=0.93$; **Extended Data Figure 1h**). For the rest of the abdominal IDPs, U-shaped patterns were observed in abdominal fat ratio (EDF=3.30; $P_1=3.0\times10^{-5}$, $P_2<1\times10^{-20}$; $P_3=0.03$; **Extended Data Figure 1i**) and corrected T1 (cT1) of the liver (EDF=1.96; $P_1<1\times10^{-20}$, $P_2=3.94\times10^{-3}$; $P_3=0.97$; **Extended Data Figure 1j**). Detailed statistics, including the P value, optimal family distribution, and EDF, are presented in **Supplementary File 2**.

We found widespread nonlinear associations between sleep duration and structural and functional imaging phenotypes across 8 organs. These organ-level changes point to systemic effects of abnormal sleep duration, leading us to examine corresponding molecular signatures in proteomic and metabolomic profiles.

**Sleep duration and plasma proteomics show a U-shaped relationship**

We analyzed associations between sleep duration and 2923 plasma proteins using the same GAM model. When considering only the 342 organ-enriched proteins (i.e., at least four-fold higher mRNA level in brain compared to any other tissues) defined by our previous study[1], we identified 77 significant proteins ($P < 0.05/2923$) (**Method 5**).

We identified 29, 14, 9, and 8 significant organ-enriched proteins corresponding to the hepatic, immune system, endocrine system, and brain, respectively (**Extended Data Figure 2a**). For instance, the brain-enriched protein FGFR2 demonstrated a U-shaped relationship with sleep duration (EDF=3.64; $P_1<1\times10^{-20}$, $P_2<1\times10^{-20}$; $P_3=0.03$; **Extended Data Figure 2b**). We performed protein-protein interaction (PPI) analysis using STRING (v12.0: https://string-db.org/) on the significant organ-enriched proteins to explore functional relationships among the 14 immune-enriched and 29 hepatic-enriched proteins. For the immune protein set, we found a cluster of immune-enriched proteins that likely reflects a coordinated immune response involving cell surface receptors, leukocyte activation, and inflammatory signaling (CD27, FCER2, ITGAM, NCR1, PLAUR, PRTN3, RETN; **Extended Data Figure 2c**). For the hepatic protein set, k-means clustering of the PPI network revealed a prominent cluster consisting of 19 proteins involved in innate immune defense and blood coagulation, particularly complement components (e.g., C3, C4A, CFB) and coagulation factors (e.g., FGA, FGB, FGG), highlighting the role of complement and coagulation cascades (**Extended Data Figure 2d**).

Our protein set enrichment analysis (PSEA), based on Gene Ontology (GO) biological process terms for the significant immune- and hepatic-enriched proteins, identified a coherent set of pathways that align with well-established hypotheses connecting sleep duration to systemic physiological regulation and disease susceptibility. Notably, a substantial proportion of enriched pathways mapped to immune activation and inflammation, including "defense response" (GO:0006952), "innate immune response" (GO:0045087), "complement activation" (GO:0006956), and "neutrophil chemotaxis" (GO:0030593), supporting prior evidence that sleep disruption can modulate both innate and adaptive immune pathways. Additionally, we observed enrichment in metabolic regulation and protein processing, including "regulation of catalytic activity" (GO:0050790), "enzyme inhibitor activity" (GO:0004857), extracellular signaling and vesicle transport (e.g., "extracellular exosome" (GO:0070062), "blood microparticle" (GO:0072562)), and cell surface and receptor signaling, reflecting the role of sleep in coordinating cellular communication, metabolic homeostasis, and systemic signaling. Enrichment in generalized response to stimulus and stress-related pathways further underscores the broad physiological perturbations linked to altered sleep duration. Collectively, these findings provide mechanistic support for the hypothesis that sleep impacts multi-organ health through diverse but interconnected biological systems. Detailed statistics, including the P value, optimal family distribution, and EDF, are presented in **Supplementary File 3**.

**Sleep duration and plasma metabolomics show a U-shaped relationship**

We further investigated the relationship between sleep duration and 327 NMR-derived metabolomic measures, which included 107 raw small-molecule metabolites and lipid subclasses, along with additional composite metrics derived from these metabolites. Among the 107 organ-associated metabolites identified in our previous study[2], we observed 21 significant associations after Bonferroni correction ($P<0.05/327$) (**Method 6**).

We identified 14 significant organ-associated metabolites linked to the hepatic system, 4 to the digestive system, and 2 to the endocrine system (**Extended Data Figure 3a**). For instance, the average number of double bonds in fatty acid chains within lipid molecules ('Unsaturation') demonstrated a U-shaped relationship with sleep duration (EDF=3.75; $P_1<1\times10^{-20}$, $P_2<1\times10^{-20}$; $P_3=0.003$; **Extended Data Figure 3b**). Similarly, the U-shaped relationship was also observed in acetoacetate (EDF=1.98; $P_1<1\times10^{-20}$, $P_2=1$; $P_3=0.46$), docosahexaenoic acid (DHA) (EDF=3.23; $P_1<1\times10^{-20}$, $P_2=1$, $P_3<1\times10^{-20}$), and acetate (EDF=1.97; $P_1<1\times10^{-20}$, $P_2<1\times10^{-20}$; $P_3=0.69$) (**Extended Data Figure 3c-e**).

Our metabolite set enrichment analysis (MSEA) using the MetaboAnalyst (v6.0: https://www.metaboanalyst.ca/MetaboAnalyst/ModuleView.xhtml), including only the small-molecule metabolites that were mapped to the platform (e.g., HMDB and PubChem ID). This resulted in acetic acid, acetoacetic acid, creatinine, dehydroepiandrosterone, glycine, L-Tyrosine, L-Valine, and sphingomyelins. We identified a coherent grouping of pathways that support potential hypotheses linking sleep duration to systemic physiology (**Extended Data Figure 3f**). Pathways within the "Transport Axis" cluster highlight the involvement of solute carrier (SLC) transporters, which regulate the movement of amino acids, neurotransmitters, and ions across cellular membranes. This cluster includes both generic and disorder-specific pathways, such as SLC-mediated transmembrane transport, amino acid transport across the plasma membrane, and Na/Cl-dependent neurotransmitter transporters, underscoring a possible role for sleep in modulating nutrient and neurotransmitter bioavailability across tissue barriers. The "Neurochemical Plasticity" cluster reflects the role of sleep in maintaining neural homeostasis through amino acid metabolism and synaptic remodeling. Enriched pathways such as tyrosine catabolism, creatine metabolism, and various tRNA aminoacylation processes suggest that sleep duration may influence neurotransmitter synthesis, protein translation fidelity, and post-translational modifications critical for brain function. Finally, the "Liver-Sleep Axis" cluster includes pathways like synthesis of ketone bodies and cyclosporin A-induced metabolic pathways, pointing toward the hepatic regulation of energy metabolism and detoxification as potential downstream targets of sleep-driven metabolic adaptation. Together, these findings further support a multi-system view of sleep biology involving molecular transport, neurochemical processing, and liver-mediated metabolic control. Detailed statistics, including the P value, optimal family distribution, and EDF, are presented in **Supplementary File 4.**

**Supplementary Note 2: The definition of genomic loci, independent significant SNP, lead SNP, candidate SNP**

FUMA defined the significant independent SNPs, lead SNPs, candidate SNPs, and genomic risk loci as follows (https://fuma.ctglab.nl/tutorial#snp2gene):

*Independent significant SNPs*

They are defined as SNPs with $P \leq 5\times10^{-8}$ that are independent of each other at the user-defined $r^2$ (set to 0.6 in the current study). We further describe *candidate SNPs* as those in linkage disequilibrium (LD) with independent significant SNPs. FUMA then queries each candidate SNP in the GWAS Catalog to check whether any clinical traits have been reported to be associated with previous GWAS studies.

*Lead SNPs*

Lead SNPs are defined as independent significant SNPs that are also independent of each other at $r^2<0.1$. If multiple independent significant SNPs are correlated at $r^2\geq0.1$, then the one with the lowest individual *P*-value becomes the lead SNP. If $r^2$ threshold is set to 0.1 for the independent significant SNPs, then they would constitute the identical set as the lead SNPs. FUMA thus advises setting $r^2$ to be 0.6 or higher.

*Genomic risk loci*

FUMA defines genomic risk loci to include all independent signals physically close or overlapping in a single locus. First, independent significant SNPs dependent on each other at $r^2\geq0.1$ are assigned to the same genomic risk locus. Then, independent significant SNPs with less than the user-defined distance (250 kilobases by default) away from one another are merged into the same genomic risk locus – the distance between two LD blocks of two independent significant SNPs is the distance between the closest points from each LD block. Each locus is represented by the SNP within the locus with the lowest *P*-value.

**Supplementary Note 3: Comparisons between our sleep trait GWAS and genetic correlation results with previous literature**

We re-ran GWAS for short and long sleep duration in the UK Biobank for both practical and methodological reasons. Practically, this allowed us to apply a unified quality-control pipeline and to obtain harmonized information on allele frequencies and linkage disequilibrium, which are not always fully available in previously published summary statistics. Methodologically, our design choices, defining short, normal, and long sleep using specific bins (short <6 h, normal 6–8 h, long >8 h), restricting the analysis to individuals reporting 4-10 h of sleep to reduce the influence of extreme outliers, and treating sleep duration consistently as a categorical phenotype in downstream models, were aligned with the aims and structure of our multi-organ analyses.

These choices differ from those in Dashti et al.[3], who primarily modeled sleep duration as a continuous trait and used different categorical thresholds for short vs. normal and long vs. normal sleep duration (≤6h, 7–8 h, ≥9 h), and from Austin-Zimmerman et al.[4], who combined UK Biobank with the MVP cohort and used alternate cutoffs (≤5 h, 7–8 h, ≥10 h). Dashti et al excluded extreme responses of less than 3 h or more than 18 h, which is much less strict than ours. Consequently, their studies achieved larger effective sample sizes and identified more genome-wide significant loci.

To directly compare these efforts, we downloaded GWAS summary statistics from Dashti et al. and Austin-Zimmerman et al. and estimated pairwise genetic correlations between their GWAS and our own short vs. normal and long vs. normal sleep GWAS using LDSC. We observed very high genetic correlations ($0.83 < r_g < 0.98$; P-value<$1x10^{-5}$; **Supplementary Figure 19**), indicating that the underlying genetic architecture is highly similar and that differences in the number of genome-wide significant loci are largely attributable to differences in sample size, power, and operational definitions of short, normal, and long sleep duration rather than to fundamentally different biology.

Genetic correlations between sleep traits and other complex traits have been reported in multiple previous studies[3,4,5,6,7]. However, these prior analyses generally follow a candidate or hypothesis-driven strategy, focusing on selected domains (such as psychiatric or cardiometabolic traits) and typically pairing each trait with the single most highly powered GWAS available. In contrast, our work leverages large biobank-scale resources (in particular for FinnGen) in a non-selective, data-driven manner and is explicitly organized around a multi-organ perspective, linking short and long sleep duration to disease endpoints across primary organ systems (brain, heart, lung, metabolic organs, etc.). We provided additional analyses by re-performing genetic correlation between the FinnGen DEs with the GWAS summary statistics from Dashti et al.[3] and Austin-Zimmerman et al.[4] (**Supplementary Figure 20-21**), re-confirming the multi-organ dysfunction relationship with both short and long sleep duration. Overall, the DE–sleep genetic associations identified in our GWAS showed strong concordance with those from the two previous studies. However, the latter generally detected a larger number of significant genetic correlation signals, likely reflecting their larger sample sizes and differing definitions of short, normal, and long sleep duration. Thus, our re-analysis of sleep GWAS and the accompanying LDSC framework are not intended to supersede prior sleep GWAS per se, but to provide a harmonized, biobank-based foundation for systematically mapping how sleep disturbances are genetically connected to diseases spanning the whole body.

**Supplementary Note 4: Sleep-DE signals identified in the UK Biobank in the TriNetX database**

**Data**: We used EHR data from the TriNetX database to validate the survival analysis of the 153 significant disease endpoints (DEs) associated with short and long sleep durations identified in the UK Biobank (UKBB). Since sleep duration is not directly available in TriNetX, we used diagnoses of insomnia and hypersomnia as proxy measures. Baseline characteristics were compared with chi-square tests for categorical variables and independent-sample t-tests for continuous variables (**Supplementary Table 6**). This active comparator cohort study used data from commercially insured adults (aged ≥18 years), with insomnia or hypersomnia diagnoses and controls without insomnia or hypersomnia, respectively, between January 1st, 2010, and August 31st, 2017. Patients were included if they had undergone a healthcare visit within one year before their cohort entry. Patients were excluded if they had any DE diagnosis at any time before cohort entry. This analysis was conducted on 6th May 2025 on the TriNetX Analytics Platform.

**Method**: The TriNetX platform was used to run 1:1 propensity score matching using logistic regression. Once it yields scores that range between 0 and 1, the platform utilizes a greedy, nearest neighbour matching with a calliper of 0.1 pooled standard deviations once propensity scores have been calculated for each patient. Kaplan–Meier analysis was used to estimate the probability of outcome at daily time intervals with censoring applied. When the last fact (outcomes of interest or other medical encounters) in the patient's record was in the time window for analysis, the patient was censored on the day after the last fact in their record. We adjusted for the following covariates: age, sex, ethnicity, race, cerebrovascular diseases, infectious diseases, neoplasms, outpatient visits, hospital inpatient visits, beta blockers, ace inhibitors, calcium channel blockers, angiotensin blockers, metformin, glucagon like peptide 1 agonist, body mass index, total cholesterol, LDL-C, HDL-C, and HbA1c. We defined participants with insomnia at baseline who were diagnosed later for the DE of interest (e.g., I10 for essential (primary) as cases and participants without insomnia at baseline who were diagnosed later for the DE as controls. The Cox proportional hazards model was employed to derive the adjusted hazard ratios (HRs). HRs and 95% confidence intervals (CIs) were used to describe the relative hazard of the outcomes based on a comparison of time-to-event rates. The proportional hazards assumption was tested using the generalized Schoenfeld approach on the TriNetX platform, with adjusted HRs recalculated for specific time intervals if the assumptions were violated.

**Compared to UKBB**: In the TriNetX database, after propensity score matching, there were 110,374 participants with insomnia, and 16,371 participants with hypersomnia. We identified 109 overlapping DEs, along with data on all-cause mortality and an additional disease category encompassing all types of dementia. Using a Bonferroni correction threshold (0.05/111), we found that 110 signals were replicated for insomnia, and 97 for hypersomnia (**Extended Data Figure 4 and Supplementary File 7**).

**Insomnia**: In TriNetX, our replication of the UKBB results for insomnia yielded consistent hazard ratios (all >1), reinforcing the robustness of the associations with short sleep duration noted in UKBB. Key endpoints such as hypertension (I10), hypercholesterolemia (E780), diverticular disease (K573), diaphragmatic hernia (K449), asthma (J459), gastroesophageal reflux disease (K219), type 2 diabetes (E119), circulatory conditions (Z867), and osteoarthritis (M179) all demonstrated hazard ratios in the range of ~6–17 in TriNetX, well above the ~1.5–2.8 range observed in UKBB. It may be noted here that these HRs across datasets are not directly comparable due to different reference populations. This pattern held across numerous endpoints, including COPD, obesity, depressive episodes, and musculoskeletal disorders, with especially high replication signals (e.g., gastritis [K297 HR ~15.6] and anemia [D649 HR ~11.5]). Importantly, we also confirmed replication for all-cause mortality (HR ~1.49) and, available in TriNetX, all-types of dementia, which showed a strong association (HR ~7.2), further validating the link between sleep disturbance and neurodegeneration. Overall, the TriNetX evidence not only supports the UKBB findings but also amplifies the magnitude of risk, particularly for neurological and chronic conditions, highlighting the profound health impact of insomnia-associated DEs.

**Hypersomnia**: The TriNetX replication analysis of hypersomnia (P<0.05/111) identified significant associations with 97 DEs, mortality, and all-type dementia, demonstrating remarkably consistent directionality with the UKBB's long sleep duration findings despite different methodologies. The strongest replicated associations included metabolic disorders (obesity: HR=16.96), psychiatric conditions (depression: HR=11.8; anxiety: HR=10.46), and gastrointestinal diseases (GERD: HR=9.52), with particularly robust effects for sleep apnea (HR=45.59). Notably, effect sizes in TriNetX were substantially larger than UK Biobank (e.g., 3.9-fold higher HR for obesity), likely reflecting the more extreme phenotype of clinically diagnosed hypersomnia versus self-reported long sleep. Both datasets converged on elevated risks for cardiometabolic, neuropsychiatric, and digestive disorders, though TriNetX revealed stronger associations with acute conditions (e.g., pulmonary fibrosis: HR=13.26) and medical interventions (chemotherapy: HR=7.20), potentially due to its electronic health record-based population with greater disease severity. The risk of dementia (HR=8.35) and replication of mortality (HR=1.15) across cohorts underscores the clinical significance of prolonged sleep phenotypes.

**Supplementary Note 5: Association of binary traits of sleep duration as outcome variables in Mendelian randomization analyses with 525 disease endpoints in FinnGen and PGC**

In our previous study[8], two-sample Mendelian randomization suggested that sleep duration may arise as a consequence (outcome) of brain aging (exposure). Importantly, that analysis was based on a sleep duration GWAS using a linear model (via Plink), which did not account for the nonlinear associations identified in the current work. Given the relatively underpowered GWAS for the LLD subtypes, we could not directly test the inverse causality from the 2 LLD subtypes to the 2 sleep duration binary traits.

To further explore the possibility of reverse causality, specifically, whether sleep duration patterns may reflect consequences of underlying disease burden, we performed additional two-sample Mendelian randomization analyses using the binary sleep duration traits (short vs. normal and long vs. normal) as outcome variables, and 525 disease endpoints (DEs) from FinnGen and PGC as exposure variables (**Supplementary File 9**). Among the 179 DEs with sufficient instrumental variables (>7 IVs), we found no widespread evidence of causal effects on sleep duration. The only association that remained statistically significant after Bonferroni correction was from obesity (FinnGen code: E4_OBESITYCAL) to long sleep duration, with a P-value of $3.00\times10^{-6}$ and an odds ratio of 0.80 (95% CI: 0.74–0.88) based on 10 SNPs. This suggests a potential inverse relationship, where genetic liability to obesity is associated with a lower likelihood of long sleep duration.

Furthermore, our sequential equation modeling results support a mediating role of organ-specific BAGs in linking sleep patterns to neuropsychiatric outcomes, suggesting a directional pathway (not strictly causal) where sleep may influence systemic aging processes that in turn modulate disease risk. Taken together, although we cannot fully rule out the possibility of reverse causality, especially for certain traits such as obesity, our current multi-modal evidence, integrating GWAS, MR, genetic correlation, and SEM, leans toward the interpretation that sleep duration is more likely a modifiable factor rather than a downstream consequence of disease burden. This perspective reinforces the potential of sleep-targeted interventions in disease prevention and aging modification strategies.

**Supplementary Figure 1: Quantile-quantile (QQ) plots and residual vs. fitted value (RVF) plots for the 23 BAGs**

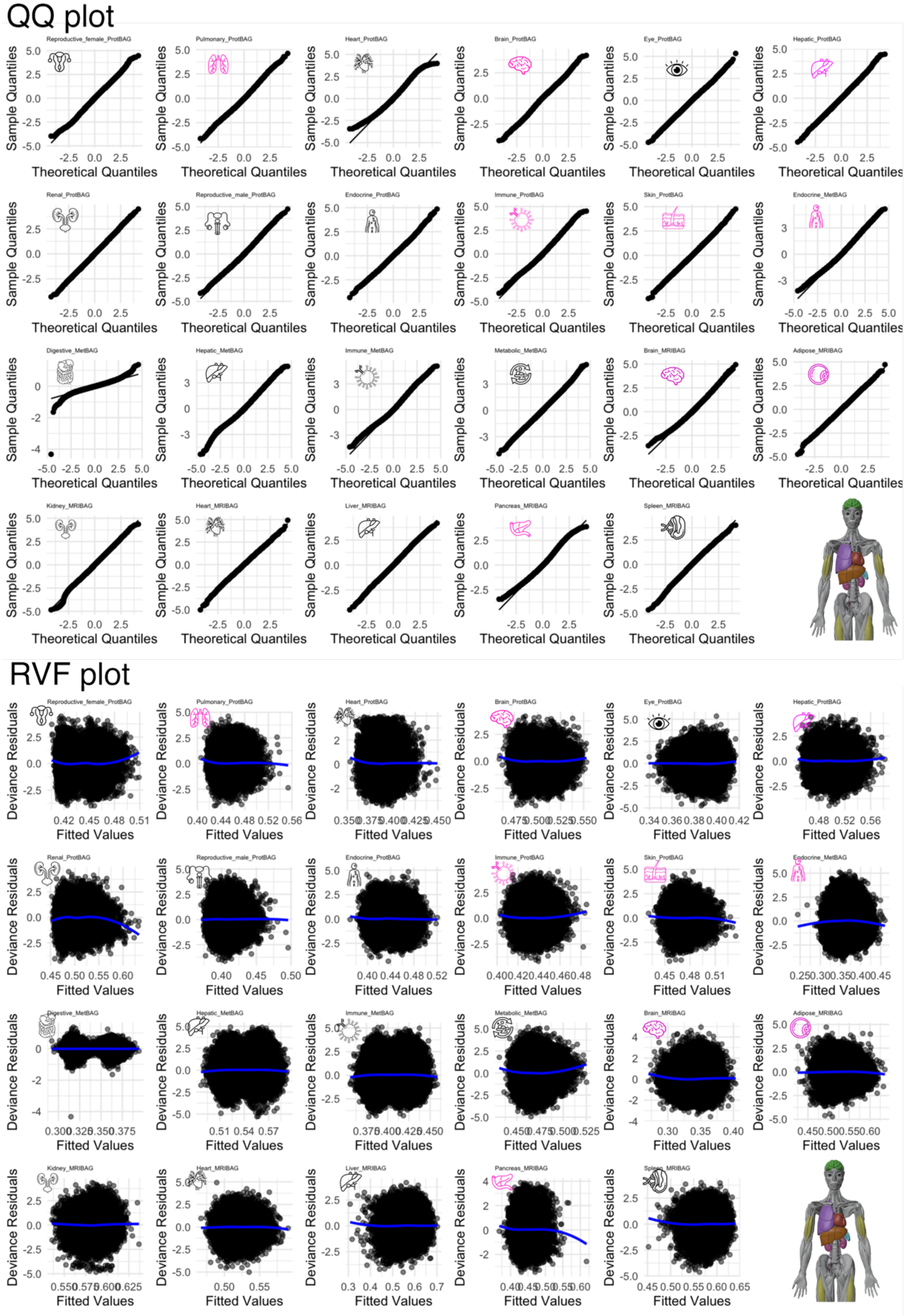


For the optimal model, defined by the selected smoothing parameter $k$ and appropriate distribution family, we present the QQ plots and residual vs. fitted value (RVF) plots for each

BAG. The QQ plots demonstrated that the residuals from these significant models generally followed the theoretical normal distribution, with only minor deviations in the tails. Similarly, the residual-vs-fitted (RVF) plots for the significant associations exhibited no major heteroscedasticity, with residuals evenly scattered around zero and smooth loess lines remaining largely flat. These patterns indicate stable variance, and a good model fit across the range of predicted values. Importantly, the significant associations did not display stronger deviations or assumption violations compared to non-significant models. Together, these diagnostics support that the reported significant sleep–BAG associations are unlikely to be artifacts of model misspecification or assumption violations.

**Supplementary Figure 2: Split-sample analysis of the 23 BAGs**

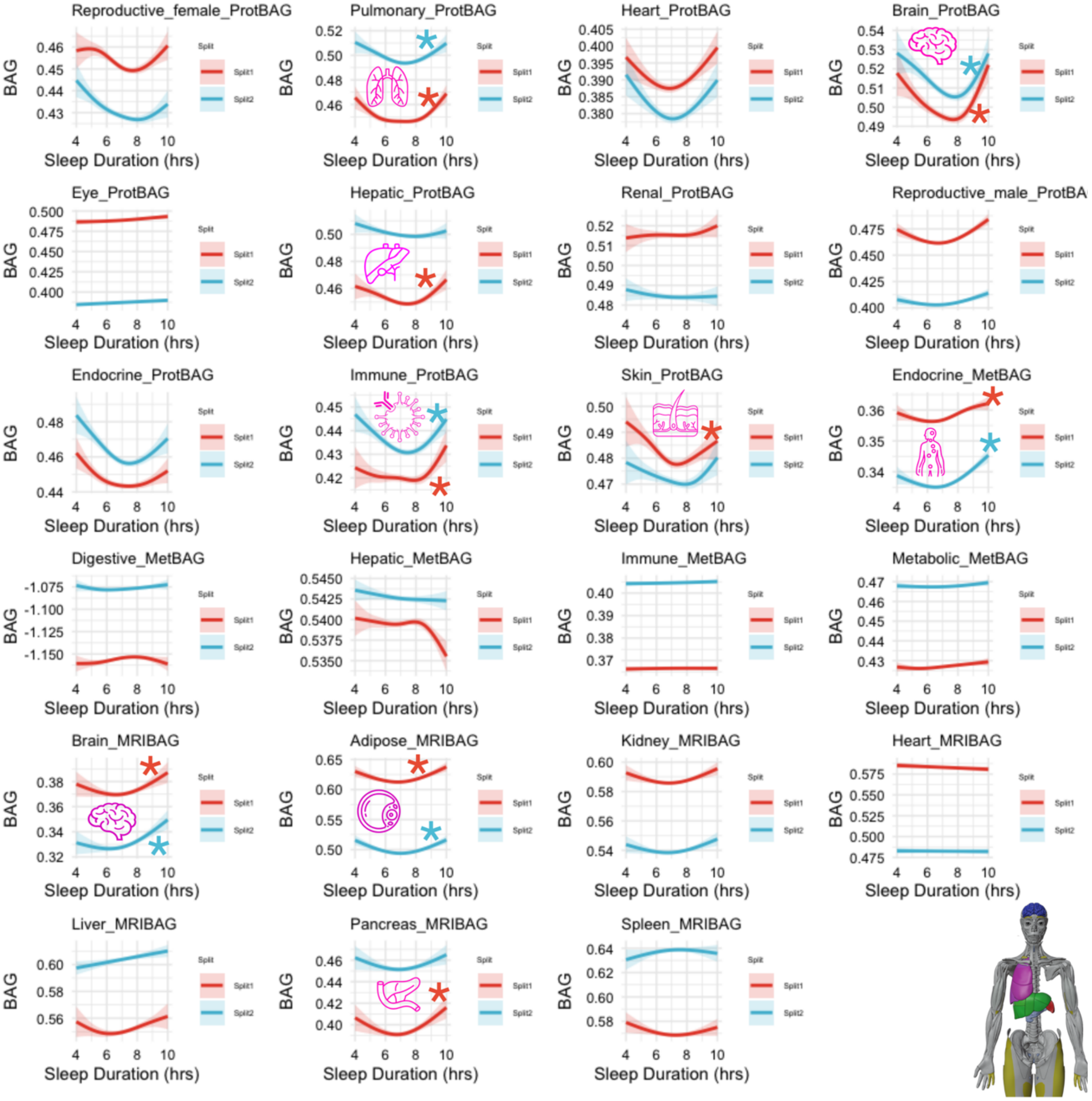


We randomly divided the entire population into split1 and split2 populations, then refitted the same GAM model (i.e., the optimal *k* and family distribution) to evaluate the consistency of the detected U-shaped patterns. Due to reduced sample size, we defined the P-value threshold here as P<0.05/9; we used the start symbol (*) to indicate significant signals. Overall, we found a robust U-shaped pattern across random splits. Reduced statistical power may explain the lack of significant findings in some BAGs. We showed the modeling results for all 23 BAGs for transparency.

**Supplementary Figure 3: Age-stratified analysis of the 23 BAGs**

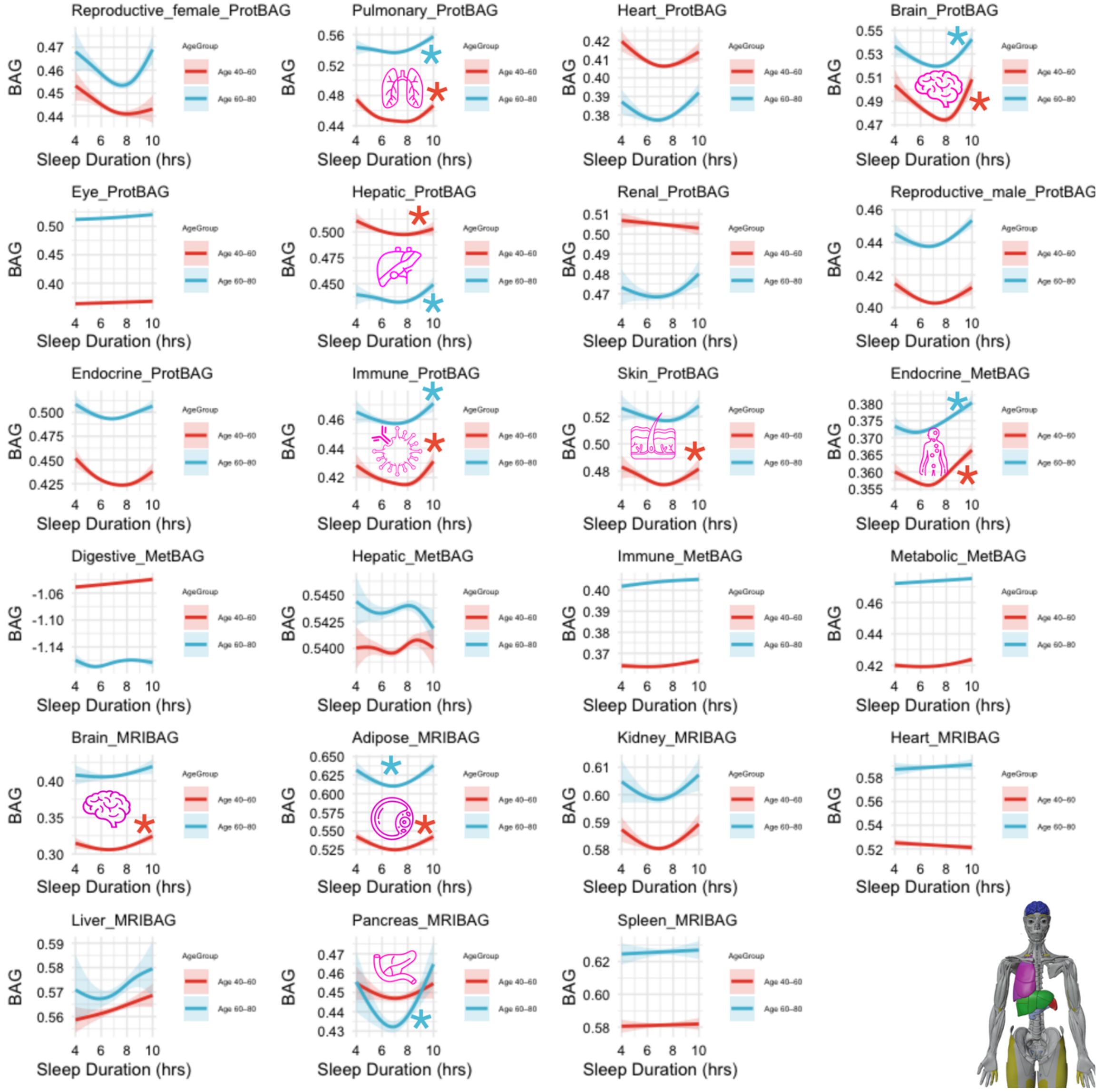


We stratified the entire population into [40-60] years old and [60-80] years old bins to evaluate the robustness of the U-shaped patterns. Due to reduced sample size, we defined the P-value threshold here as P<0.05/9; we used the start symbol (*) to indicate significant signals. Overall, we found a robust U-shaped pattern across age strata. Age-stratified analyses largely reproduced the main associations, with only non-significant effects observed in the 60–80-year group for skin ProtBAG and brain MRIBAG, and in the 40–60-year group for pancreas MRIBAG. Reduced statistical power and a narrower age range within strata may explain the lack of significant findings in these age groups. We showed the modeling results for all 23 BAGs for transparency.

**Supplementary Figure 4: Sex-stratified analysis of 23 BAGs**

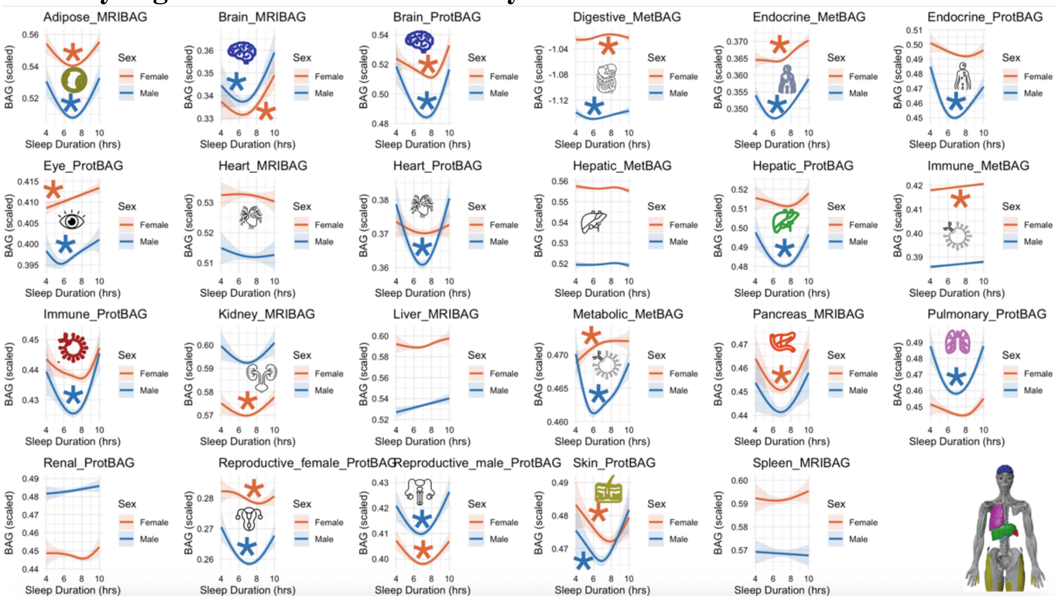


We split the cohort into females and males and re-fit the same GAM specification selected in the main analysis (fixed optimal k and family), testing the sleep term within each sex. Significance was assessed with Bonferroni correction ($\alpha = 0.05/23$); significant panels are marked with icons and asterisks. Overall, U-shaped associations were broadly reproducible across sexes, with some BAGs showing stronger effects in males (e.g., the endocrine ProtBAG) and others in females (e.g., pancreas and kidney MRIBAG). These sex differences may reflect biology, such as hormone milieu (estrogen/androgen signaling), sex-specific sleep architecture and circadian regulation, organ-specific metabolic and inflammatory pathways, and life-stage effects (e.g., menopause), in addition to possible sample-size or residual confounding differences between strata. Colored icons represent the 9 significant sleep-BAG signals identified in **Fig. 1**.

**Supplementary Figure 5: Sensitivity check analysis by including age-sex interaction as an additional term**

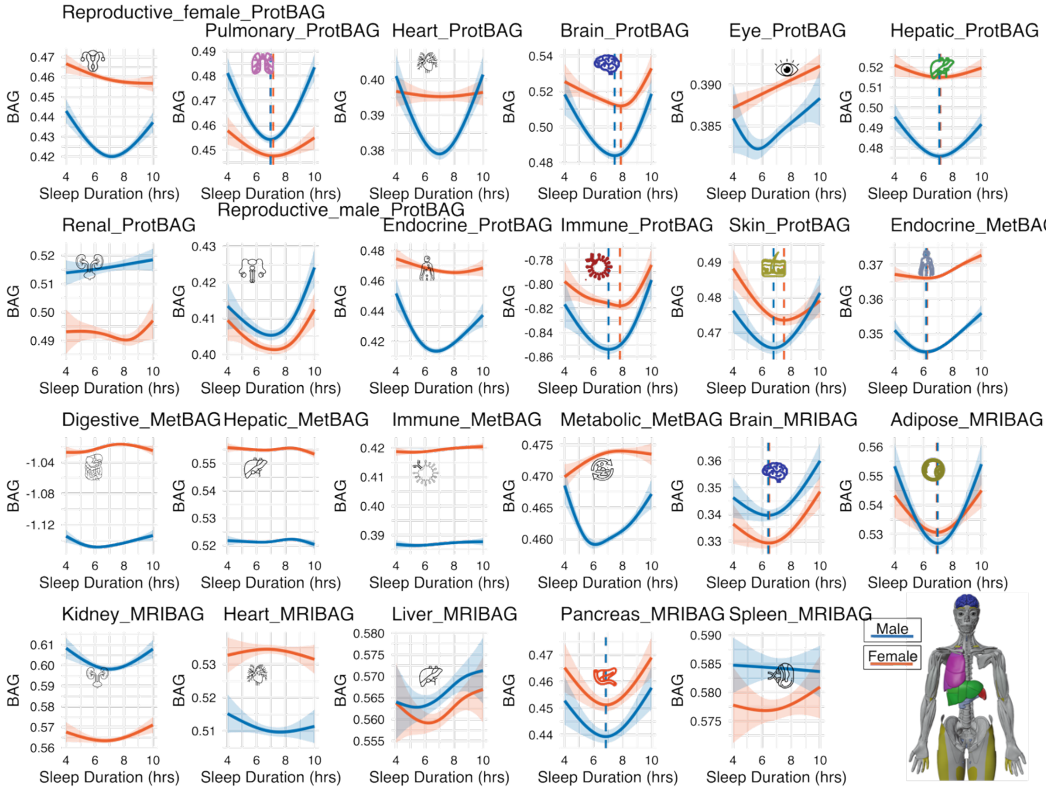


Generalized additive models were fit in the UK Biobank after adding the age-sex interaction term in the original GAM model. Dashed vertical lines indicate the sex-specific sleep duration corresponding to the minimum predicted BAG (“optimal” sleep). Colored icons correspond to the 9 significant BAGs identified in **Fig. 1**. The age–sex interaction term was significant for several BAGs, consistent with the age-stratified results shown in **Supplementary Fig. 4**, where the estimated “optimal” sleep duration differs between the two age groups. Nevertheless, the overall U-shaped association remains, with only minor shifts in the curves. Sex differences in the sleep–BAG associations may also be influenced by sex-specific reporting/measurement differences in self-reported sleep duration, as well as by other sleep exposures with marked sex disparities, such as higher prevalence of insomnia/restless legs syndrome in females and sleep apnea in males, which could confound or modify the observed relationships.

**Supplementary Figure 6: Sensitivity check analysis by excluding participants with 3 sleep-related diseases**

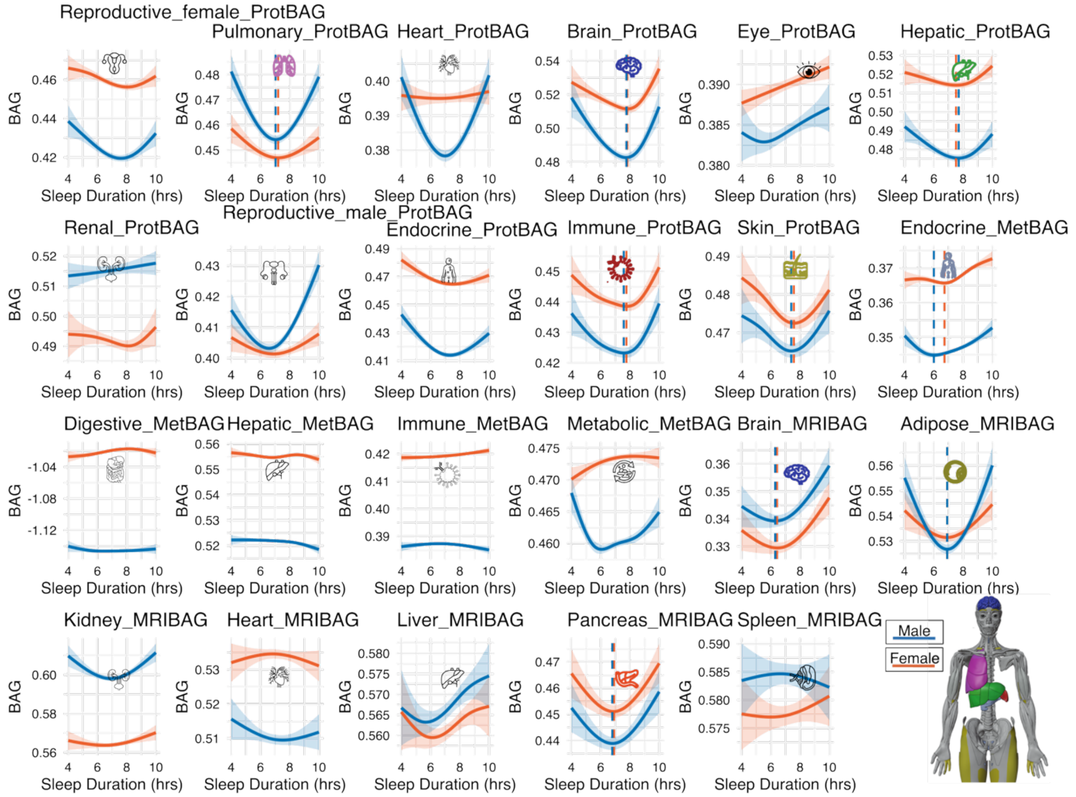


Generalized additive models were fit in the UK Biobank after removing individuals with any of the following sleep-related diagnoses (sleep apnea, insomnia, or restless legs syndrome). For each organ-specific BAG, we refit the same model as in **Fig. 1**. Dashed vertical lines indicate the sex-specific sleep duration corresponding to the minimum predicted BAG (“optimal” sleep). Colored icons correspond to the 9 significant BAGs identified in **Fig. 1**. After excluding participants with three types of sleep disorders, the U-shaped pattern persisted, although the estimated curve and the "optimal" sleep duration changed slightly.

**Supplementary Figure 7: Sensitivity check analysis by including smoking status as an additional covariate**

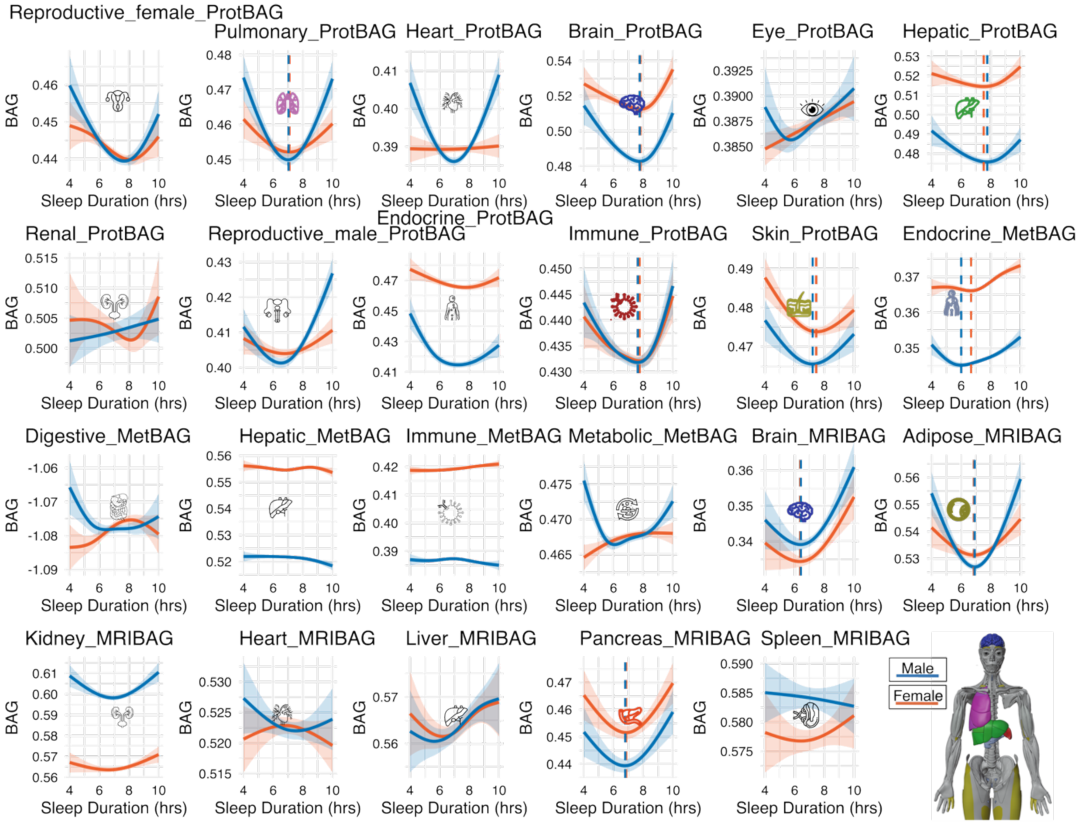


Generalized additive models were fit in the UK Biobank after adding smoking status (Field ID: 20116) as an additional covariate in the original GAM model. Dashed vertical lines indicate the sex-specific sleep duration corresponding to the minimum predicted BAG ("optimal" sleep). Colored icons correspond to the 9 significant BAGs identified in **Fig. 1**. After accounting for smoking status in the GAM, the U-shaped relationship pattern between sleep and BAG was robust, although the curve and the "optimal" sleep duration showed slight variations.

**Supplementary Figure 8: Sensitivity check analysis by including alcohol consumption status as an additional covariate**

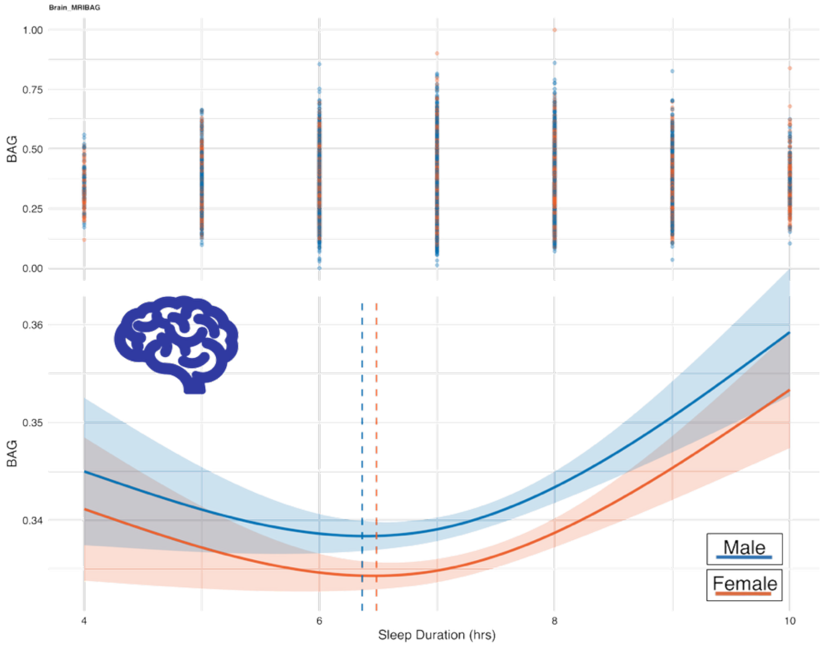


Generalized additive models were fit in the UK Biobank after adding alcohol consumption status (Field ID: 20117) as an additional covariate in the original GAM model for the brain MRIBAG (our data only covers the MRIBAG population for this variable). Dashed vertical lines indicate the sex-specific sleep duration corresponding to the minimum predicted BAG (“optimal” sleep).

**Supplementary Figure 9: Sensitivity check analysis by including Townsend deprivation index as an additional covariate**

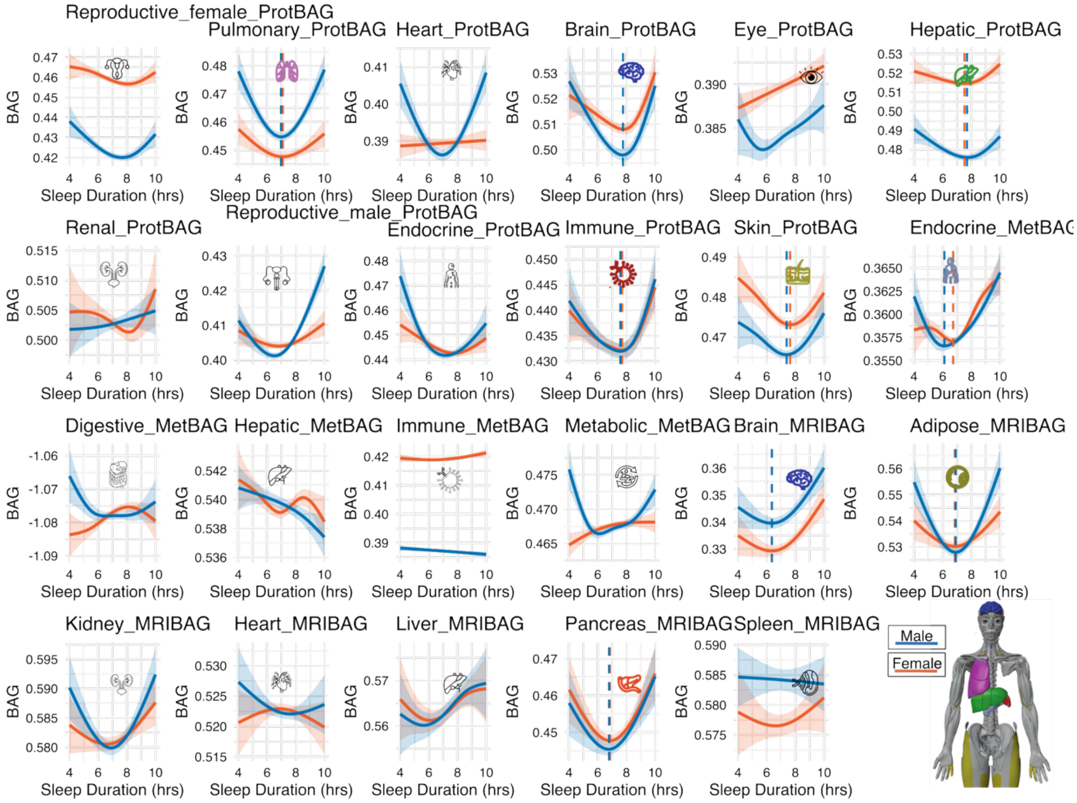


Generalized additive models were fit in the UK Biobank after adding the Townsend deprivation index (TDI; Field ID: 22189) as an additional covariate in the original GAM model. Dashed vertical lines indicate the sex-specific sleep duration corresponding to the minimum predicted BAG ("optimal" sleep). Colored icons correspond to the 9 significant BAGs identified in **Fig. 1**. After accounting for TDI in the GAM, the U-shaped relationship pattern between sleep and BAG was robust, although the curve and the "optimal" sleep duration showed slight variations (e.g., endocrine MetBAG).

**Supplementary Figure 10: Sensitivity check analysis by including computer use time as an additional covariate**

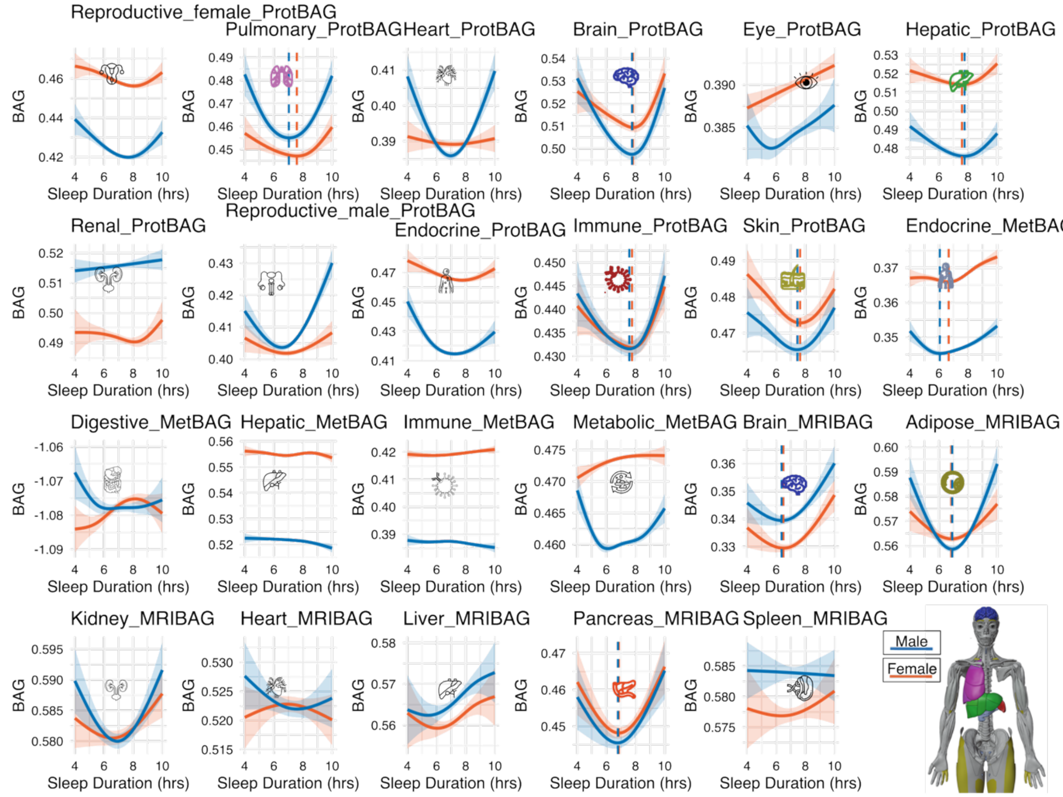


Generalized additive models were fit in the UK Biobank after adding the computer use time (Field ID: 1080) as an additional covariate in the original GAM model. Dashed vertical lines indicate the sex-specific sleep duration corresponding to the minimum predicted BAG (“optimal” sleep). Colored icons correspond to the 9 significant BAGs identified in **Fig. 1**. After accounting for computer use time in the GAM, the U-shaped relationship pattern between sleep and BAG was robust, although the curve and the "optimal" sleep duration showed slight variations.

**Supplementary Figure 11: Sensitivity check analysis by testing the interaction of sleep duration and testosterone**

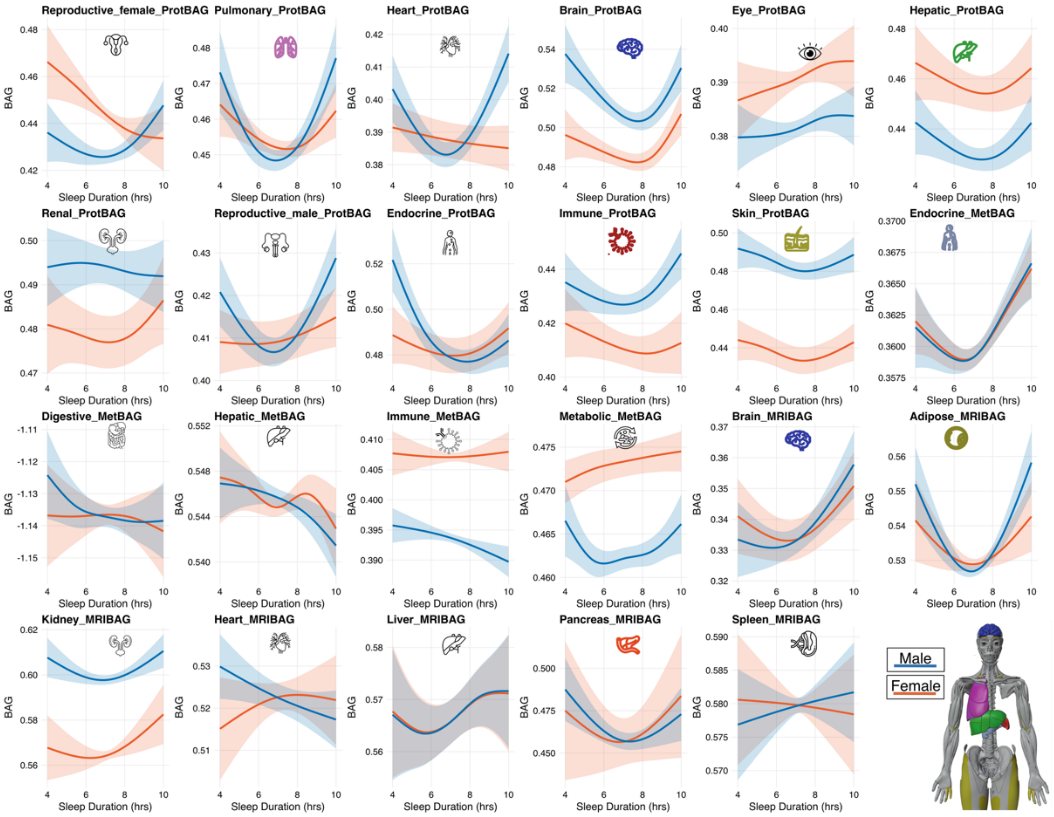


We refit the GAMs to include testosterone as a covariate and an interaction term with sleep duration (sleep duration × testosterone). None of the interaction effects reached significance (all P > 0.05), and while the 9 U-shaped associations remained, based on the results from **Fig. 1** in the main text, their curvature or inflection points shifted slightly in a few BAGs. Of note, the sample sizes reduced slightly compared to **Fig. 1**, because not all the full UKBB samples have data available for testosterone (i.e., UKBB only covers 426,570 participants out of the 500k population before merging with the BAG populations: https://biobank.ndph.ox.ac.uk/ukb/field.cgi?id=30850).

**Supplementary Figure 12: Sensitivity check analysis by testing the interaction of sleep duration and SHBG**

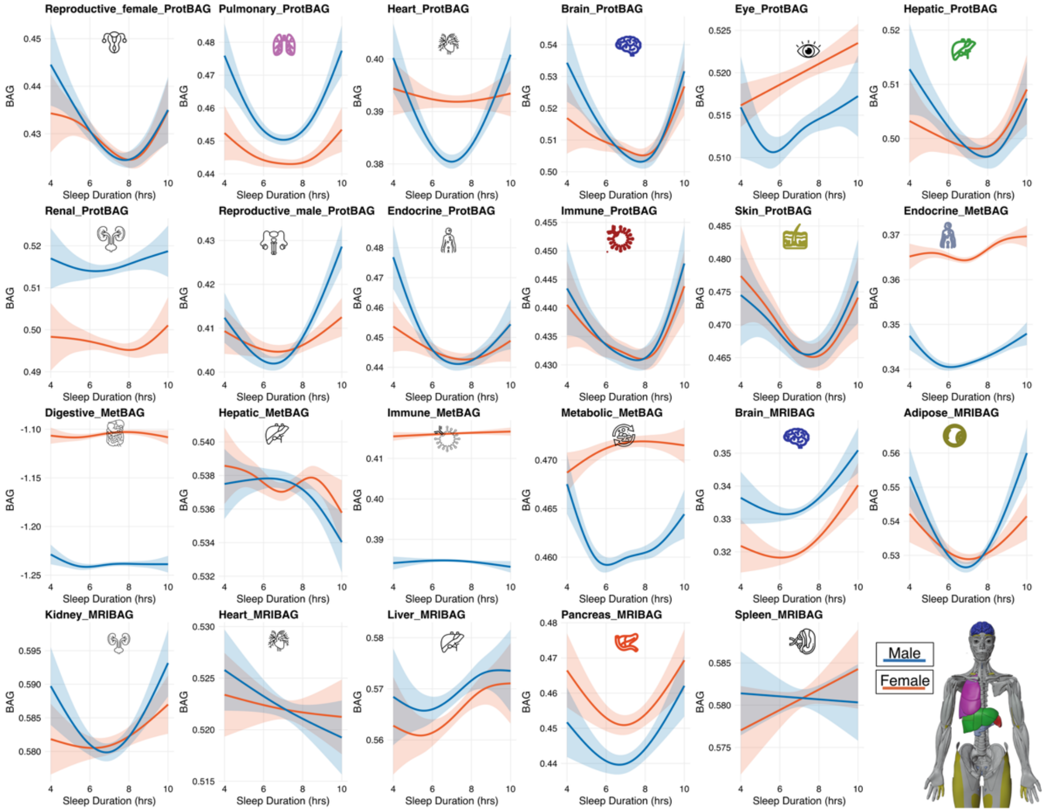


We refit the GAMs to include SHBG as a covariate and an interaction term with sleep duration (sleep duration × SHBG). None of the interaction effects reached significance (all $P > 0.05$), and while the 9 U-shaped associations remained, based on the results from **Fig. 1** in the main text, their curvature or inflection points shifted slightly in a few BAGs. Of note, the sample sizes reduced slightly compared to **Fig. 1**, because not all the full UKBB samples have data available for SHBG (i.e., UKBB only covers 427,697 participants out of the 500k population before merging with the BAG populations: https://biobank.ndph.ox.ac.uk/ukb/field.cgi?id=30830).

**Supplementary Figure 13: Sensitivity check analysis by testing the interaction of sleep duration and estradiol**

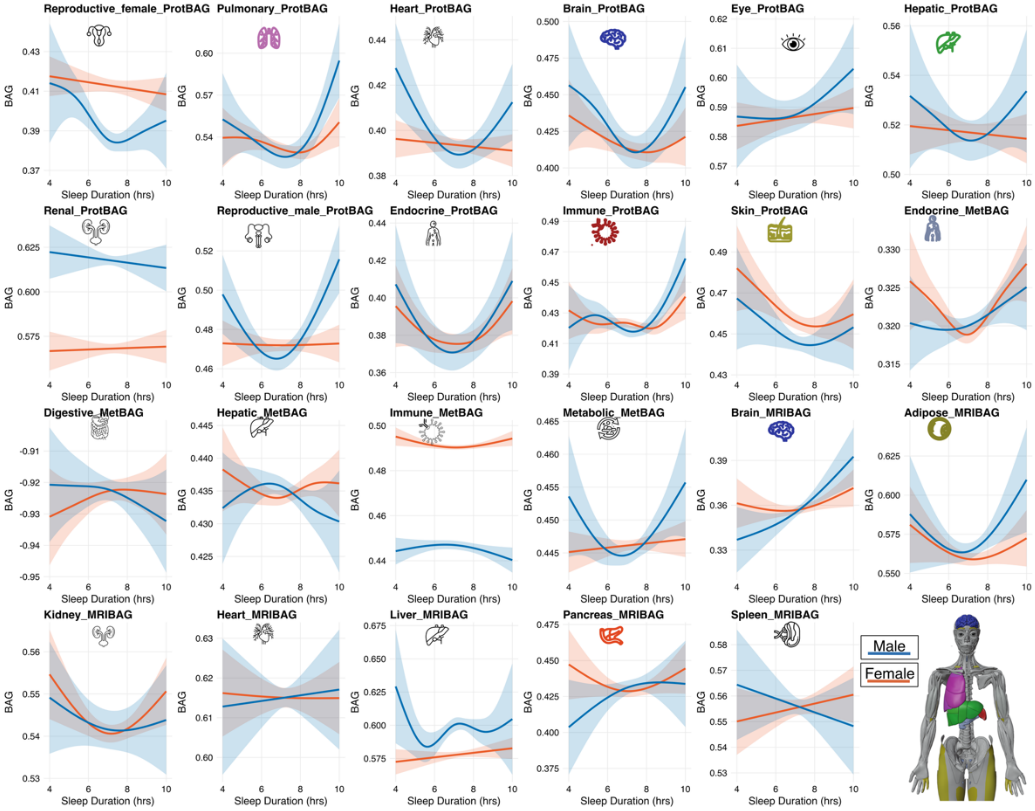


We refit the GAMs to include estradiol as a covariate and an interaction term with sleep duration (sleep duration × estradiol). Adding estradiol altered the fitted curves and, for the hepatic ProtBAG, the sleep × estradiol interaction was nominally significant (P=0.0196). That means the association between sleep duration and hepatic BAG appears to depend on estradiol level, e.g., the U-shape's curvature/inflection may differ at low vs. high estradiol. Of note, the sample sizes reduced dramatically compared to **Fig. 1**, because not all the full UKBB samples have data available for estradiol (i.e., UKBB only covers 77,594 participants out of the 500k population before merging with the BAG populations: https://biobank.ndph.ox.ac.uk/ukb/field.cgi?id=30800).

**Supplementary Figure 14: GAMLSS modeling of the 23 BAGs**

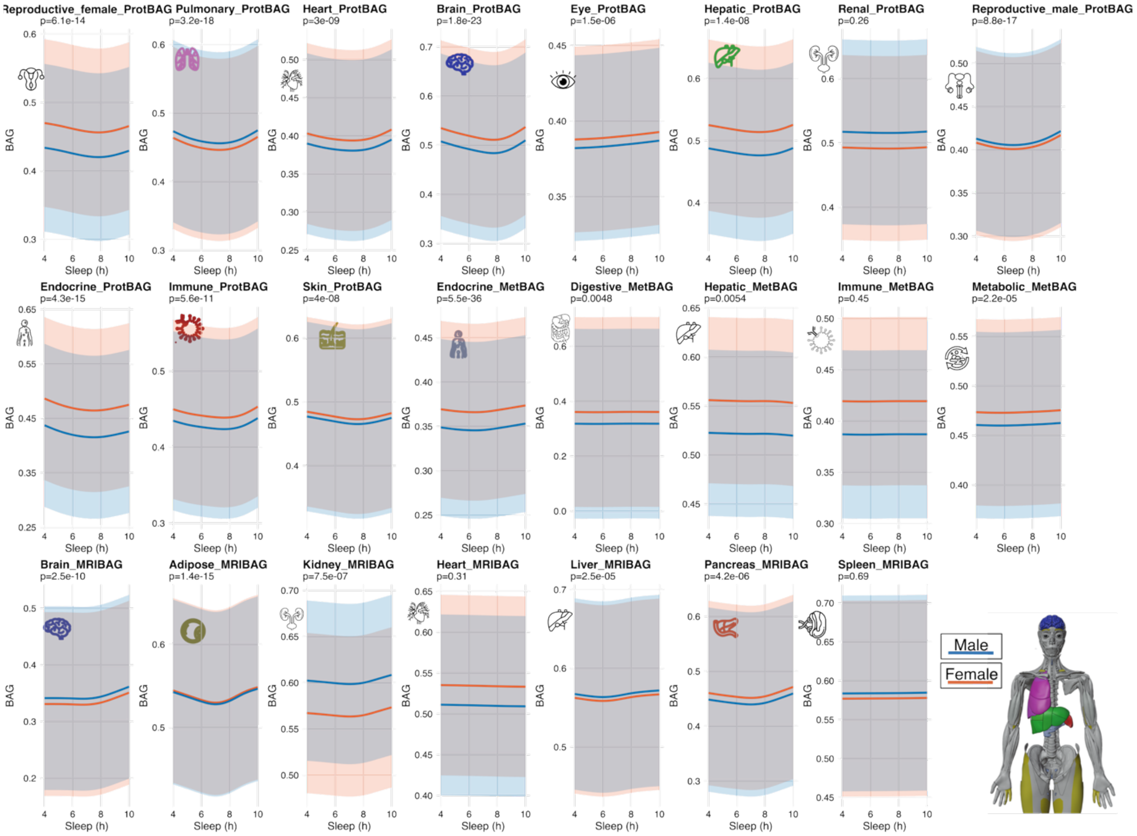


All 23 sleep–BAG associations were modelled using the *gamlss::gamlss()* model, with consistent U-shaped patterns observed across the 9 significant signals identified in Fig. 1, although minor differences exist due to different modelling assumptions. This likely reflects the more conservative nature of the gamlss framework, which not only models the mean but also incorporates variance and shape parameters, often smoothing over subtle group-specific interactions. In contrast, *mgcv::gam()* flexibly fits group-specific splines (e.g., *s(sleep_duration, by = sex)*), which can accentuate interaction effects. Despite harmonizing settings such as the smoothing method, number of knots, and family distribution, the underlying assumptions and modeling focus differ: mgcv emphasizes flexibility in mean structure. At the same time, gamlss prioritizes stable distributional fitting, potentially underestimating sex-sleep interactions in the presence of noisy predictors like self-reported sleep duration. These results highlight that the associations are robust across distinct statistical frameworks, despite differences in how they handle complexity and interaction.

**Supplementary Figure 15: External validation of the U-shaped patterns using MESA self-reported sleep duration**

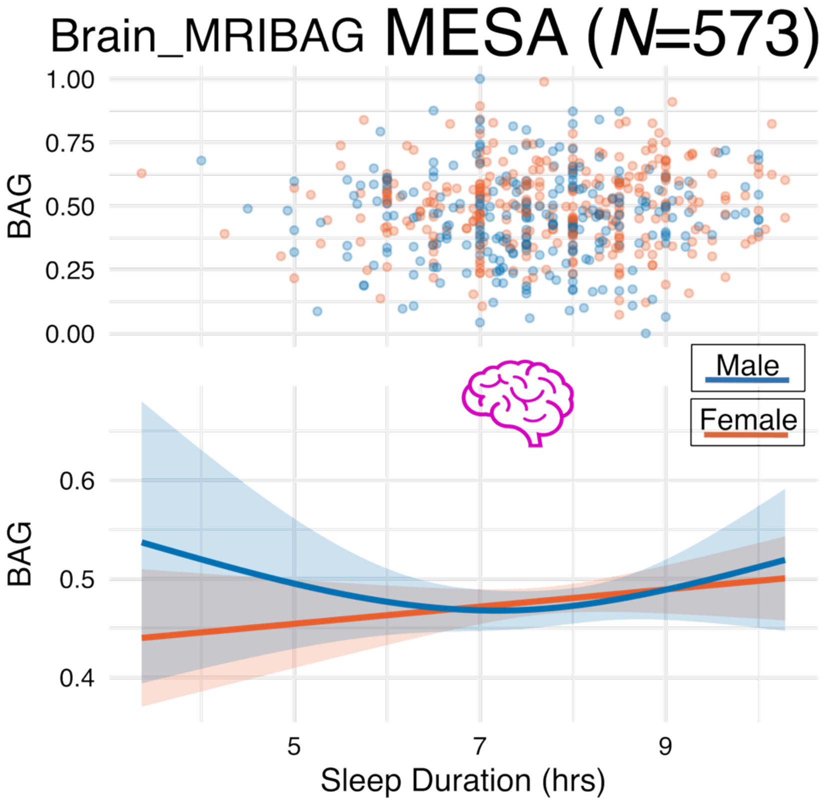


Using data from 573 MESA participants, we applied the same GAM model to examine the association between brain MRIBAG and self-reported sleep duration, calculated as the average of weekday and weekend sleep durations measured separately. In the MESA cohort, we observed a weak U-shaped relationship between sleep duration and brain MRIBAG, particularly among males, although this pattern did not reach statistical significance (P1=0.47). Several factors may contribute to the attenuated signal in MESA compared to the UK Biobank. First, the sample size in MESA was substantially smaller (*N*=573), limiting statistical power to detect subtle non-linear associations. Second, the MESA population is older on average (mean age of 72.06 years), which may reduce variability in sleep duration and increase the influence of age-related comorbidities and neurodegenerative changes, thereby masking the effect of sleep. Sex differences may also play a role, as the U-shaped pattern was visually evident only in males, suggesting potential biological or behavioral differences in sleep-brain aging dynamics.

**Supplementary Figure 16: External validation of the U-shaped patterns based on BLSA actigraphy-derived sleep duration**

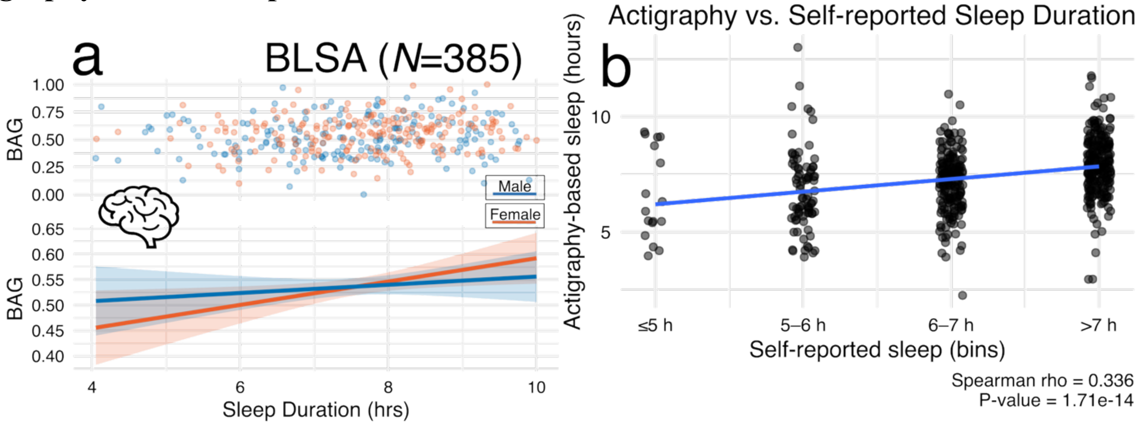


**a**) Actigraphy-derived sleep duration (hours) is not linked to the brain MRIBAG among BLSA participants (*N*=385; P=0.40). Lines represent sex-specific with shaded 95% confidence intervals. No significant associations were observed, likely due to the limited statistical power from the small sample size and the relatively healthy, older BLSA cohort (predominantly >70 years), which may exhibit a narrower range of both sleep and brain aging phenotypes. The absence of a U-shaped pattern often observed in younger or more heterogeneous populations could reflect physiological adaptation in late life, where sleep variability may exert weaker effects on brain aging due to homeostatic and circadian adjustments associated with aging. **b**) Correlation between actigraphy-based and self-reported sleep duration, indicating only a modest concordance between objective and subjective measures, echoing previous studies[9,10]. The U-shaped association between sleep duration and biological age gap observed with self-reported sleep likely reflects perceived sleep adequacy and long-term behavioral patterns, which integrate both physiological and psychosocial dimensions of sleep health. In contrast, actigraphy-based sleep provides a more objective, momentary measure of sleep–wake activity, capturing nightly variability but less influenced by subjective factors such as fatigue, sleep satisfaction, or chronic insomnia symptoms. Self-reports tend to overestimate sleep duration and incorporate cognitive and emotional perceptions of restfulness, which are closely tied to overall well-being and biological aging processes. Conversely, actigraphy may detect shorter but fragmented sleep periods that do not necessarily correspond to self-perceived sleep deficiency. As a result, self-reported sleep duration may better reflect long-term sleep behaviors and their cumulative physiological impact, whereas actigraphy captures short-term sleep patterns that might not yet manifest as measurable biological aging differences.

**Supplementary Figure 17: Sensitivity check of the ProWAS for protein FGFR2 of sleep duration mediated by the AST/ALT ratio**

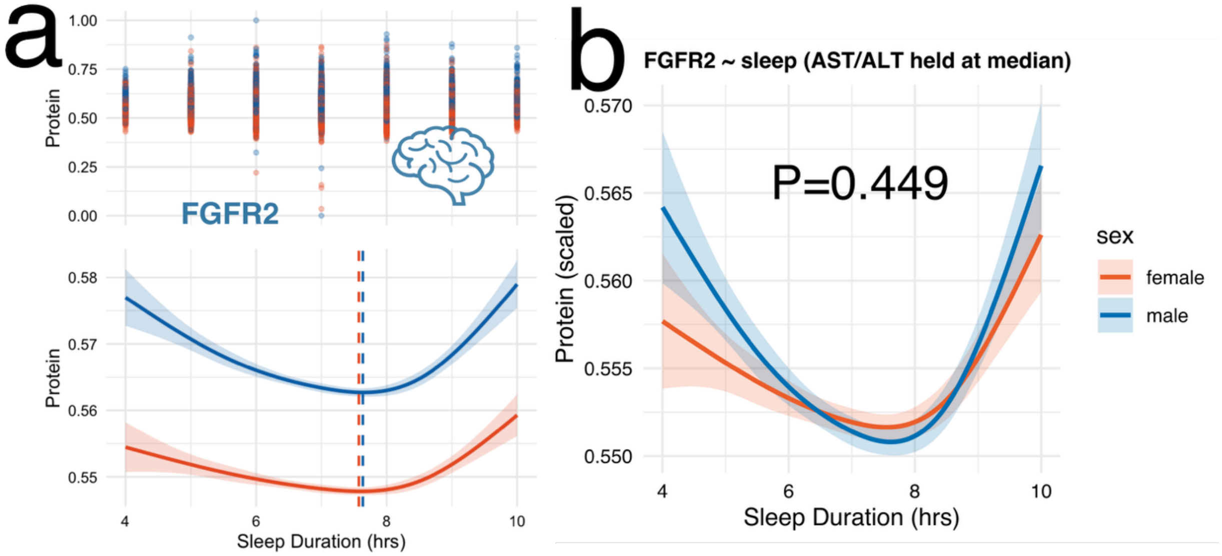


**a**) Original ProWAS results for FGFR2. **b**) Results after including AST/ALT as an additional covariate and its interaction with sleep duration. The interaction term was not statistically significant (P-value>0.05), although the U-shaped association pattern showed a slight change.

**Supplementary Figure 18: QQ and Manhattan plots of the GWAS results**

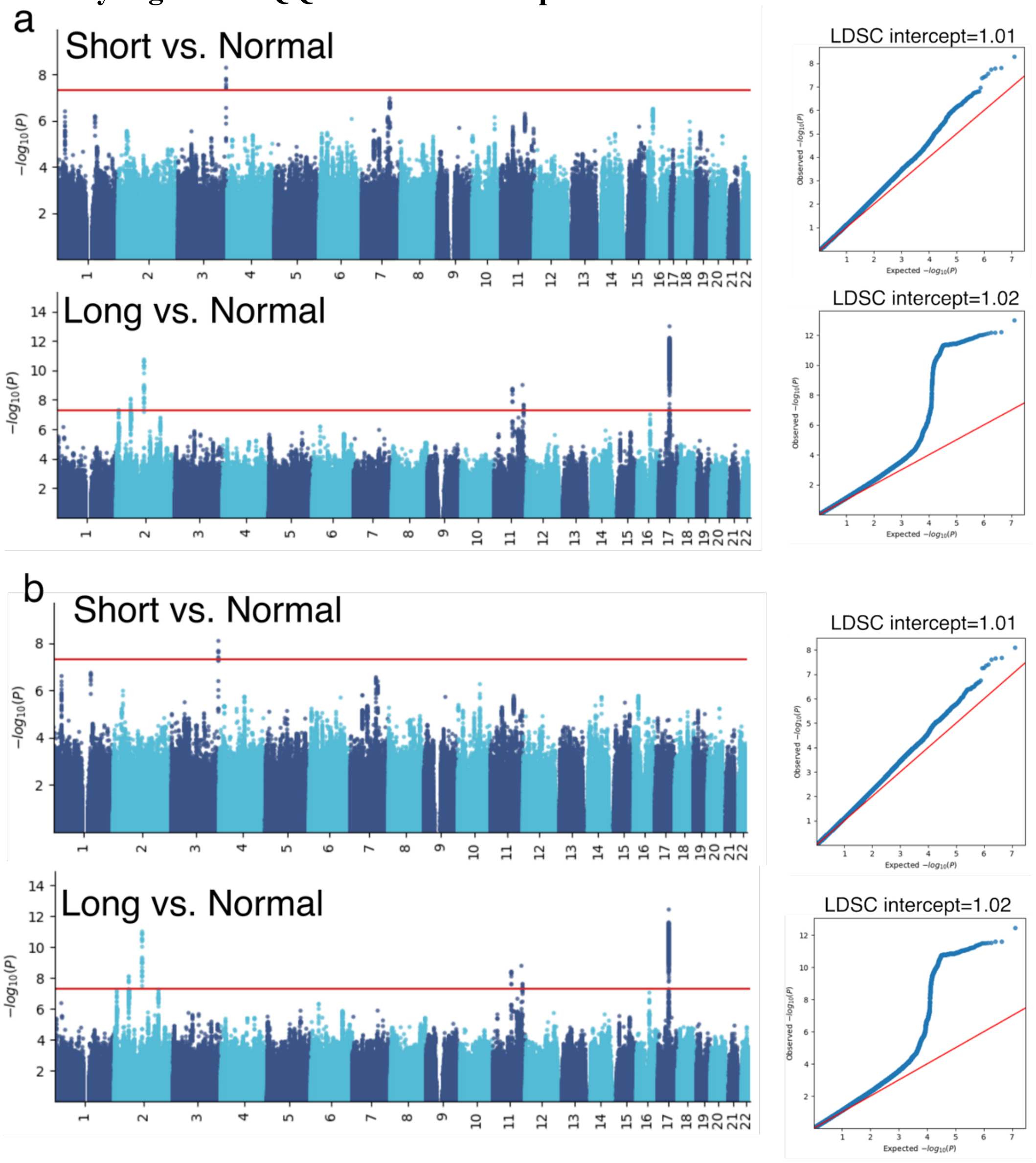


**a**) We present Manhattan and QQ plots for the two GWAS results: short vs. normal sleep duration and long versus normal sleep duration. The LDSC intercepts were close to 1, suggesting minimal bias from population structure. **b**) Manhattan and QQ plots for the two GWASs by adding additional covariates, including smoke status, Townsend deprivation index, and computer use time, in the original GWASs.

**Supplementary Figure 19: Genetic correlation between our binary sleep GWAS with two previous GWASs from Dashti et al and Austin-Zimmerman et al**

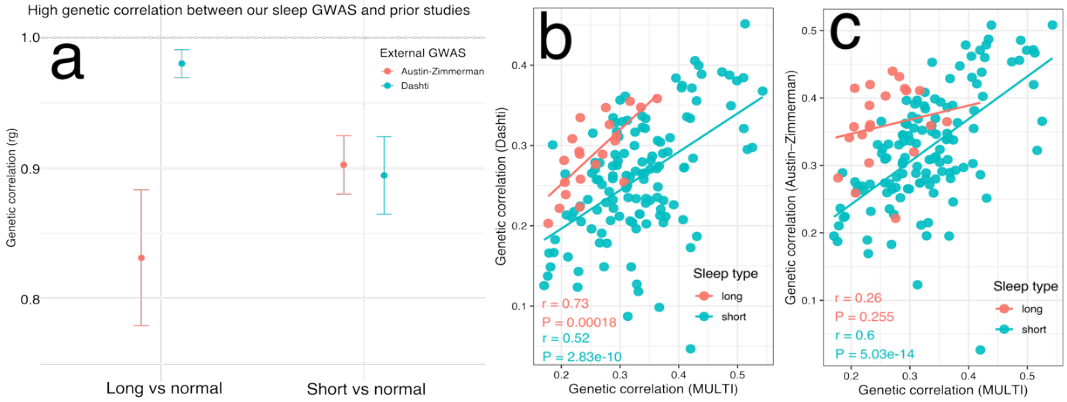


**a**) We downloaded GWAS summary statistics from Dashti et al. and Austin-Zimmerman et al. and used LDSC to estimate pairwise genetic correlations between our GWAS and each of these studies for short vs. normal and long vs. normal sleep. We observed consistently high genetic correlations, indicating that differences in the number of genomic loci across studies are largely driven by differences in sample size and in the operational definitions of short, normal, and long sleep duration. **b**) Using FinnGen, we demonstrate that the significant DE-sleep signals we found in our GWAS correlate with signals in the sleep GWAS summary statistics of Dashti et al. **c**) Using FinnGen, we demonstrate that the significant DE-sleep signals we found in our GWAS correlate with signals in the sleep GWAS summary statistics of Austin-Zimmerman et al.

**Supplementary Figure 20: Genetic correlation between FinnGen DEs and previous sleep GWASs from Dashti et al**

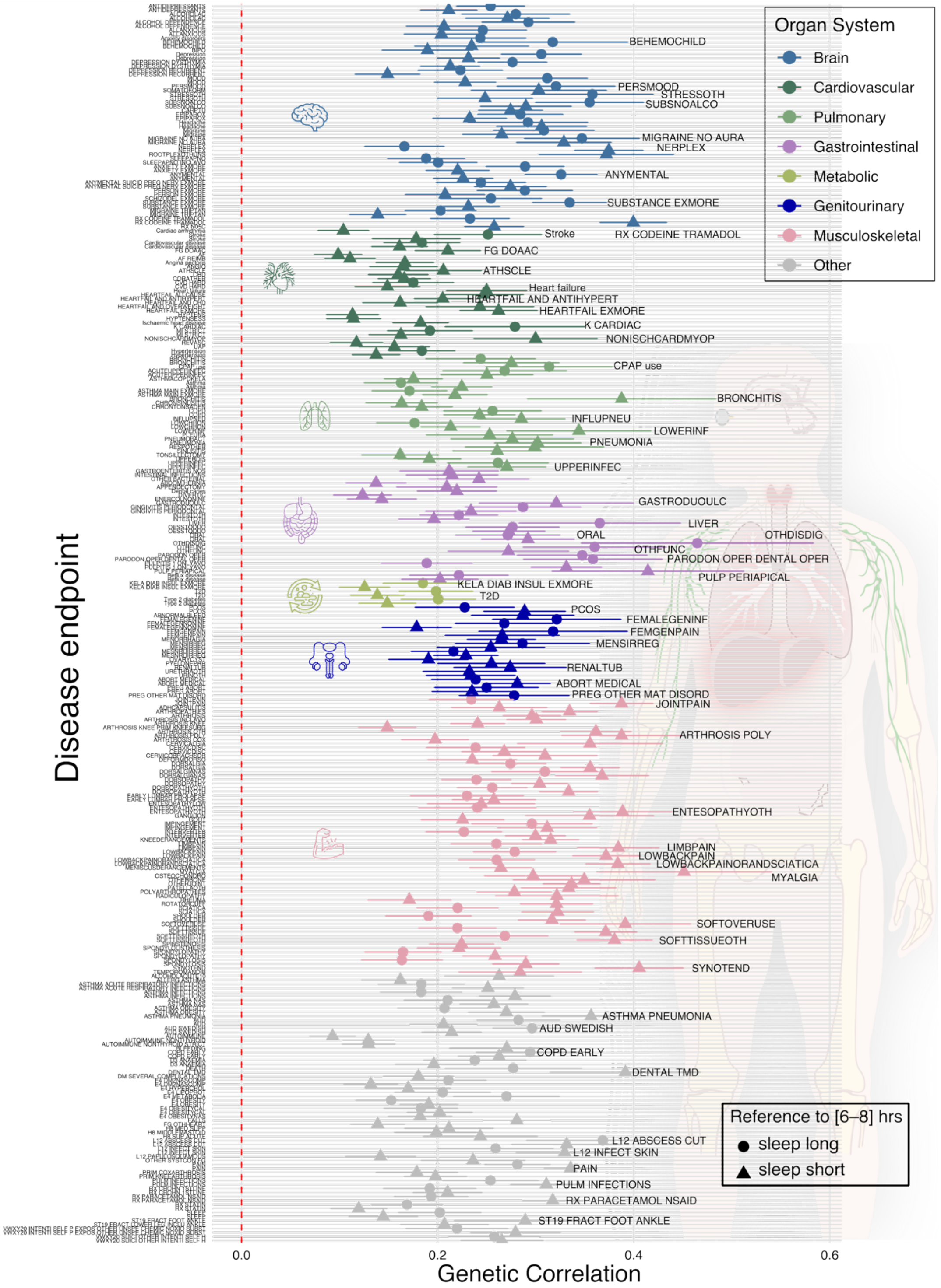


Genetic correlations (LDSC; mean ± s.e.) were shown between short sleep vs. normal sleep (triangles) and long sleep vs. normal sleep (circles) and 309 disease endpoints (DEs) from FinnGen (P-value<0.05/527). Points are colored by organ system and ordered on the y-axis from the brain (top) to other (bottom) to reflect approximate human anatomy. Horizontal error bars denote ±1 s.e., and the dashed red-vertical line indicates zero genetic correlation. For interpretability, the disease names shown as text annotations correspond to the top 10 representative DEs within each organ category (ranked by the smallest available p-value for the sleep–DE genetic correlation; otherwise by the largest absolute genetic correlation). **Supplementary File 5b** presents detailed statistics for our genetic correlation analysis.

**Supplementary Figure 21: Genetic correlation between FinnGen DEs and previous sleep GWASs from Austin-Zimmerman et al**

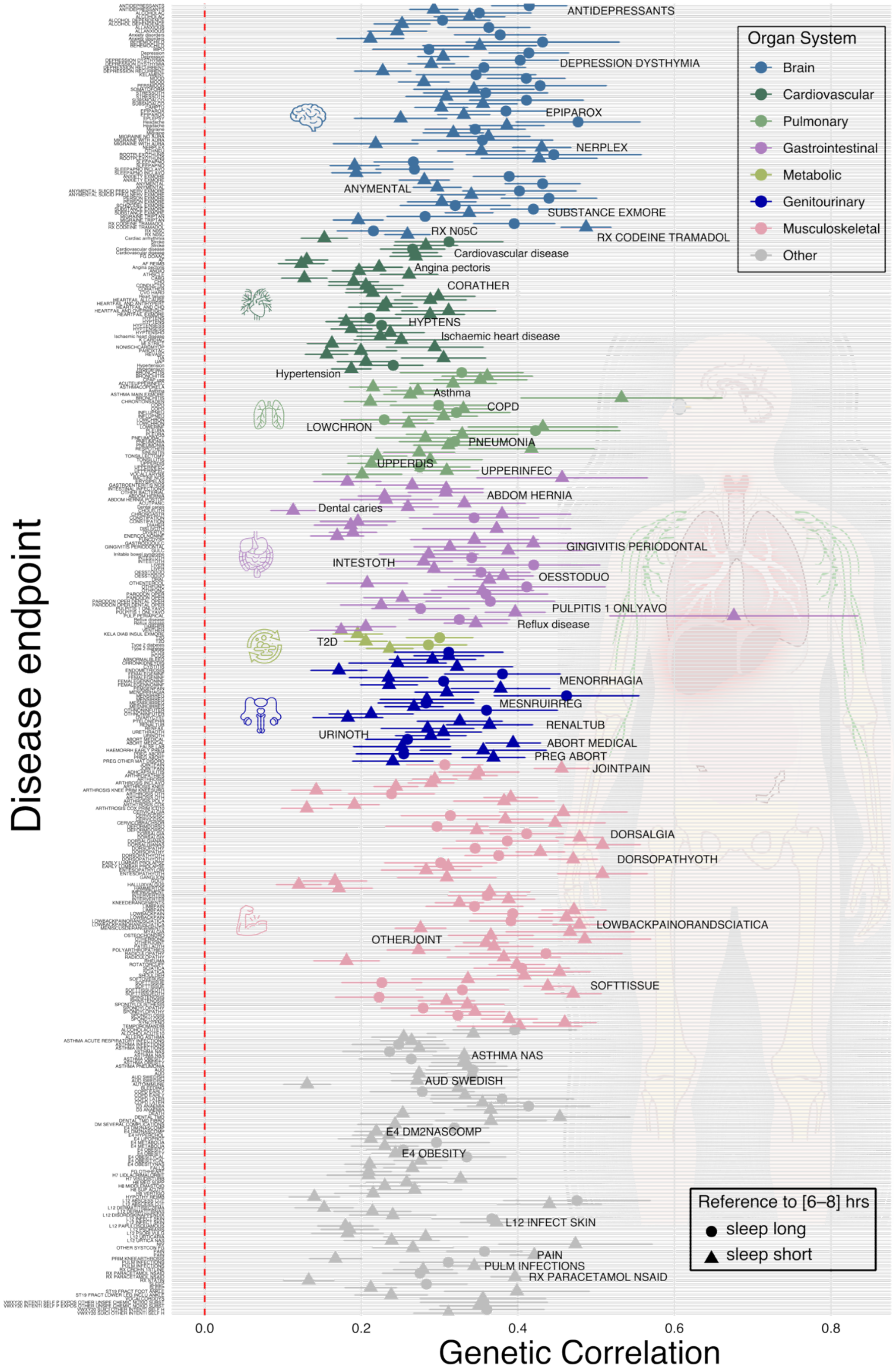


Genetic correlations (LDSC; mean ± s.e.) were shown between short sleep vs. normal sleep (triangles) and long sleep vs. normal sleep (circles) and 361 disease endpoints (DEs) from

FinnGen (P-value<0.05/527). Points are colored by organ system and ordered on the y-axis from the brain (top) to other (bottom) to reflect approximate human anatomy. Horizontal error bars denote ±1 s.e., and the dashed red-vertical line indicates zero genetic correlation. For interpretability, the disease names shown as text annotations correspond to the top 10 representative DEs within each organ category (ranked by the smallest available p-value for the sleep–DE genetic correlation; otherwise by the largest absolute genetic correlation). **Supplementary File 5c** presents detailed statistics for our genetic correlation analysis.

**Supplementary Figure 22: Sensitivity analysis using 7–9 hours as the reference category for normal sleep duration in disease incidence prediction**

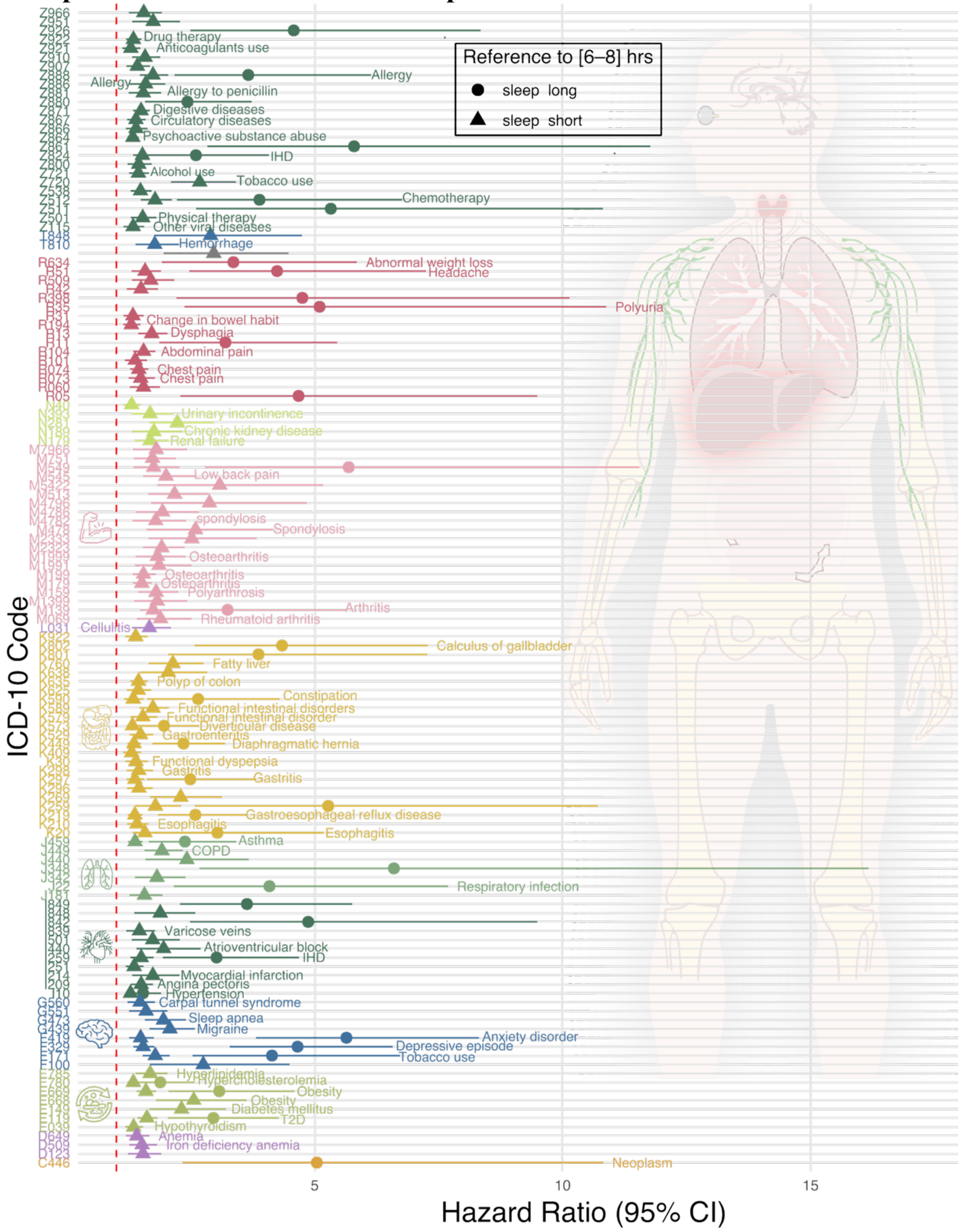


We re-ran the survival analysis using 7–9 hours as the reference for normal sleep duration and confirmed replication of the main sleep–DE associations observed in the primary analysis.

**Supplementary Table 1: The characteristics of participants consolidated via the MULTI study**

| Data type | BAG/Omics | Study | *N* | Age | Sex (female) |
|---|---|---|---|---|---|
| Individual | 7 MRIBAG | UKBB | 43,135 | 64.15±7.53 | 19,744/53% |
| | 11 ProtBAG | | 50,316 | 64.16±8.04 | 28,581/54% |
| | 5 MetBAG | | 274,247 | 56.56±8.08 | 147,994/54% |
| | Genetics | | 342,341 | 56.80±7.91 | 181,440/53% |
| | Sleep duration (self-report) | | 494,951 | 56.53±8.09 | 268,970/54% |
| | Disease endpoints | TriNetX[a] | 162,253,371 | 47.00±25.00 | 84,372,753/52% |
| | Sleep duration[c] (self-report & actigraphy) & Brain MRIBAG | BLSA | 385 | 70.07±13.45 | 206/53% |
| | Sleep duration (self-report) & Brain MRIBAG | MESA | 573 | 72.06±6.63 | 315/55% |
| Summary | Genetics | FinnGen | 521 | NA[b] | NA[b] |
| | | PGC | 6 | NA[b] | NA[b] |

[a]A detailed table for TriNex is presented in **Table 8**.
[b]FinnGen and PGC only provide summary-level GWAS statistics; individual-level age and sex information are unavailable.
[c]In BLSA, sleep duration was assessed through both actigraphy and self-reported questionnaires. For the questionnaire-based measure, participants were asked: ‘On average, in the past month, how many hours of sleep did you get each night?’ with the following response options: ‘More than 7 hours,’ ‘More than 6, up to 7 hours,’ ‘More than 5, up to 6 hours,’ ‘5 hours or fewer’, ‘Didn’t know’, and ‘Refuse to answer’. This categorical format differs from the -UK Biobank, which recorded sleep duration as rounded hourly values.

**Supplementary Table 2: Statistics of the GAM analysis between sleep duration and the 23 multi-organ, multi-omics biological aging clocks**

For each BAG, we fitted a generalized additive model (GAM) with cubic regression splines to assess the nonlinear association between sleep duration and BAG, stratified by sex and sex-sleep interaction term. Model selection was performed by evaluating combinations of smoothing dimensions (*k*=1, 3, 5, 10, 15, 20) and GAM families (i.e., Gaussian, t-distribution, and gamma distribution), with the optimal model chosen based on the lowest Akaike information criterion. We tested *i*) the main effect (sleep duration; $P_1$), *ii*) sex difference in population mean ($P_2$), and *iii*) sex-sleep interaction terms ($P_3$) on each BAG, including relevant covariates. Using bold text, we denote the significant signals (P-value<0.05/23). Of note, EDF reflects the complexity of the smoothing term, serving as an indicator of non-linearity in the modeled relationship rather than the model's overall effect size and fit to the outcome (BAG). The full stats for the covariates are included in **Supplementary File 1**. We used bold-text to highlight the 9 significant sleep-BAG signals identified in **Fig. 1**.

| BAG | Family | EDF | $P_1$ | $P_2$-coeff | $P_2$ | $P_3$ | Optimal (F) | Optimal (M) | N |
|---|---|---|---|---|---|---|---|---|---|
| Reproductive_female_ProtBAG | tdist | 3.12417885 | 0.00302409 | -0.036085 | 0 | 0.04204807 | 8.06060606 | 7.63636364 | 39450 |
| **Pulmonary_ProtBAG** | **tdist** | **3.20864257** | **0.00010919** | **0** | **NA** | **8.16E-05** | **7.63636364** | **7.03030303** | 39450 |
| Heart_ProtBAG | tdist | 1.36861696 | 0.78544472 | -0.0131752 | 2.58E-30 | 0 | 7.03030303 | 6.96969697 | 39450 |
| **Brain_ProtBAG** | **tdist** | **3.60700639** | **0** | **-0.0270772** | **0** | **0.06225086** | **7.81818182** | **7.6969697** | 39450 |
| Eye_ProtBAG | tdist | 1.01359891 | 0.22772588 | -0.0057015 | 1.11E-23 | 0.14519924 | 4 | 5.63636364 | 39450 |
| **Hepatic_ProtBAG** | **tdist** | **3.12884684** | **0.00136737** | **-0.0374589** | **1.5E-207** | **0.12538411** | **7.51515152** | **7.6969697** | 39450 |
| Renal_ProtBAG | tdist | 1.00023152 | 0.69806026 | 0 | NA | 0.70418125 | 8.12121212 | 4 | 39450 |
| Reproductive_male_ProtBAG | tdist | 1.89888028 | 0.01125616 | 0.00470686 | 1.26E-06 | 7.6986E-07 | 6.84848485 | 6.54545455 | 39450 |
| Endocrine_ProtBAG | tdist | 2.88686349 | 0.00236183 | -0.0490795 | 0 | 0.02173371 | 7.75757576 | 7.27272727 | 39450 |
| **Immune_ProtBAG** | **tdist** | **3.4725053** | **1.7753E-05** | **0** | **NA** | **0.34395093** | **7.75757576** | **7.57575758** | 39450 |
| **Skin_ProtBAG** | **tdist** | **3.21098138** | **3.1166E-06** | **-0.0075548** | **0** | **0.99394745** | **7.6969697** | **7.39393939** | 39450 |

| | | | | | | | | | |
|---|---|---|---|---|---|---|---|---|---|
| **Endocrine_MetBAG** | **tdist** | **1.0387028** | **3.968E-05** | **-0.0204961** | **0** | **5.71E-07** | **6.66666667** | **6.36867576** | 237781 |
| Digestive_MetBAG | gamma | 1.99159838 | 0.10923283 | 0 | NA | 0.06909867 | 4 | 6.3030303 | 244056 |
| Hepatic_MetBAG | tdist | 2.6392064 | 0.00518285 | 0 | 1 | 0.99924933 | 10 | 10 | 242727 |
| Immune_MetBAG | tdist | 1.01432365 | 0.00356703 | 0 | NA | 0.00632848 | 4.84848485 | 10 | 247310 |
| Metabolic_MetBAG | tdist | 1.03035596 | 0.13040643 | 0 | NA | 0 | 4 | 6.12121212 | 227395 |
| **Brain_MRIBAG** | **tdist** | **1.94299432** | **3.85E-07** | **0.01052919** | **0** | **0.84315248** | **6.48484848** | **6.42424242** | 33051 |
| **Adipose_MRIBAG** | **tdist** | **1.93399299** | **0.00028366** | **-0.0018702** | **0** | **0.00058391** | **6.90909091** | **6.90909091** | 22214 |
| Kidney_MRIBAG | tdist | 1.85929874 | 0.04736254 | 0 | NA | 0.03085264 | 6.60606061 | 6.90909091 | 31584 |
| Heart_MRIBAG | tdist | 1.00145304 | 0.76759541 | -0.0240222 | 0 | 0.23399526 | 10 | 7.57575758 | 32624 |
| Liver_MRIBAG | tdist | 2.56868492 | 0.05168732 | 0.00488097 | 0.00104338 | 0.48552144 | 6.12121212 | 5.57575758 | 22250 |
| **Pancreas_MRIBAG** | **tdist** | **1.95402342** | **9.9544E-06** | **-0.0118994** | **1.018E-09** | **0.71800183** | **6.84848485** | **6.78787879** | 25481 |
| Spleen_MRIBAG | tdist | 1.01340261 | 0.59509968 | 0.00674866 | 4.1389E-06 | 0.80134742 | 6.42424242 | 10 | 24411 |

**Supplementary Table 3: Sensitivity check analyses between sleep duration and the 23 BAGs**

We used bold-text to highlight the 9 significant sleep-BAG signals identified in **Fig. 1**. We present *i*) the main effect (sleep duration; $P_1$), *ii*) sex difference in population mean ($P_2$), and *iii*) sex-sleep interaction terms ($P_3$) on each BAG.

**a) Split-sample analysis**

| BAG | Family | Optimal k | EDF | $P_1$ | Split |
|---|---|---|---|---|---|
| Reproductive_female_ProtBAG | tdist | 5 | 3.40990517 | 4.32E-07 | Split1 |
| **Pulmonary_ProtBAG** | **tdist** | **5** | **3.60464839** | **0** | **Split1** |
| Heart_ProtBAG | tdist | 3 | 1.94577602 | 4.49E-05 | Split1 |
| **Brain_ProtBAG** | **tdist** | **5** | **3.54561137** | **0** | **Split1** |
| Eye_ProtBAG | tdist | 5 | 1.68030972 | 0.00130492 | Split1 |
| Hepatic_ProtBAG | tdist | 5 | 3.25321596 | 1.52E-06 | Split1 |
| Renal_ProtBAG | tdist | 5 | 2.25456061 | 0.46852205 | Split1 |
| Reproductive_male_ProtBAG | tdist | 3 | 1.97650719 | 0 | Split1 |
| Endocrine_ProtBAG | tdist | 5 | 3.10868703 | 0.00014551 | Split1 |
| **Immune_ProtBAG** | **tdist** | **5** | **3.32770689** | **0.00417126** | **Split1** |
| **Skin_ProtBAG** | **tdist** | **5** | **3.06500098** | **2.22E-05** | **Split1** |
| **Endocrine_MetBAG** | **tdist** | **5** | **3.31403875** | **0** | **Split1** |
| Digestive_MetBAG | gamma | 5 | 2.86811232 | 0.00749071 | Split1 |
| Hepatic_MetBAG | tdist | 5 | 3.42593569 | 0.00172921 | Split1 |
| Immune_MetBAG | tdist | 3 | 1.17866086 | 0.83085718 | Split1 |
| Metabolic_MetBAG | tdist | 5 | 2.50322362 | 0.00078332 | Split1 |
| **Brain_MRIBAG** | **tdist** | **3** | **1.89567361** | **0.00100219** | **Split1** |
| **Adipose_MRIBAG** | **tdist** | **3** | **1.96445018** | **0** | **Split1** |
| Kidney_MRIBAG | tdist | 3 | 1.92878969 | 0.0003637 | Split1 |
| Heart_MRIBAG | tdist | 3 | 1.00697631 | 0.18228465 | Split1 |
| Liver_MRIBAG | tdist | 5 | 2.6642057 | 0.00176361 | Split1 |
| **Pancreas_MRIBAG** | **tdist** | **3** | **1.9331389** | **9.03E-05** | **Split1** |
| Spleen_MRIBAG | tdist | 3 | 1.84051695 | 0.06429083 | Split1 |
| Reproductive_female_ProtBAG | tdist | 5 | 3.07378452 | 2.40E-06 | Split2 |
| **Pulmonary_ProtBAG** | **tdist** | **5** | **3.21715252** | **3.41E-07** | **Split2** |
| Heart_ProtBAG | tdist | 3 | 1.9537369 | 3.49E-05 | Split2 |
| **Brain_ProtBAG** | **tdist** | **5** | **3.46777297** | **0** | **Split2** |
| Eye_ProtBAG | tdist | 5 | 1.04558127 | 0.00034558 | Split2 |
| **Hepatic_ProtBAG** | **tdist** | **5** | **2.46189641** | **0.01697513** | **Split2** |
| Renal_ProtBAG | tdist | 5 | 1.65843831 | 0.49527592 | Split2 |
| Reproductive_male_ProtBAG | tdist | 3 | 1.93289922 | 9.83E-06 | Split2 |
| Endocrine_ProtBAG | tdist | 5 | 3.2131639 | 0 | Split2 |
| **Immune_ProtBAG** | **tdist** | **5** | **3.29984535** | **0** | **Split2** |
| **Skin_ProtBAG** | **tdist** | **5** | **2.99831599** | **0.00950557** | **Split2** |
| **Endocrine_MetBAG** | **tdist** | **5** | **3.3599701** | **0** | **Split2** |
| Digestive_MetBAG | gamma | 5 | 2.49705697 | 0.11742895 | Split2 |
| Hepatic_MetBAG | tdist | 5 | 1.55641378 | 0.25311001 | Split2 |
| Immune_MetBAG | tdist | 3 | 1.1798613 | 0.28855037 | Split2 |
| Metabolic_MetBAG | tdist | 5 | 2.12113067 | 0.04992084 | Split2 |
| **Brain_MRIBAG** | **tdist** | **3** | **1.91007336** | **1.05E-08** | **Split2** |
| **Adipose_MRIBAG** | **tdist** | **3** | **1.97245593** | **0** | **Split2** |
| Kidney_MRIBAG | tdist | 3 | 1.89660316 | 0.00242667 | Split2 |
| Heart_MRIBAG | tdist | 3 | 1.00691438 | 0.83872591 | Split2 |
| Liver_MRIBAG | tdist | 5 | 1.01535176 | 0.00531592 | Split2 |
| **Pancreas_MRIBAG** | **tdist** | **3** | **1.86852578** | **0.02436188** | **Split2** |
| Spleen_MRIBAG | tdist | 3 | 1.70219838 | 0.21934225 | Split2 |

**b) Age-stratified analysis**

| BAG | Age Group (yrs) | Family | Optimal k | EDF | $P_1$ |
|---|---|---|---|---|---|
| Reproductive_female_ProtBAG | [40-60] | tdist | 5 | 2.47929624 | 0.00010859 |
| Reproductive_female_ProtBAG | [60-80] | tdist | 5 | 3.28924924 | 9.98E-08 |
| Pulmonary_ProtBAG | [40-60] | tdist | 5 | 3.68528492 | 0 |
| Pulmonary_ProtBAG | [60-80] | tdist | 5 | 3.07717932 | 4.35E-07 |
| Heart_ProtBAG | [40-60] | tdist | 3 | 1.94462295 | 5.79E-05 |
| Heart_ProtBAG | [60-80] | tdist | 3 | 1.9508635 | 2.83E-06 |
| Brain_ProtBAG | [40-60] | tdist | 5 | 3.62099293 | 0 |
| Brain_ProtBAG | [60-80] | tdist | 5 | 3.37986947 | 0 |
| Eye_ProtBAG | [40-60] | tdist | 5 | 1.00940526 | 0.00623431 |
| Eye_ProtBAG | [60-80] | tdist | 5 | 1.59300727 | 5.66E-06 |
| Hepatic_ProtBAG | [40-60] | tdist | 5 | 2.70418844 | 0.00092072 |
| Hepatic_ProtBAG | [60-80] | tdist | 5 | 3.06908616 | 0.00010663 |
| Renal_ProtBAG | [40-60] | tdist | 5 | 1.01691156 | 0.26518242 |
| Renal_ProtBAG | [60-80] | tdist | 5 | 2.50885389 | 0.01038931 |
| Reproductive_male_ProtBAG | [40-60] | tdist | 3 | 1.96209676 | 2.29E-06 |
| Reproductive_male_ProtBAG | [60-80] | tdist | 3 | 1.95599299 | 0 |
| Endocrine_ProtBAG | [40-60] | tdist | 5 | 3.34820355 | 0 |
| Endocrine_ProtBAG | [60-80] | tdist | 5 | 3.02443553 | 3.63E-05 |
| Immune_ProtBAG | [40-60] | tdist | 5 | 3.43257411 | 4.42E-06 |
| Immune_ProtBAG | [60-80] | tdist | 5 | 2.99060222 | 5.42E-05 |
| Skin_ProtBAG | [40-60] | tdist | 5 | 2.98678363 | 0.00014909 |
| Skin_ProtBAG | [60-80] | tdist | 5 | 2.74687839 | 0.01336974 |
| Endocrine_MetBAG | [40-60] | tdist | 5 | 3.57871596 | 0 |
| Endocrine_MetBAG | [60-80] | tdist | 5 | 3.06890784 | 0 |
| Digestive_MetBAG | [40-60] | gamma | 5 | 1.15932782 | 0.00014016 |
| Digestive_MetBAG | [60-80] | gamma | 5 | 3.44468294 | 2.34E-05 |
| Hepatic_MetBAG | [40-60] | tdist | 5 | 3.24040023 | 0.04218487 |
| Hepatic_MetBAG | [60-80] | tdist | 5 | 3.03710161 | 0.09495551 |
| Immune_MetBAG | [40-60] | tdist | 3 | 1.83095236 | 0.00065235 |
| Immune_MetBAG | [60-80] | tdist | 3 | 1.54177702 | 0.00050281 |
| Metabolic_MetBAG | [40-60] | tdist | 5 | 2.6207173 | 4.41E-05 |
| Metabolic_MetBAG | [60-80] | tdist | 5 | 1.00667415 | 0.0008325 |
| Brain_MRIBAG | [40-60] | tdist | 3 | 1.93961673 | 4.11E-06 |
| Brain_MRIBAG | [60-80] | tdist | 3 | 1.66287313 | 0.05840601 |
| Adipose_MRIBAG | [40-60] | tdist | 3 | 1.97104816 | 0 |
| Adipose_MRIBAG | [60-80] | tdist | 3 | 1.96251605 | 3.39E-07 |
| Kidney_MRIBAG | [40-60] | tdist | 3 | 1.93981775 | 0.00015496 |
| Kidney_MRIBAG | [60-80] | tdist | 3 | 1.85182348 | 0.02694113 |
| Heart_MRIBAG | [40-60] | tdist | 3 | 1.00713063 | 0.15132763 |
| Heart_MRIBAG | [60-80] | tdist | 3 | 1.00868926 | 0.42449593 |
| Liver_MRIBAG | [40-60] | tdist | 5 | 1.30033668 | 0.01175562 |
| Liver_MRIBAG | [60-80] | tdist | 5 | 2.1660193 | 0.02186497 |
| Pancreas_MRIBAG | [40-60] | tdist | 3 | 1.80016153 | 0.13829659 |
| Pancreas_MRIBAG | [60-80] | tdist | 3 | 1.9447499 | 1.45E-05 |
| Spleen_MRIBAG | [40-60] | tdist | 3 | 1.0200997 | 0.72290745 |
| Spleen_MRIBAG | [60-80] | tdist | 3 | 1.00722982 | 0.68151139 |

**c) Sex-stratified analysis**

| BAG | Sex | Family | Optimal k | EDF | $P_1$ |
|---|---|---|---|---|---|
| Reproductive_female_ProtBAG | Female | tdist | 5 | 2.88048221 | 0.00056244 |
| Reproductive_female_ProtBAG | Male | tdist | 5 | 3.36851646 | 0 |
| **Pulmonary_ProtBAG** | **Female** | **tdist** | **5** | **3.01162763** | **0.00351465** |

| | | | | | |
|---|---|---|---|---|---|
| **Pulmonary_ProtBAG** | **Male** | **tdist** | **5** | **3.62453121** | **0** |
| Heart_ProtBAG | Female | tdist | 3 | 1.69323001 | 0.33620365 |
| Heart_ProtBAG | Male | tdist | 3 | 1.98176342 | 0 |
| **Brain_ProtBAG** | **Female** | **tdist** | **5** | **3.50572566** | **6.65E-07** |
| **Brain_ProtBAG** | **Male** | **tdist** | **5** | **3.5298186** | **0** |
| Eye_ProtBAG | Female | tdist | 5 | 1.0336401 | 0.00040973 |
| Eye_ProtBAG | Male | tdist | 5 | 3.06303629 | 2.53E-05 |
| **Hepatic_ProtBAG** | **Female** | **tdist** | **5** | **2.55688216** | **0.09725832** |
| **Hepatic_ProtBAG** | **Male** | **tdist** | **5** | **3.24561795** | **7.53E-07** |
| Renal_ProtBAG | Female | tdist | 5 | 2.9050947 | 0.10148904 |
| Renal_ProtBAG | Male | tdist | 5 | 1.20101925 | 0.21590821 |
| Reproductive_male_ProtBAG | Female | tdist | 3 | 1.96230451 | 2.19E-06 |
| Reproductive_male_ProtBAG | Male | tdist | 3 | 1.96535989 | 0 |
| Endocrine_ProtBAG | Female | tdist | 5 | 2.55659993 | 0.00508666 |
| Endocrine_ProtBAG | Male | tdist | 5 | 3.63940804 | 0 |
| **Immune_ProtBAG** | **Female** | **tdist** | **5** | **3.1698081** | **0.0091142** |
| **Immune_ProtBAG** | **Male** | **tdist** | **5** | **3.34800558** | **0** |
| **Skin_ProtBAG** | **Female** | **tdist** | **5** | **2.82703424** | **0.00103676** |
| **Skin_ProtBAG** | **Male** | **tdist** | **5** | **2.97219816** | **2.94E-05** |
| **Endocrine_MetBAG** | **Female** | **tdist** | **5** | **3.20835097** | **0** |
| **Endocrine_MetBAG** | **Male** | **tdist** | **5** | **3.66051872** | **0** |
| Digestive_MetBAG | Female | gamma | 5 | 3.14616228 | 1.60E-05 |
| Digestive_MetBAG | Male | gamma | 5 | 3.19365559 | 7.61E-06 |
| Hepatic_MetBAG | Female | tdist | 5 | 3.12204622 | 0.17469654 |
| Hepatic_MetBAG | Male | tdist | 5 | 2.85656673 | 0.1138149 |
| Immune_MetBAG | Female | tdist | 3 | 1.02242724 | 0.0005673 |
| Immune_MetBAG | Male | tdist | 3 | 1.02028543 | 0.00754742 |
| Metabolic_MetBAG | Female | tdist | 5 | 2.2384226 | 0.00185076 |
| Metabolic_MetBAG | Male | tdist | 5 | 3.77371641 | 0 |
| **Brain_MRIBAG** | **Female** | **tdist** | **3** | **1.90713689** | **2.06E-05** |
| **Brain_MRIBAG** | **Male** | **tdist** | **3** | **1.90690714** | **3.10E-05** |
| **Adipose_MRIBAG** | **Female** | **tdist** | **3** | **1.96388916** | **9.31E-07** |
| **Adipose_MRIBAG** | **Male** | **tdist** | **3** | **1.97042448** | **0** |
| Kidney_MRIBAG | Female | tdist | 3 | 1.92169393 | 0.00039019 |
| Kidney_MRIBAG | Male | tdist | 3 | 1.89530592 | 0.01012402 |
| Heart_MRIBAG | Female | tdist | 3 | 1.36932306 | 0.51730877 |
| Heart_MRIBAG | Male | tdist | 3 | 1.41461327 | 0.60453837 |
| Liver_MRIBAG | Female | tdist | 5 | 2.41880644 | 0.00600284 |
| Liver_MRIBAG | Male | tdist | 5 | 1.01205988 | 0.00568765 |
| **Pancreas_MRIBAG** | **Female** | **tdist** | **3** | **1.92706747** | **0.000531** |
| **Pancreas_MRIBAG** | **Male** | **tdist** | **3** | **1.87493757** | **0.01912217** |
| Spleen_MRIBAG | Female | tdist | 3 | 1.50248213 | 0.3868693 |
| Spleen_MRIBAG | Male | tdist | 3 | 1.00816804 | 0.75391225 |

**d) Age-sex interaction as an additional term**

| BAG | Family | Optimal k | EDF | $P_1$ | Age-sex coef | Age-sex P-value |
|---|---|---|---|---|---|---|
| Reproductive_female_ProtBAG | tdist | 3 | 1.68718855 | 0.00224312 | -0.0019905 | 2.72E-112 |
| **Pulmonary_ProtBAG** | **tdist** | **3** | **1.92699698** | **0.00104244** | **-0.0010592** | **1.20E-29** |
| Heart_ProtBAG | tdist | 3 | 1.00408756 | 0.63188287 | 0.00035741 | 9.05E-05 |
| **Brain_ProtBAG** | **tdist** | **5** | **3.51963313** | **1.15E-05** | **-0.0021568** | **9.55E-70** |

| | | | | | | |
|---|---|---|---|---|---|---|
| Eye_ProtBAG | tdist | 5 | 1.01885855 | 0.00031284 | -0.000359 | 1.55E-15 |
| **Hepatic_ProtBAG** | **tdist** | **3** | **1.83386282** | **0.00189172** | **-0.0019799** | **2.22E-94** |
| Renal_ProtBAG | tdist | 5 | 1.00067997 | 0.6212926 | -0.0002332 | 0.02001364 |
| Reproductive_male_ProtBAG | tdist | 5 | 3.28783379 | 2.58E-05 | 0.00261804 | 1.88E-261 |
| Endocrine_ProtBAG | tdist | 5 | 2.35731362 | 0.01982627 | -0.0035014 | 1.40E-265 |
| **Immune_ProtBAG** | **gamma** | **5** | **3.35238495** | **0.00724521** | **-0.0040787** | **3.53E-87** |
| **Skin_ProtBAG** | **tdist** | **3** | **1.94319598** | **0.00013296** | **-0.0024061** | **5.52E-126** |
| **Endocrine_MetBAG** | **tdist** | **5** | **3.14329301** | **0** | **-0.001135** | **0** |
| Digestive_MetBAG | gamma | 5 | 1.99008831 | 0.5609586 | -0.0038222 | 0 |
| Hepatic_MetBAG | tdist | 5 | 3.5085749 | 0.00590508 | -0.0011115 | 0 |
| Immune_MetBAG | tdist | 5 | 2.60771981 | 0.00492697 | -0.0010527 | 0 |
| Metabolic_MetBAG | tdist | 5 | 1.15110441 | 0.02825922 | -0.0006325 | 4.30E-116 |
| **Brain_MRIBAG** | **tdist** | **3** | **1.95036718** | **6.63E-07** | **0.00016203** | **0.22194206** |
| **Adipose_MRIBAG** | **tdist** | **3** | **1.94656684** | **0.00014905** | **0.00157058** | **3.09E-43** |
| Kidney_MRIBAG | tdist | 3 | 1.89000532 | 0.01594308 | 0.00076695 | 3.40E-23 |
| Heart_MRIBAG | tdist | 3 | 1.00838371 | 0.18279153 | 5.95E-05 | 0.51846454 |
| Liver_MRIBAG | tdist | 5 | 2.34729655 | 0.03189794 | 0.00043212 | 0.00061975 |
| **Pancreas_MRIBAG** | **tdist** | **3** | **1.95271412** | **1.99E-05** | **0.00086172** | **1.72E-07** |
| Spleen_MRIBAG | tdist | 3 | 1.00439983 | 0.80203926 | -0.0004056 | 0.00105952 |

##### e) Exclude participants with the 3 sleep-related disorders

| BAG | Family | Optimal k | EDF | $P_1$ |
|---|---|---|---|---|
| Reproductive_female_ProtBAG | tdist | 5 | 3.06199438 | 0.00068348 |
| **Pulmonary_ProtBAG** | **tdist** | **3** | **1.93343141** | **0.00041776** |
| Heart_ProtBAG | tdist | 3 | 1.43630571 | 0.6698409 |
| **Brain_ProtBAG** | **tdist** | **5** | **3.59501735** | **0** |
| Eye_ProtBAG | tdist | 5 | 1.00920811 | 0.00163023 |
| **Hepatic_ProtBAG** | **tdist** | **5** | **3.07745069** | **0.00298033** |
| Renal_ProtBAG | tdist | 5 | 1.00054027 | 0.93706198 |
| Reproductive_male_ProtBAG | tdist | 3 | 1.89460705 | 0.00935819 |

| | | | | |
|---|---|---|---|---|
| Endocrine_ProtBAG | tdist | 3 | 1.94760656 | 0 |
| **Immune_ProtBAG** | **tdist** | **5** | **3.48830349** | **5.00E-06** |
| **Skin_ProtBAG** | **tdist** | **5** | **3.20537479** | **6.31E-06** |
| **Endocrine_MetBAG** | **tdist** | **5** | **1.11848162** | **0.12476373** |
| Digestive_MetBAG | gamma | 5 | 1.41633043 | 0.81268972 |
| Hepatic_MetBAG | tdist | 5 | 2.74813197 | 0.00516592 |
| Immune_MetBAG | tdist | 3 | 1.01636922 | 0.00278415 |
| Metabolic_MetBAG | tdist | 5 | 1.05526167 | 0.03866805 |
| **Brain_MRIBAG** | **tdist** | **3** | **1.94357569** | **5.77E-08** |
| **Adipose_MRIBAG** | **tdist** | **3** | **1.93006499** | **0.00122662** |
| Kidney_MRIBAG | tdist | 3 | 1.81667525 | 0.04390236 |
| Heart_MRIBAG | tdist | 3 | 1.00811013 | 0.20544092 |
| Liver_MRIBAG | tdist | 5 | 2.74229668 | 0.02682607 |
| **Pancreas_MRIBAG** | **tdist** | **3** | **1.95448438** | **9.85E-06** |
| Spleen_MRIBAG | tdist | 3 | 1.00329122 | 0.49425251 |

#### f) Smoking status as an additional covariate

| BAG | Family | Optimal k | EDF | $P_1$ | Smoking P-value |
|---|---|---|---|---|---|
| Reproductive_female_ProtBAG | tdist | 5 | 3.07647389 | 0.00020392 | 1.38E-06 |
| **Pulmonary_ProtBAG** | **tdist** | **3** | **1.92565438** | **0.00156381** | **3.15E-157** |
| Heart_ProtBAG | tdist | 3 | 1.16350055 | 0.88334803 | 8.29E-15 |
| **Brain_ProtBAG** | **tdist** | **5** | **3.58453521** | **0** | **5.26E-51** |
| Eye_ProtBAG | tdist | 5 | 1.0182443 | 0.08746729 | 7.75E-17 |
| **Hepatic_ProtBAG** | **tdist** | **5** | **3.09768189** | **5.72E-05** | **2.58E-09** |
| Renal_ProtBAG | tdist | 5 | 1.00054783 | 0.71742104 | 0.00581246 |
| Reproductive_male_ProtBAG | tdist | 3 | 1.88355152 | 0 | 8.94E-43 |
| Endocrine_ProtBAG | tdist | 5 | 2.79259193 | 0.00522227 | 5.78E-45 |
| **Immune_ProtBAG** | **tdist** | **5** | **3.46806149** | **1.47E-05** | **3.89E-27** |
| **Skin_ProtBAG** | **tdist** | **3** | **1.95637863** | **3.24E-06** | **7.62E-61** |
| **Endocrine_MetBAG** | **tdist** | **5** | **1.12400433** | **0.08133393** | **2.11E-80** |
| Digestive_MetBAG | gamma | 5 | 1.87037714 | 0.11885018 | 1.09E-27 |
| Hepatic_MetBAG | tdist | 5 | 2.65050042 | 0.03504743 | 8.62E-20 |
| Immune_MetBAG | tdist | 5 | 1.14129896 | 0.0106696 | 2.09E-112 |
| Metabolic_MetBAG | tdist | 5 | 1.03560927 | 0.0318311 | 1.11E-13 |
| **Brain_MRIBAG** | **tdist** | **3** | **1.92414741** | **6.58E-06** | **1.05E-31** |
| **Adipose_MRIBAG** | **tdist** | **3** | **1.92935164** | **0.00042851** | **2.83E-18** |
| Kidney_MRIBAG | tdist | 3 | 1.86974092 | 0.02318972 | 1.88E-07 |
| Heart_MRIBAG | tdist | 3 | 1.00598814 | 0.25169447 | 2.43E-05 |
| Liver_MRIBAG | tdist | 5 | 2.54371042 | 0.06699496 | 1.23E-16 |
| **Pancreas_MRIBAG** | **tdist** | **3** | **1.95314256** | **1.27E-05** | **0.02078851** |
| Spleen_MRIBAG | tdist | 3 | 1.01337469 | 0.60022369 | 0.68429839 |

#### g) Drinking status as an additional covariate

| BAG | Family | Optimal k | EDF | $P_1$ | Drinking P-value |
|---|---|---|---|---|---|
| Brain_MRIBAG | tdist | 3 | 1.94357937 | 6.78E-07 | 0.9794196 |

#### h) Townsend deprivation index as an additional covariate

| BAG | Family | Optimal k | EDF | $P_1$ | TDI P-value |
|---|---|---|---|---|---|
| Reproductive_female_ProtBAG | tdist | 5 | 3.03173739 | 0.00176373 | 1.13E-21 |
| **Pulmonary_ProtBAG** | **tdist** | **3** | **1.92909154** | **0.0014534** | **3.12E-46** |
| Heart_ProtBAG | tdist | 3 | 1.00450041 | 0.64082859 | 5.25E-30 |
| **Brain_ProtBAG** | **tdist** | **5** | **3.57445827** | **6.16E-08** | **4.40E-42** |
| Eye_ProtBAG | tdist | 5 | 1.00866595 | 0.03456436 | 7.96E-05 |
| **Hepatic_ProtBAG** | **tdist** | **5** | **3.08802828** | **0.00231753** | **1.73E-11** |
| Renal_ProtBAG | tdist | 5 | 1.02227627 | 0.7559679 | 0.00592547 |
| Reproductive_male_ProtBAG | tdist | 3 | 1.88206974 | 0 | 1.87E-12 |
| Endocrine_ProtBAG | gaussian | 5 | 2.62155036 | 0.00025258 | 4.60E-18 |
| **Immune_ProtBAG** | **tdist** | **5** | **3.43097426** | **1.43E-05** | **4.82E-28** |
| **Skin_ProtBAG** | **tdist** | **5** | **3.13677944** | **3.55E-05** | **6.64E-12** |
| **Endocrine_MetBAG** | **tdist** | **5** | **1.05443513** | **0.07202273** | **0.0008329** |
| Digestive_MetBAG | gamma | 5 | 1.29447643 | 0.13015654 | 6.64E-07 |
| Hepatic_MetBAG | tdist | 5 | 1.82609884 | 0.00137251 | 1.23E-42 |
| Immune_MetBAG | tdist | 3 | 1.00542948 | 0.00381486 | 1.90E-57 |
| Metabolic_MetBAG | tdist | 5 | 1.03798729 | 0.026891 | 0.02246045 |
| **Brain_MRIBAG** | **tdist** | **3** | **1.9369181** | **7.63E-07** | **8.12E-09** |
| **Adipose_MRIBAG** | **tdist** | **3** | **1.92548561** | **0.00058181** | **0.02314232** |
| Kidney_MRIBAG | tdist | 3 | 1.8572008 | 0.04731968 | 0.59550499 |
| Heart_MRIBAG | tdist | 3 | 1.00113383 | 0.86635344 | 0.23855493 |
| Liver_MRIBAG | tdist | 5 | 2.63688227 | 0.03493911 | 0.00014025 |
| **Pancreas_MRIBAG** | **tdist** | **3** | **1.95213973** | **1.62E-05** | **0.11460292** |
| Spleen_MRIBAG | tdist | 3 | 1.00480559 | 0.7863594 | 0.41165241 |

#### i) Computer use time as an additional covariate

| BAG | Family | Optimal k | EDF | $P_1$ | Computer P-value |
|---|---|---|---|---|---|
| Reproductive_female_ProtBAG | tdist | 5 | 3.18944764 | 0.00067925 | 0.74821762 |
| **Pulmonary_ProtBAG** | **tdist** | **5** | **3.22474103** | **0.00019265** | **0.00544064** |
| Heart_ProtBAG | tdist | 3 | 1.50862194 | 0.62910784 | 0.09686478 |
| Brain_ProtBAG | tdist | 5 | 3.61909242 | 0 | 0.01987495 |
| Eye_ProtBAG | tdist | 5 | 1.00911381 | 0.02520375 | 0.5545165 |
| **Hepatic_ProtBAG** | **tdist** | **5** | **3.18615336** | **0.00061533** | **0.88429267** |
| Renal_ProtBAG | tdist | 5 | 1.00054743 | 0.65898516 | 0.49449189 |
| Reproductive_male_ProtBAG | tdist | 3 | 1.88764293 | 0 | 0.71022606 |
| Endocrine_ProtBAG | tdist | 5 | 2.84427693 | 0 | 0.79565575 |
| **Immune_ProtBAG** | **tdist** | **5** | **3.46868396** | **1.62E-05** | **0.85143241** |
| **Skin_ProtBAG** | **tdist** | **5** | **3.24114512** | **0** | **0.03997704** |
| **Endocrine_MetBAG** | **tdist** | **5** | **1.20433165** | **0.22905252** | **6.30E-05** |
| Digestive_MetBAG | gamma | 5 | 1.94910956 | 0.53611163 | 0.00018247 |
| Hepatic_MetBAG | tdist | 5 | 2.56076449 | 0.04778857 | 6.40E-07 |
| Immune_MetBAG | tdist | 5 | 1.32332746 | 0.09150513 | 0.07623785 |
| Metabolic_MetBAG | tdist | 5 | 1.03429731 | 0.00107858 | 1.09E-22 |

| | | | | | |
|---|---|---|---|---|---|
| **Brain_MRIBAG** | **tdist** | **3** | **1.95218679** | **5.26E-07** | **0.43210818** |
| **Adipose_MRIBAG** | **tdist** | **3** | **1.93069926** | **0.00040982** | **0.00149075** |
| Kidney_MRIBAG | tdist | 3 | 1.86228808 | 0.04389015 | 0.00020942 |
| Heart_MRIBAG | tdist | 3 | 1.00153078 | 0.18121213 | 0.00096716 |
| Liver_MRIBAG | tdist | 5 | 2.43983282 | 0.05015158 | 4.23E-05 |
| **Pancreas_MRIBAG** | **tdist** | **3** | **1.95303053** | **2.20E-05** | **0.01033234** |
| Spleen_MRIBAG | tdist | 3 | 1.00459786 | 0.84805194 | 0.0025427 |

#### j) Testosterone as an additional covariate

| BAG | Family | Optimal k | EDF | $P_1$ | Testosterone-sex P-value |
|---|---|---|---|---|---|
| Reproductive_female_ProtBAG | tdist | 5 | 2.84226889 | 0.00046516 | 0.02277355 |
| **Pulmonary_ProtBAG** | **tdist** | **5** | **3.0433898** | **0.00113916** | **0.56362198** |
| Heart_ProtBAG | tdist | 3 | 1.01709612 | 0.12902099 | 0.08104452 |
| Brain_ProtBAG | tdist | 5 | 3.53915248 | 6.28E-07 | 0.69955614 |
| Eye_ProtBAG | tdist | 5 | 1.01560282 | 0.48292884 | 0.20427132 |
| **Hepatic_ProtBAG** | **tdist** | **5** | **3.08174128** | **0.0008631** | **0.60014667** |
| Renal_ProtBAG | tdist | 5 | 1.00153944 | 0.7516835 | 0.0252769 |
| Reproductive_male_ProtBAG | tdist | 3 | 1.65681854 | 0.10800177 | 0.21850358 |
| Endocrine_ProtBAG | tdist | 5 | 2.80241613 | 1.49E-06 | 0.03290308 |
| **Immune_ProtBAG** | **tdist** | **5** | **2.78996734** | **0.03230766** | **0.4342481** |
| **Skin_ProtBAG** | **tdist** | **5** | **3.18241734** | **1.82E-05** | **0.31569186** |
| **Endocrine_MetBAG** | **tdist** | **5** | **3.52594116** | **0** | **0.17216201** |
| Digestive_MetBAG | gamma | 5 | 1.20767056 | 0.67224535 | 0.37888247 |
| Hepatic_MetBAG | tdist | 5 | 2.62979272 | 0.12726215 | 0.61544352 |
| Immune_MetBAG | tdist | 3 | 1.02801117 | 0.30731306 | 0.01430927 |
| Metabolic_MetBAG | tdist | 5 | 1.05727051 | 0.1803341 | 0.07914344 |
| **Brain_MRIBAG** | **tdist** | **3** | **1.94272223** | **3.55E-06** | **0.42789179** |
| **Adipose_MRIBAG** | **tdist** | **3** | **1.92812902** | **0.00104041** | **0.94711766** |
| Kidney_MRIBAG | tdist | 3 | 1.94279662 | 0.00041159 | 0.36445145 |
| Heart_MRIBAG | tdist | 3 | 1.00557206 | 0.39871486 | 0.25835639 |
| Liver_MRIBAG | tdist | 5 | 2.75106161 | 0.08235739 | 0.94124085 |
| **Pancreas_MRIBAG** | **tdist** | **3** | **1.9475421** | **0.00012862** | **0.06519536** |
| Spleen_MRIBAG | tdist | 3 | 1.00197394 | 0.54937148 | 0.51936839 |

#### k) SHBG use time as an additional covariate

| BAG | Family | Optimal k | EDF | $P_1$ | SHBG-sex P-value |
|---|---|---|---|---|---|
| Reproductive_female_ProtBAG | tdist | 5 | 3.01870211 | 0.03401377 | 0.05484021 |
| **Pulmonary_ProtBAG** | **tdist** | **5** | **3.08981744** | **0.01059715** | **0.1126294** |
| Heart_ProtBAG | tdist | 3 | 1.49161172 | 0.61211574 | 0.77058205 |
| **Brain_ProtBAG** | **tdist** | **5** | **3.49913133** | **0** | **0.5581973** |
| Eye_ProtBAG | tdist | 5 | 1.0103298 | 0.00059469 | 0.6029346 |
| **Hepatic_ProtBAG** | **tdist** | **5** | **3.11971439** | **0.00147547** | **0.20173444** |
| Renal_ProtBAG | tdist | 5 | 1.09991854 | 0.7397906 | 0.68639925 |
| Reproductive_male_ProtBAG | tdist | 3 | 1.89352713 | 5.81E-07 | 0.45821297 |
| Endocrine_ProtBAG | tdist | 5 | 2.75622067 | 0.00687666 | 0.05384894 |
| **Immune_ProtBAG** | **tdist** | **5** | **3.43253247** | **7.45E-05** | **0.05227508** |
| **Skin_ProtBAG** | **tdist** | **5** | **3.18867577** | **2.38E-06** | **0.45275612** |
| **Endocrine_MetBAG** | **tdist** | **5** | **1.04511838** | **0.44538986** | **0.22282905** |
| Digestive_MetBAG | gamma | 5 | 2.75642165 | 0.2497407 | 0.16843459 |

| | | | | | |
|---|---|---|---|---|---|
| Hepatic_MetBAG | tdist | 5 | 2.09930704 | 0.25170464 | 0.35219559 |
| Immune_MetBAG | tdist | 3 | 1.00954793 | 0.1021914 | 0.00714115 |
| Metabolic_MetBAG | tdist | 5 | 1.08186155 | 0.06099562 | 0.59147502 |
| **Brain_MRIBAG** | **tdist** | **3** | **1.93187785** | **1.59E-06** | **0.76742474** |
| **Adipose_MRIBAG** | **tdist** | **3** | **1.93135174** | **0.00065938** | **0.45277903** |
| Kidney_MRIBAG | tdist | 3 | 1.74368015 | 0.06400277 | 0.74039525 |
| Heart_MRIBAG | tdist | 3 | 1.00579189 | 0.054327 | 0.64550715 |
| Liver_MRIBAG | tdist | 5 | 2.69601661 | 0.02442577 | 0.12881184 |
| **Pancreas_MRIBAG** | **tdist** | **3** | **1.95187447** | **3.67E-06** | **0.85898724** |
| Spleen_MRIBAG | tdist | 3 | 1.00217496 | 0.13323092 | 0.7381724 |

##### l) Estradiol use time as an additional covariate

Of note, the sample sizes reduced dramatically compared to **Fig. 1**, because not all the full UKBB samples have data available for estradiol (i.e., UKBB only covers 77,594 participants out of the 500k population before merging with the BAG populations: https://biobank.ndph.ox.ac.uk/ukb/field.cgi?id=30800).

| BAG | Family | Optimal k | EDF | $P_1$ | Estradiol-sex P-value |
|---|---|---|---|---|---|
| Reproductive_female_ProtBAG | tdist | 5 | 1.00087574 | 0.52593908 | 0.33062635 |
| **Pulmonary_ProtBAG** | **tdist** | **5** | **2.97862243** | **0.02639053** | **0.27371533** |
| Heart_ProtBAG | tdist | 3 | 1.00042744 | 0.0850369 | 0.85407943 |
| **Brain_ProtBAG** | **tdist** | **5** | **2.3158273** | **0.18105885** | **0.0911572** |
| Eye_ProtBAG | tdist | 5 | 1.00134332 | 0.75437986 | 0.27806343 |
| **Hepatic_ProtBAG** | **tdist** | **5** | **1.00067089** | **0.1629343** | **0.01958318** |
| Renal_ProtBAG | tdist | 5 | 1.0013687 | 0.81846298 | 0.04770111 |
| Reproductive_male_ProtBAG | tdist | 3 | 1.41017194 | 0.69433227 | 0.39839317 |
| Endocrine_ProtBAG | tdist | 5 | 2.74968973 | 0.04483631 | 0.23368963 |
| **Immune_ProtBAG** | **tdist** | **5** | **1.82752125** | **0.89679547** | **0.40191012** |
| **Skin_ProtBAG** | **tdist** | **5** | **2.33726922** | **0.07899631** | **0.06714145** |
| **Endocrine_MetBAG** | **tdist** | **5** | **1.91030582** | **0.37967898** | **0.43184208** |
| Digestive_MetBAG | gamma | 5 | 1.95075084 | 0.25088444 | 0.39171171 |
| Hepatic_MetBAG | tdist | 5 | 1.00426347 | 0.64147517 | 0.62064032 |
| Immune_MetBAG | tdist | 3 | 1.00663804 | 0.70033281 | 0.91679333 |
| Metabolic_MetBAG | tdist | 5 | 1.00846985 | 0.78364727 | 0.48745155 |
| **Brain_MRIBAG** | **tdist** | **3** | **1.7138767** | **0.25726918** | **0.94053148** |
| **Adipose_MRIBAG** | **tdist** | **3** | **1.8343143** | **0.08377288** | **0.21982963** |
| Kidney_MRIBAG | tdist | 3 | 1.52938374 | 0.56890008 | 0.94408945 |
| Heart_MRIBAG | tdist | 3 | 1.00089616 | 0.79391252 | 0.44827056 |
| Liver_MRIBAG | tdist | 5 | 1.00032981 | 0.17315766 | 0.79358438 |
| **Pancreas_MRIBAG** | **tdist** | **3** | **1.00134533** | **0.01147831** | **0.96429587** |
| Spleen_MRIBAG | tdist | 3 | 1.00027543 | 0.4406184 | 0.9974435 |

##### m) GAMLSS analysis for the 9 significant sleep-BAG relationships

| BAG | Family | $P_1$ | $P_2$ | $P_3$ |
|---|---|---|---|---|
| Reproductive_female_ProtBAG | TF | 6.10E-14 | 0 | 1 |
| **Pulmonary_ProtBAG** | **TF** | **3.22E-18** | **0** | **1** |
| Heart_ProtBAG | TF | 3.01E-09 | 0 | 1 |
| **Brain_ProtBAG** | **TF** | **1.77E-23** | **0** | **1** |
| Eye_ProtBAG | TF | 1.48E-06 | 0 | 0.30910717 |
| Hepatic_ProtBAG | TF | 1.35E-08 | 0 | 1 |
| Renal_ProtBAG | TF | 0.25631225 | 0 | 1 |
| Reproductive_male_ProtBAG | TF | 8.83E-17 | 2.58E-11 | 1 |

| | | | | |
|---|---|---|---|---|
| Endocrine_ProtBAG | TF | 4.29E-15 | 0 | 1 |
| **Immune_ProtBAG** | **TF** | **5.60E-11** | **0** | **1** |
| **Skin_ProtBAG** | **TF** | **4.00E-08** | **2.81E-11** | **1** |
| **Endocrine_MetBAG** | **TF** | **5.47E-36** | **0** | **1** |
| Digestive_MetBAG | gamma | 0.00484392 | 0 | 1 |
| Hepatic_MetBAG | TF | 0.00536906 | 0 | 1 |
| Immune_MetBAG | TF | 0.45163538 | 0 | 0.52623149 |
| Metabolic_MetBAG | TF | 2.18E-05 | 0 | 1 |
| **Brain_MRIBAG** | **TF** | **2.50E-10** | **2.46E-13** | **1** |
| **Adipose_MRIBAG** | **TF** | **1.40E-15** | **0.06256714** | **1** |
| Kidney_MRIBAG | TF | 7.49E-07 | 0 | 1 |
| Heart_MRIBAG | TF | 0.30835973 | 0 | 0.16592368 |
| Liver_MRIBAG | TF | 2.48E-05 | 2.11E-05 | 1 |
| **Pancreas_MRIBAG** | **TF** | **4.17E-06** | **8.24E-12** | **1** |
| Spleen_MRIBAG | TF | 0.69124192 | 4.31E-09 | 0.13406013 |

**Supplementary Table 4: Genomic loci identified in our GWAS of the abnormal sleep duration patterns**

Using sleep durations as a binary trait for Short vs. Normal sleep duration and Long vs. Normal sleep duration, we identified genomics loci linked to these patterns using REGENIE. Genome-wide P-value threshold was used ($P<5x10^{-8}$).

| Top Lead SNP | Chromosome | Position | P-value | Phenotype | Cytogenetic region |
|---|---|---|---|---|---|
| rs34382732 | 3 | 193108140 | 5.23E-09 | short_vs_normal | 3q29 |
| rs62118577 | 2 | 9266185 | 4.80E-08 | long_vs_normal | 2p25.1 |
| rs74889896 | 2 | 58933235 | 8.40E-09 | long_vs_normal | 2p16.1 |
| rs62158206 | 2 | 114084596 | 1.81E-11 | long_vs_normal | 2q13 |
| 11:76414249_GT_G | 11 | 76414249 | 1.75E-09 | long_vs_normal | 11q13.5 |
| rs75458655 | 11 | 118115331 | 9.85E-10 | long_vs_normal | 11q23.3 |
| rs1057703 | 11 | 122830251 | 2.21E-08 | long_vs_normal | 11q24.1 |
| rs62061734 | 17 | 44018488 | 1.01E-13 | long_vs_normal | 17q21.31 |

**Supplementary Table 5: Genetic correlation between short/long sleep duration and 23 BAGs**

Using sleep durations as a binary trait for short/long vs. normal sleep duration, we identified significant signals (bold text; P-value<0.05/23). NC indicated the LDSC model did not converge.

| Sleep type | BAG | BAG_type | gc_mean | gc_std | Z | P |
|---|---|---|---|---|---|---|
| short_vs_normal | Reproductive_female | ProtBAG | 0.1131 | 0.128 | 0.883 | 0.3772 |
| short_vs_normal | Pulmonary | ProtBAG | 0.013 | 0.0602 | 0.2162 | 0.8288 |
| short_vs_normal | Heart | ProtBAG | 0.0094 | 0.0883 | 0.1061 | 0.9155 |
| short_vs_normal | Brain | ProtBAG | 0.0186 | 0.0763 | 0.2443 | 0.807 |
| short_vs_normal | Eye | ProtBAG | NC | NC | NC | NC |
| short_vs_normal | Hepatic | ProtBAG | 0.1863 | 0.0764 | 2.439 | 0.0147 |
| short_vs_normal | Renal | ProtBAG | -0.0007 | 0.0585 | -0.0112 | 0.9911 |
| short_vs_normal | Reproductive_male | ProtBAG | 0.0776 | 0.0613 | 1.2653 | 0.2058 |
| short_vs_normal | Endocrine | ProtBAG | 0.0111 | 0.0802 | 0.1383 | 0.89 |
| short_vs_normal | Immune | ProtBAG | 0.0344 | 0.0546 | 0.6295 | 0.529 |
| short_vs_normal | Skin | ProtBAG | 0.017 | 0.0579 | 0.294 | 0.7688 |
| **short_vs_normal** | **Endocrine** | **MetBAG** | **0.1442** | **0.0394** | **3.6582** | **0.0003** |
| short_vs_normal | Digestive | MetBAG | -0.0826 | 0.048 | -1.7224 | 0.085 |
| short_vs_normal | Hepatic | MetBAG | 0.0841 | 0.0427 | 1.9725 | 0.0485 |
| short_vs_normal | Immune | MetBAG | 0.0099 | 0.0447 | 0.2213 | 0.8249 |
| short_vs_normal | Metabolic | MetBAG | 0.0121 | 0.0494 | 0.2445 | 0.8068 |
| short_vs_normal | brain | MRIBAG | 0.0239 | 0.0542 | 0.4404 | 0.6596 |
| short_vs_normal | adipose | MRIBAG | 0.1948 | 0.0759 | 2.5673 | 0.0102 |
| short_vs_normal | heart | MRIBAG | 0.107 | 0.1 | 1.0693 | 0.2849 |
| short_vs_normal | kidney | MRIBAG | 0.0944 | 0.0654 | 1.4447 | 0.1486 |
| short_vs_normal | liver | MRIBAG | 0.0508 | 0.075 | 0.6767 | 0.4986 |
| short_vs_normal | pancreas | MRIBAG | -0.0365 | 0.0668 | -0.5459 | 0.5851 |
| short_vs_normal | spleen | MRIBAG | -0.1161 | 0.094 | -1.2348 | 0.2169 |
| long_vs_normal | Reproductive_female | ProtBAG | 0.138 | 0.1121 | 1.2307 | 0.2184 |
| long_vs_normal | Pulmonary | ProtBAG | 0.0081 | 0.0611 | 0.132 | 0.895 |
| long_vs_normal | Heart | ProtBAG | 0.1566 | 0.0852 | 1.8389 | 0.0659 |
| long_vs_normal | Brain | ProtBAG | -0.0886 | 0.074 | -1.1974 | 0.2311 |
| long_vs_normal | Eye | ProtBAG | NC | NC | NC | NC |
| long_vs_normal | Hepatic | ProtBAG | 0.0183 | 0.0771 | 0.2371 | 0.8126 |
| long_vs_normal | Renal | ProtBAG | 0.1303 | 0.0592 | 2.2001 | 0.0278 |
| long_vs_normal | Reproductive_male | ProtBAG | 0.0822 | 0.0672 | 1.2231 | 0.2213 |
| long_vs_normal | Endocrine | ProtBAG | 0.1031 | 0.0765 | 1.3475 | 0.1778 |
| long_vs_normal | Immune | ProtBAG | 0.0468 | 0.0531 | 0.8822 | 0.3777 |
| long_vs_normal | Skin | ProtBAG | -0.0762 | 0.0573 | -1.3294 | 0.1837 |
| long_vs_normal | Endocrine | MetBAG | 0.0391 | 0.039 | 1.0038 | 0.3155 |
| long_vs_normal | Digestive | MetBAG | -0.0563 | 0.0493 | -1.1415 | 0.2537 |
| long_vs_normal | Hepatic | MetBAG | -0.0165 | 0.0396 | -0.4173 | 0.6765 |
| long_vs_normal | Immune | MetBAG | 0.0102 | 0.048 | 0.2134 | 0.831 |
| long_vs_normal | Metabolic | MetBAG | -0.1108 | 0.0507 | -2.1847 | 0.0289 |
| long_vs_normal | brain | MRIBAG | 0.0723 | 0.0574 | 1.2594 | 0.2079 |
| long_vs_normal | adipose | MRIBAG | 0.1803 | 0.076 | 2.3739 | 0.0176 |
| long_vs_normal | heart | MRIBAG | 0.0024 | 0.0739 | 0.0322 | 0.9743 |
| long_vs_normal | kidney | MRIBAG | 0.1227 | 0.0575 | 2.1317 | 0.033 |
| long_vs_normal | liver | MRIBAG | 0.079 | 0.0753 | 1.0503 | 0.2936 |
| **long_vs_normal** | **pancreas** | **MRIBAG** | **0.2079** | **0.0663** | **3.1372** | **0.0017** |
| long_vs_normal | spleen | MRIBAG | 0.0634 | 0.0817 | 0.7763 | 0.4376 |

**Supplementary Table 6: Sensitivity analysis for our mediation analysis using brain MRI as the mediator**

In the mediation analysis framework, *a1* and *c1* represent the indirect pathways' coefficients, capturing the exposure's effect on the outcome that operates through intermediate variables (mediators). In contrast, *c2* denotes the coefficient of the direct pathway, quantifying the portion of the exposure's effect on the outcome that is not mediated. The variable prop_mediated reflects the proportion of the total effect that is explained by the indirect pathways, offering insight into how much of the exposure's influence on the outcome is transmitted through mediators rather than occurring directly.

**a**) The results of the forward mediational analysis: sleep → brain MRIBAG → LLD1/2.

| Outcome | Exposure | Mediator | label | EST | SE | Pvalue | CI.lower | CI.upper |
|---|---|---|---|---|---|---|---|---|
| LLD_1 | short | Brain_MRIBAG | c1 | -0.1159724 | 0.00374705 | 0 | -0.1234494 | -0.1087631 |
| LLD_1 | short | Brain_MRIBAG | c2 | -0.2929474 | 0.06010315 | 1.09E-06 | -0.4121581 | -0.1781465 |
| LLD_1 | short | Brain_MRIBAG | a1 | 0.07285311 | 0.10015028 | 0.46695777 | -0.1288059 | 0.27273862 |
| LLD_1 | short | Brain_MRIBAG | indirect | -0.0084489 | NA | NA | NA | NA |
| LLD_1 | short | Brain_MRIBAG | total | -0.3013963 | NA | NA | NA | NA |
| LLD_1 | short | Brain_MRIBAG | prop_mediated | **0.02803268** | NA | NA | NA | NA |
| LLD_1 | long | Brain_MRIBAG | c1 | -0.1133255 | 0.00362608 | 0 | -0.1203902 | -0.1059577 |
| LLD_1 | long | Brain_MRIBAG | c2 | -0.0166764 | 0.05004374 | 0.73895606 | -0.1127656 | 0.0847863 |
| LLD_1 | long | Brain_MRIBAG | a1 | 0.54340736 | 0.07821339 | 3.71E-12 | 0.38809163 | 0.69511799 |
| LLD_1 | long | Brain_MRIBAG | indirect | -0.0615819 | NA | NA | NA | NA |
| LLD_1 | long | Brain_MRIBAG | total | -0.0782583 | NA | NA | NA | NA |
| LLD_1 | long | Brain_MRIBAG | prop_mediated | **0.78690595** | NA | NA | NA | NA |
| LLD_2 | short | Brain_MRIBAG | c1 | 0.10853972 | 0.00278399 | 0 | 0.1027339 | 0.11385597 |
| LLD_2 | short | Brain_MRIBAG | c2 | 0.23816434 | 0.04826107 | 8.02E-07 | 0.14301172 | 0.33321658 |
| LLD_2 | short | Brain_MRIBAG | a1 | 0.07285311 | 0.1005749 | 0.46884064 | -0.1308664 | 0.27253572 |
| LLD_2 | short | Brain_MRIBAG | indirect | 0.00790746 | NA | NA | NA | NA |
| LLD_2 | short | Brain_MRIBAG | total | **0.2460718** | NA | NA | NA | NA |
| LLD_2 | short | Brain_MRIBAG | prop_mediated | 0.03213475 | NA | NA | NA | NA |
| LLD_2 | long | Brain_MRIBAG | c1 | 0.10719328 | 0.00274408 | 0 | 0.10184037 | 0.11248424 |
| LLD_2 | long | Brain_MRIBAG | c2 | 0.03422376 | 0.03930302 | 0.38388161 | -0.0418017 | 0.10925456 |
| LLD_2 | long | Brain_MRIBAG | a1 | 0.54340736 | 0.07861616 | 4.77E-12 | 0.38747366 | 0.69499392 |
| LLD_2 | long | Brain_MRIBAG | indirect | 0.05824962 | NA | NA | NA | NA |
| LLD_2 | long | Brain_MRIBAG | total | 0.09247337 | NA | NA | NA | NA |
| LLD_2 | long | Brain_MRIBAG | prop_mediated | **0.6299069** | NA | NA | NA | NA |

**b**) The results of the forward mediational analysis: sleep → LLD1/2 → brain MRIBAG. Unreasonable prop_mediated value > 1 is highlighted to support the potential violation of the timing-order underlying the data presented in this analysis.

| Mediator | Outcome | Exposure | Label | EST | SE | Pvalue | CI.lower | CI.upper |
|---|---|---|---|---|---|---|---|---|
| LLD_1 | Brain_MRIBAG | short | c1 | -0.2649692 | 0.00852904 | 0 | -0.2813043 | -0.2485956 |
| LLD_1 | Brain_MRIBAG | short | c2 | -0.0070076 | 0.09757933 | 0.94274941 | -0.1956114 | 0.18025664 |
| LLD_1 | Brain_MRIBAG | short | a1 | -0.3013963 | 0.06307823 | 1.77E-06 | -0.4288353 | -0.1800933 |
| LLD_1 | Brain_MRIBAG | short | indirect | 0.07986073 | NA | NA | NA | NA |
| LLD_1 | Brain_MRIBAG | short | total | 0.07285311 | NA | NA | NA | NA |
| LLD_1 | Brain_MRIBAG | short | prop_mediated | **1.09618843** | NA | NA | NA | NA |
| LLD_1 | Brain_MRIBAG | long | c1 | -0.260579 | 0.00844469 | 0 | -0.27672 | -0.2433674 |
| LLD_1 | Brain_MRIBAG | long | c2 | 0.52301489 | 0.08037933 | 7.68E-11 | 0.36693716 | 0.68122753 |
| LLD_1 | Brain_MRIBAG | long | a1 | -0.0782583 | 0.05124766 | 0.12674575 | -0.1770108 | 0.01915737 |
| LLD_1 | Brain_MRIBAG | long | indirect | 0.02039247 | NA | NA | NA | NA |
| LLD_1 | Brain_MRIBAG | long | total | 0.54340736 | NA | NA | NA | NA |
| LLD_1 | Brain_MRIBAG | long | prop_mediated | 0.03752705 | NA | NA | NA | NA |
| LLD_2 | Brain_MRIBAG | short | c1 | 0.42961069 | 0.01064015 | 0 | 0.4085734 | 0.45158347 |
| LLD_2 | Brain_MRIBAG | short | c2 | -0.032862 | 0.09725948 | 0.73545411 | -0.2268089 | 0.15327048 |
| LLD_2 | Brain_MRIBAG | short | a1 | 0.2460718 | 0.05076844 | 1.25E-06 | 0.14794822 | 0.34476181 |
| LLD_2 | Brain_MRIBAG | short | indirect | 0.10571508 | NA | NA | NA | NA |
| LLD_2 | Brain_MRIBAG | short | total | 0.07285311 | NA | NA | NA | NA |
| LLD_2 | Brain_MRIBAG | short | prop_mediated | **1.45107162** | NA | NA | NA | NA |
| LLD_2 | Brain_MRIBAG | long | c1 | 0.42576711 | 0.01061222 | 0 | 0.40287709 | 0.44528729 |
| LLD_2 | Brain_MRIBAG | long | c2 | 0.50403524 | 0.07977 | 2.64E-10 | 0.34946602 | 0.65886541 |
| LLD_2 | Brain_MRIBAG | long | a1 | 0.09247337 | 0.03889352 | 0.01742556 | 0.01569276 | 0.16699961 |
| LLD_2 | Brain_MRIBAG | long | indirect | 0.03937212 | NA | NA | NA | NA |
| LLD_2 | Brain_MRIBAG | long | total | 0.54340736 | NA | NA | NA | NA |

| | | | | | | | | |
|---|---|---|---|---|---|---|---|---|
| LLD_2 | Brain_MRIBAG | long | prop_mediated | 0.07245415 | NA | NA | NA | NA |

**Supplementary Table 7: 521 FinnGen and 6 PGC GWAS summary statistics**

We used summary statistics from 6 PGC GWAS in all genetic correlation analyses, two of which lacked allele frequency information and were therefore excluded from the Mendelian randomization analysis.

**PGC**:

| Author | DE | Download URL | PMID | Build # | Ancestry |
|---|---|---|---|---|---|
| Ditte Demontis | ADHD | https://figshare.com/articles/dataset/adhd2022/22564390 | 36702997 | GRCh37 | European |
| Douglas P. Wightman | AD | https://ctg.cncr.nl/software/summary_statistics | 34493870 | GRCh37 | European |
| Jakob Grove | ASD | https://figshare.com/articles/dataset/asd2019/14671989 | 30804558 | GRCh37 | European |
| Niamh Mullins | BIP | https://figshare.com/articles/dataset/PGC3_bipolar_disorder_GWAS_summary_statistics/14102594 | 34002096 | GRCh37 | European |
| OCGAS | OCD | https://figshare.com/articles/dataset/ocd2018/14672103 | 28761083 | GRCh37 | European |
| Vassily Trubetskoy | SCZ | https://figshare.com/articles/dataset/scz2022/19426775 | 35396580 | GRCh37 | European |

**FinnGen:**

| DE | Full disease name | Category | N-cases | N-controls |
|---|---|---|---|---|
| RX_PARACETAMOL_NSAID | Paracetamol of NSAID medication | Drug purchase endpoints | 314977 | 62300 |
| RX_ANTIHYP | Antihypertensive medication - note that there are other indications | Drug purchase endpoints | 199546 | 177731 |
| FG_CVD | Cardiovascular diseases (excluding rheumatic etc) | Cardiometabolic endpoints | 185353 | 191924 |
| PAIN | Pain (limb, back, neck, head abdominally) | Miscellaneous, not yet classified endpoints | 171922 | 204598 |
| K11_CARIES_1_ONLYAVO | Dental caries 1, only avohilmo | XI Diseases of the digestive system (K11_) | 161113 | 216164 |
| M13_ARTHROPATHIES | Arthropathies | XIII Diseases of the musculoskeletal system and connective tissue (M13_) | 136415 | 240862 |
| RX_STATIN | Statin medication | IX Diseases of the circulatory system (I9_) | 127169 | 250108 |
| K11_ORAL | Diseases of oral cavity, salivary glands and jaws | XI Diseases of the digestive system (K11_) | 118043 | 259234 |
| PULM_INFECTIONS | COPD/asthma/ILD related infections | Interstitial lung disease endpoints | 117312 | 259965 |
| I9_HYPTENS | Hypertension | IX Diseases of the circulatory system (I9_) | 111581 | 265626 |
| ANTIDEPRESSANTS | Depression medications | Comorbidities of Neurological endpoints | 106785 | 88536 |
| O15_DELIV_SPONT | Single spontaneous delivery | XV Pregnancy, childbirth and the puerperium (O15_) | 106627 | 92306 |
| M13_DORSOPATHY | Dorsopathies | XIII Diseases of the musculoskeletal system and connective tissue (M13_) | 106313 | 270964 |
| N14_FEMALEGENNONINF | Noninflammatory disorders of female genital tract | XIV Diseases of the genitourinary system (N14_) | 103306 | 107564 |
| M13_SOFTTISSUE | Soft tissue disorders | XIII Diseases of the musculoskeletal system and connective tissue (M13_) | 102065 | 275212 |
| KRA_PSY_ANYMENTAL | Any mental disorder | Psychiatric endpoints from Katri R√§ikk√∂nen | 99751 | 277526 |
| M13_ARTHROSIS_INCLAVO | Artrosis, including avohilmo | XIII Diseases of the musculoskeletal system and connective tissue (M13_) | 96500 | 280777 |
| AUTOIMMUNE | Autoimmune diseases | Diseases marked as autimmune origin | 96150 | 281127 |
| J10_UPPERDIS | Other diseases of upper respiratory tract | X Diseases of the respiratory system (J10_) | 93935 | 283342 |
| FALLS | Falls/tendency to fall | Miscellaneous, not yet classified endpoints | 92857 | 284420 |

| | | | | |
|---|---|---|---|---|
| I9_HYPTENSESS | Hypertension, essential | IX Diseases of the circulatory system (I9_) | 92462 | 265626 |
| G6_EPIPAROX | Episodal and paroxysmal disorders | VI Diseases of the nervous system (G6_) | 89440 | 287837 |
| K11_GINGIVITIS_PERIODONTAL | Gingivitis and periodontal diseases | XI Diseases of the digestive system (K11_) | 87497 | 259234 |
| K11_PULPITIS_1_ONLYAVO | Dental pulpitis 1, only avohilmo | XI Diseases of the digestive system (K11_) | 81713 | 295564 |
| M13_OTHERJOINT | Other joint disorders | XIII Diseases of the musculoskeletal system and connective tissue (M13_) | 81676 | 240862 |
| M13_DORSOPATHYOTH | Other dorsopathies, not elsewhere classified | XIII Diseases of the musculoskeletal system and connective tissue (M13_) | 79212 | 270964 |
| FG_OTHHEART | Other heart diseases | Cardiometabolic endpoints | 77711 | 191924 |
| RX_CROHN_1STLINE | First line medication for Crohn's disease | Drug purchase endpoints | 77497 | 299780 |
| C3_CANCER_EXALLC | Malignant neoplasm (controls excluding all cancers) | II Neoplasms, from cancer register (ICD-O-3) | 77325 | 287137 |
| K11_INTESTOTH | Other diseases of intestines | XI Diseases of the digestive system (K11_) | 75346 | 301931 |
| M13_ARTHROSIS | Arthrosis | XIII Diseases of the musculoskeletal system and connective tissue (M13_) | 73710 | 240862 |
| J10_UPPERINFEC | Acute upper respiratory infections | X Diseases of the respiratory system (J10_) | 69111 | 308166 |
| CARDIAC_ARRHYTM | Cardiac arrhythmias | Comorbidities of COPD | 67035 | 220224 |
| J10_LOWCHRON | Chronic lower respiratory diseases | X Diseases of the respiratory system (J10_) | 65991 | 311286 |
| RX_N05C | Use of hypnotics and sedatives | V Mental and behavioural disorders (F5_) | 65814 | 311463 |
| I9_IHD | Ischaemic heart disease, wide definition | IX Diseases of the circulatory system (I9_) | 63744 | 313533 |
| J10_INFLUPNEU | Influenza and pneumonia | X Diseases of the respiratory system (J10_) | 62604 | 314673 |
| O15_PREG_ABORT | Pregnancy with abortive outcome | XV Pregnancy, childbirth and the puerperium (O15_) | 61248 | 149622 |
| I9_CVD_HARD | Hard cardiovascular diseases | Cardiometabolic endpoints | 61240 | 316037 |
| H7_CATARACTSENILE | Senile cataract | VII Diseases of the eye and adnexa (H7_) | 59522 | 312864 |
| M13_DORSALGIA | Dorsalgia | XIII Diseases of the musculoskeletal system and connective tissue (M13_) | 59438 | 270964 |
| J10_PNEUMONIA | All pneumoniae | X Diseases of the respiratory system (J10_) | 58174 | 319103 |
| RX_CODEINE_TRAMADOL | Codeine or tramadol medication | Drug purchase endpoints | 57823 | 319454 |
| T2D | Type 2 diabetes, definitions combined | Diabetes endpoints | 57698 | 308252 |
| K11_OESSTODUO | Diseases of oesophagus, stomach and duodenum | XI Diseases of the digestive system (K11_) | 56890 | 320387 |
| E4_THYROID | Disorders of the thyroid gland | IV Endocrine, nutritional and metabolic diseases (E4_) | 56574 | 320703 |
| BRONCHITIS | Bronchitis | Asthma and related endpoints | 55222 | 322055 |
| K11_HERNIA | Hernia | XI Diseases of the digestive system (K11_) | 54691 | 322586 |
| KELA_DIAB_INSUL_EXMORE | Diabetes, insulin treatment (Kela reimbursement) (more control exclusions) | Diabetes endpoints | 53892 | 308249 |
| AUTOIMMUNE_NONTHYROID | Autoimmune diseases excluding thyroid diseases | Diseases marked as autimmune origin | 53213 | 280251 |

| | | | | |
|---|---|---|---|---|
| I9_DISVEINLYMPH | Diseases of veins, lymphatic vessels and lymph nodes, not elsewhere classified | IX Diseases of the circulatory system (I9_) | 53156 | 324121 |
| E4_METABOLIA | Metabolic disorders | IV Endocrine, nutritional and metabolic diseases (E4_) | 53127 | 324150 |
| AUTOIMMUNE_NONTHYROID_STRICT | Autoimmune diseases excluding thyroid diseases, strict definition | Diseases marked as autimmune origin | 51863 | 275890 |
| N14_URINOTH | Other diseases of urinary system | XIV Diseases of the genitourinary system (N14_) | 49276 | 328001 |
| F5_DEPRESSION_DYSTHYMIA | Depression or dysthymia | V Mental and behavioural disorders (F5_) | 48847 | 225483 |
| O15_COMPLIC_LAB_DELIV | Complications of labour and delivery | XV Pregnancy, childbirth and the puerperium (O15_) | 48093 | 162777 |
| F5_MOOD | Mood [affective] disorders | V Mental and behavioural disorders (F5_) | 48085 | 329192 |
| I9_CORATHER | Coronary atherosclerosis | IX Diseases of the circulatory system (I9_) | 47550 | 313400 |
| N14_MALEGEN | Diseases of male genital organs | XIV Diseases of the genitourinary system (N14_) | 47110 | 119297 |
| G6_NERPLEX | Nerve, nerve root and plexus disorders | VI Diseases of the nervous system (G6_) | 46900 | 330377 |
| E4_DM2NASCOMP | Type 2 diabetes with other specified/multiple/unspecified complications | IV Endocrine, nutritional and metabolic diseases (E4_) | 46373 | 308280 |
| I9_AF | Atrial fibrillation and flutter | IX Diseases of the circulatory system (I9_) | 45766 | 191924 |
| H8_OTHEREAR | Other disorders of ear | VIII Diseases of the ear and mastoid process (H8_) | 45541 | 331736 |
| AB1_OTHER_BACTERIAL | Other bacterial diseases | I Certain infectious and parasitic diseases (AB1_) | 44934 | 332343 |
| M13_ARTHROSIS_KNEE | Gonarthrosis | XIII Diseases of the musculoskeletal system and connective tissue (M13_) | 44688 | 240862 |
| SLEEP | Sleep disorders (combined) | Neurological endpoints | 44299 | 329251 |
| I9_CHD | Major coronary heart disease event | IX Diseases of the circulatory system (I9_) | 43518 | 333759 |
| J10_CHRONTONSADEN | Chronic diseases of tonsils and adenoids | X Diseases of the respiratory system (J10_) | 43325 | 283342 |
| F5_DEPRESSIO | Depression | V Mental and behavioural disorders (F5_) | 43280 | 329192 |
| M13_SOFTTISSUEOTH | Other soft tissue disorders, not elsewhere classified | XIII Diseases of the musculoskeletal system and connective tissue (M13_) | 43190 | 275212 |
| J10_ASTHMA_EXMORE | Asthma (more control exclusions) | X Diseases of the respiratory system (J10_) | 42163 | 202399 |
| O15_MATERN_CARE | Maternal care related to the fetus and amniotic cavity and possible delivery problems | XV Pregnancy, childbirth and the puerperium (O15_) | 41941 | 168929 |
| M13_LOWBACKPAINORANDSCIATICA | Lower back pain or/and sciatica | XIII Diseases of the musculoskeletal system and connective tissue (M13_) | 41815 | 335462 |
| G6_SLEEPAPNO_INCLAVO | Sleep apnoea, including avohilmo | VI Diseases of the nervous system (G6_) | 41704 | 335573 |
| J10_ASTHMACOPDKELA | Asthma/COPD (KELA code 203) | X Diseases of the respiratory system (J10_) | 41495 | 311286 |
| E4_HYTHY_AI_STRICT | Hypothyroidism, strict autoimmune | IV Endocrine, nutritional and metabolic diseases (E4_) | 40926 | 274069 |
| AB1_INTESTINAL_INFECTIONS | Intestinal infectious diseases | I Certain infectious and parasitic diseases (AB1_) | 40881 | 336396 |

| | | | | |
|---|---|---|---|---|
| L12_DERMATITISECZEMA | Dermatitis and eczema | XII Diseases of the skin and subcutaneous tissue (L12_) | 40688 | 336589 |
| DEATH | Any death | Common endpoint | 40455 | 336822 |
| KRA_PSY_ANXIETY_EXMORE | Anxiety disorders (more control exclusions) | Psychiatric endpoints from Katri R√§ikk√∂nen | 40191 | 277526 |
| C_STROKE | STROKE | Comorbidities of Diabetes | 39818 | 271817 |
| MIGRAINE_TRIPTAN | Migraine, single triptan purchase ok & required. ICD-code if available is included | Neurological endpoints | 39387 | 337890 |
| G6_SLEEPAPNO | Sleep apnoea | VI Diseases of the nervous system (G6_) | 38998 | 336659 |
| T2D_WIDE | Type 2 diabetes, wide definition | Diabetes endpoints | 38657 | 310131 |
| N14_RENALTUB | Renal tubulo-interstitial diseases | XIV Diseases of the genitourinary system (N14_) | 38335 | 338942 |
| M13_SPONDYLOPATHY | Spondylopathies | XIII Diseases of the musculoskeletal system and connective tissue (M13_) | 38276 | 270964 |
| E4_LIPOPROT | Disorders of lipoprotein metabolism and other lipidaemias | IV Endocrine, nutritional and metabolic diseases (E4_) | 37742 | 324150 |
| M13_INTERVERTEB | Other intervertebral disc disorders | XIII Diseases of the musculoskeletal system and connective tissue (M13_) | 37636 | 270964 |
| K11_CHOLELITH | Cholelithiasis | XI Diseases of the digestive system (K11_) | 37041 | 330903 |
| N14_MESNRUIRREG | Excessive, frequent and irregular menstruation | XIV Diseases of the genitourinary system (N14_) | 36824 | 107564 |
| J10_ACUTEUPPERINFEC | Acute upper respiratory infections of multiple and unspecified sites | X Diseases of the respiratory system (J10_) | 36445 | 308166 |
| O15_ABORT_MEDICAL | Medical abortion | XV Pregnancy, childbirth and the puerperium (O15_) | 36232 | 149622 |
| K11_CONSTIPATION | Constipation | XI Diseases of the digestive system (K11_) | 36022 | 341255 |
| BLEEDING | Bleeding | Miscellaneous, not yet classified endpoints | 35917 | 341360 |
| M13_KNEEDERANGEMENTS | Internal derangement of knee | XIII Diseases of the musculoskeletal system and connective tissue (M13_) | 35511 | 240862 |
| M13_SHOULDER | Shoulder lesions | XIII Diseases of the musculoskeletal system and connective tissue (M13_) | 34594 | 275212 |
| I9_ANGINA | Angina pectoris | IX Diseases of the circulatory system (I9_) | 34456 | 313400 |
| J10_ASTHMA_MAIN_EXMORE | Asthma (only as main-diagnosis) (more control exclusions) | X Diseases of the respiratory system (J10_) | 34343 | 202399 |
| N14_URETHRAOTH | Other disorders of urethra and urinary system | XIV Diseases of the genitourinary system (N14_) | 32905 | 328001 |
| H7_CHOROIDRETINA | Disorders of choroid and retina | VII Diseases of the eye and adnexa (H7_) | 32708 | 344569 |
| H7_LIDLACRIMALORBIT | Disorders of eyelid, lacrimal system and orbit | VII Diseases of the eye and adnexa (H7_) | 32593 | 344684 |
| H8_HL_SEN_NAS | Sensorineural hearing loss | VIII Diseases of the ear and mastoid process (H8_) | 32487 | 331736 |
| K11_HERING | Inguinal hernia | XI Diseases of the digestive system (K11_) | 32335 | 322586 |
| M13_POLYARTHROPATHIES | Polyarthropathies | XIII Diseases of the musculoskeletal system and connective tissue (M13_) | 32199 | 240862 |
| CD2_BENIGN_LEIOMYOMA_UTERI | Leiomyoma of uterus | II Neoplasms from hospital discharges (CD2_) | 31661 | 179209 |

| | | | | |
|---|---|---|---|---|
| E4_PCOS_CONCORTIUM | Polycystic ovarian syndrome, consortium definition | IV Endocrine, nutritional and metabolic diseases (E4_) | 31548 | 179322 |
| O15_PREG_OTHER_MAT_DISORD | Other maternal disorders predominantly related to pregnancy | XV Pregnancy, childbirth and the puerperium (O15_) | 30971 | 179899 |
| K11_DIVERTIC | Diverticular disease of intestine | XI Diseases of the digestive system (K11_) | 30649 | 301931 |
| M13_LIMBPAIN | Pain in limb | XIII Diseases of the musculoskeletal system and connective tissue (M13_) | 30485 | 275212 |
| K11_PARODON_OPER | Parodontitis or operation codes | XI Diseases of the digestive system (K11_) | 30377 | 346900 |
| N14_PROSTHYPERPLA | Hyperplasia of prostate | XIV Diseases of the genitourinary system (N14_) | 30066 | 119297 |
| I9_VARICVE | Varicose veins | IX Diseases of the circulatory system (I9_) | 29539 | 324121 |
| AB1_GASTROENTERITIS_NOS | Diarrhoea and gastroenteritis of presumed infectious origin | I Certain infectious and parasitic diseases (AB1_) | 29401 | 336396 |
| M13_LOWBACKPAIN | Low back pain | XIII Diseases of the musculoskeletal system and connective tissue (M13_) | 29329 | 270964 |
| H7_CONJUNCTIVITIS | Conjunctivitis | VII Diseases of the eye and adnexa (H7_) | 28895 | 345122 |
| K11_APPENDACUT | Acute appendicitis | XI Diseases of the digestive system (K11_) | 28745 | 346283 |
| I9_OTHARR | Other arrhythmias | IX Diseases of the circulatory system (I9_) | 28079 | 286109 |
| K11_PARODON_OPER_DENTAL_OPER | Parodontitis or/and operation codes | XI Diseases of the digestive system (K11_) | 27833 | 349444 |
| JOINTPAIN | Pain in joint | XIII Diseases of the musculoskeletal system and connective tissue (M13_) | 27688 | 205355 |
| J10_TONSILLECTOMY | Tonsillectomy | X Diseases of the respiratory system (J10_) | 27449 | 349828 |
| D3_ANAEMIA | Anaemias | III Diseases of the blood and blood-forming organs and certain disorders involving the immune mechanism (D3_) | 27371 | 88536 |
| I9_HEARTFAIL | Heart failure,strict | IX Diseases of the circulatory system (I9_) | 27304 | 349973 |
| I9_HEARTFAIL_EXMORE | Heart failure,strict (more control exclusions) | IX Diseases of the circulatory system (I9_) | 27304 | 328105 |
| G6_OTHNEU | Other neurological diseases | VI Diseases of the nervous system (G6_) | 27026 | 350251 |
| I9_HEARTFAIL_ALLCAUSE | All-cause Heart Failure | IX Diseases of the circulatory system (I9_) | 26872 | 349361 |
| K11_CHOLECYSTECTOMY | Cholecystectomy | XI Diseases of the digestive system (K11_) | 26778 | 350499 |
| K11_REFLUX | Gastro-oesophageal reflux disease | Interstitial lung disease endpoints | 26184 | 320387 |
| K11_APPENDECTOMY | Appendectomy | XI Diseases of the digestive system (K11_) | 26013 | 351264 |
| E4_HYPERCHOL | Pure hypercholesterolaemia | IV Endocrine, nutritional and metabolic diseases (E4_) | 25928 | 324150 |
| I9_HEARTFAIL_AND_ANTIHYPERT | Heart failure and antihypertensive medication | Cardiometabolic endpoints | 25827 | 177731 |
| I9_STR | Stroke, excluding SAH | IX Diseases of the circulatory system (I9_) | 25398 | 339920 |

| | | | | |
|---|---|---|---|---|
| K11_CARIES_2 | Dental caries 2 | XI Diseases of the digestive system (K11_) | 25095 | 352182 |
| H8_MIDDLEMASTOID | Diseases of middle ear and mastoid | VIII Diseases of the ear and mastoid process (H8_) | 24706 | 352571 |
| F5_ALLANXIOUS | All anxiety disorders | V Mental and behavioural disorders (F5_) | 24662 | 337577 |
| L12_OTHERSKINSUBCUTIS | Other disorders of skin and subcutaneous tissue | XII Diseases of the skin and subcutaneous tissue (L12_) | 24189 | 353088 |
| I9_MI_STRICT | Myocardial infarction, strict | IX Diseases of the circulatory system (I9_) | 24185 | 313400 |
| M13_MENISCUSDERANGEMENTS | Meniscus derangement | XIII Diseases of the musculoskeletal system and connective tissue (M13_) | 24158 | 240862 |
| M13_ROTATORCUFF | Rotator cuff syndrome | XIII Diseases of the musculoskeletal system and connective tissue (M13_) | 24061 | 275212 |
| N14_PYELONEPHR | Acute tubulo-interstitial nephritis | XIV Diseases of the genitourinary system (N14_) | 23912 | 338942 |
| H7_RETINALDISOTH | Other retinal disorders | VII Diseases of the eye and adnexa (H7_) | 23610 | 344569 |
| KRA_PSY_SUBSTANCE_EXMORE | Substance abuse (more control exclusions) | Psychiatric endpoints from Katri R√§ikk√∂nen | 23194 | 277526 |
| I9_REVASC | Coronary revascularization (ANGIO or CABG) | IX Diseases of the circulatory system (I9_) | 23139 | 313400 |
| N14_FEMALEGENINF | Inflammatory diseases of female pelvic organs | XIV Diseases of the genitourinary system (N14_) | 22575 | 188295 |
| G6_CARPTU | Carpal tunnel syndrome | VI Diseases of the nervous system (G6_) | 22426 | 330377 |
| M13_ARTHTROSIS_COX | Coxarthrosis, | XIII Diseases of the musculoskeletal system and connective tissue (M13_) | 22254 | 240862 |
| FG_DOAAC | Diseases of arteries, arterioles and capillaries (FINNGEN) | IX Diseases of the circulatory system (I9_) | 22224 | 191924 |
| APPENDACUT_NOCOMPLIC | Acute appendicitis, no complications | XI Diseases of the digestive system (K11_) | 21998 | 346283 |
| AUD_SWEDISH | Alcohol use disorder, Swedish definition | Alcohol related diseases | 21996 | 355281 |
| L12_DERMATITISNAS | Other and unspecified dermatitis | XII Diseases of the skin and subcutaneous tissue (L12_) | 21951 | 336589 |
| ASTHMA_INFECTIONS | Asthma-related infections | Asthma and related endpoints | 21927 | 335114 |
| I9_AF_REIMB | Atrial fibrillation and flutter with reimbursement | IX Diseases of the circulatory system (I9_) | 21700 | 191924 |
| N14_MENORRHAGIA | Menorrhagia | XIV Diseases of the genitourinary system (N14_) | 21496 | 107564 |
| ASTHMA_NAS | Asthma, unspecified (mode) | Asthma and related endpoints | 21392 | 210122 |
| E4_OBESITY | Obesity | IV Endocrine, nutritional and metabolic diseases (E4_) | 21375 | 355786 |
| L12_INFECT_SKIN | Infections of the skin and subcutaneous tissue | XII Diseases of the skin and subcutaneous tissue (L12_) | 21308 | 355969 |
| H7_ALLERGICCONJUNCTIVITIS | Allergic conjunctivitis | VII Diseases of the eye and adnexa (H7_) | 20958 | 356319 |
| C3_SKIN_EXALLC | Malignant neoplasm of skin (controls excluding all cancers) | II Neoplasms, from cancer register (ICD-O-3) | 20951 | 287137 |
| M13_ARTHROSIS_OTH | Other arthrosis | XIII Diseases of the musculoskeletal system and connective tissue (M13_) | 20949 | 240862 |
| I9_VHD | Valvular heart disease excluding rheumatic fever | IX Diseases of the circulatory system (I9_) | 20929 | 286109 |
| J10_LOWERINF | Other acute lower respiratory infections | X Diseases of the respiratory system (J10_) | 20873 | 356404 |
| I9_NONRHEVALV | Non-rheumatic valve diseases | IX Diseases of the circulatory system (I9_) | 20772 | 286109 |

| | | | | |
|---|---|---|---|---|
| N14_OVARYCYST | Ovarian cyst | XIV Diseases of the genitourinary system (N14_) | 20750 | 107564 |
| K11_REIMB_202 | KELA_REIMBURSEMENT_202 | Gastrointestinal endpoints | 20730 | 356547 |
| J10_SINUSITIS | Acute sinusitis | X Diseases of the respiratory system (J10_) | 20520 | 308166 |
| M13_SYNOTEND | Disorders of synovium and tendon | XIII Diseases of the musculoskeletal system and connective tissue (M13_) | 20492 | 275212 |
| AB1_ERYSIPELAS | Erysipelas | I Certain infectious and parasitic diseases (AB1_) | 20470 | 332343 |
| RX_CROHN_2NDLINE | Second line medication for Crohn's disease | Drug purchase endpoints | 20353 | 356924 |
| CD2_BENIGN_COLORECANI | Benign neoplasm of colon, rectum, anus and anal canal | II Neoplasms from hospital discharges (CD2_) | 20336 | 356941 |
| M13_ARTHTROSIS_COX_PRIM_ICD10 | Coxarthrosis, primary | XIII Diseases of the musculoskeletal system and connective tissue (M13_) | 20056 | 357221 |
| ST19_FRACT_LOWER_LEG_INCLU_ANKLE | Fracture of lower leg, including ankle | XIX Injury, poisoning and certain other consequences of external causes (ST19_) | 19994 | 323307 |
| M13_SPONDYLOSIS | Spondylosis | XIII Diseases of the musculoskeletal system and connective tissue (M13_) | 19892 | 270964 |
| ST19_FRACT_FOREA | Fracture of forearm | XIX Injury, poisoning and certain other consequences of external causes (ST19_) | 19577 | 351196 |
| J10_VOCALLARYNX | Diseases of vocal cords and larynx+other diseases of upper respiratory tract, no elsewhere classified | X Diseases of the respiratory system (J10_) | 19530 | 283342 |
| C3_BASAL_CELL_CARCINOMA_INCLAVO_EXALLC | Basal cell carcinoma, including avohilmo (controls excluding all cancers) | II Neoplasms, from cancer register (ICD-O-3) | 19522 | 286768 |
| H7_SCLERACORNEA | Disorders of sclera, cornea, iris and ciliary body | VII Diseases of the eye and adnexa (H7_) | 19463 | 357814 |
| I9_VTE | Venous thromboembolism | IX Diseases of the circulatory system (I9_) | 19372 | 357905 |
| I9_K_CARDIAC | Death due to cardiac causes | IX Diseases of the circulatory system (I9_) | 19295 | 357982 |
| M13_OSTEOCHONDRO | Osteopathies and chondropathies | XIII Diseases of the musculoskeletal system and connective tissue (M13_) | 19263 | 358014 |
| M13_ARTHROSIS_KNEE_PRIM_KNEESURG | Gonarthrosis, primary, with knee surgery | XIII Diseases of the musculoskeletal system and connective tissue (M13_) | 19126 | 337196 |
| M13_SPINSTENOSIS | Spinal stenosis | XIII Diseases of the musculoskeletal system and connective tissue (M13_) | 18989 | 270964 |
| C3_BASAL_CELL_CARCINOMA_EXALLC | Basal cell carcinoma (controls excluding all cancers) | II Neoplasms, from cancer register (ICD-O-3) | 18982 | 287137 |
| H7_GLAUCOMA | Glaucoma | VII Diseases of the eye and adnexa (H7_) | 18902 | 358375 |
| AB1_BACTINF_NOS | Bacterial infection, other or unspecified | I Certain infectious and parasitic diseases (AB1_) | 18576 | 332343 |
| M13_SCIATICA | Sciatica+with lumbago | XIII Diseases of the musculoskeletal system and connective tissue (M13_) | 18569 | 270964 |
| G6_MIGRAINE | Migraine | VI Diseases of the nervous system (G6_) | 18477 | 287837 |
| I9_TIA | Transient ischemic attack | IX Diseases of the circulatory system (I9_) | 18398 | 342294 |

| | | | | |
|---|---|---|---|---|
| RX_GLUCOSAMINE | Glucosamine medication | Drug purchase endpoints | 18341 | 358936 |
| J10_COPD | COPD | X Diseases of the respiratory system (J10_) | 18266 | 311286 |
| N14_CYSTITIS | Cystitis | XIV Diseases of the genitourinary system (N14_) | 18080 | 328001 |
| C3_OTHER_SKIN_EXALLC | Other malignant neoplasms of skin (=non-melanoma skin cancer) (controls excluding all cancers) | II Neoplasms, from cancer register (ICD-O-3) | 17958 | 287137 |
| F5_DEMENTIA_INCLAVO | Dementia, including avohilmo | V Mental and behavioural disorders (F5_) | 17901 | 359376 |
| H7_CATARACTOTHER | Other cataract | VII Diseases of the eye and adnexa (H7_) | 17699 | 312864 |
| K11_ENERCOLNONINF | Noninfective enteritis and colitis | XI Diseases of the digestive system (K11_) | 17350 | 359927 |
| N14_MENSIRREG | Irregular menses | XIV Diseases of the genitourinary system (N14_) | 17228 | 179322 |
| F5_DEPRESSION_RECURRENT | Recurrent or chronic depression | V Mental and behavioural disorders (F5_) | 17227 | 228647 |
| N14_FEMGENPROL | Female genital prolapse | XIV Diseases of the genitourinary system (N14_) | 17150 | 107564 |
| PRIM_KNEEARTHROSIS | Primary gonarthrosis, bilateral | Rheuma endpoints | 17009 | 332589 |
| O15_ABORT_SPONTAN | Spontaneous abortion | XV Pregnancy, childbirth and the puerperium (O15_) | 16906 | 149622 |
| N14_BREAST | Disorders of breast | XIV Diseases of the genitourinary system (N14_) | 16897 | 193973 |
| F5_ANXIETY | Other anxiety disorders | V Mental and behavioural disorders (F5_) | 16887 | 337577 |
| I9_HEARTFAIL_AND_OVERWEIGHT | Heart failure and bmi 25plus | Cardiometabolic endpoints | 16707 | 154410 |
| F5_DEMENTIA | Dementia | V Mental and behavioural disorders (F5_) | 16499 | 356660 |
| CD2_BENIGN_COLON | Benign neoplasm: Colon | II Neoplasms from hospital discharges (CD2_) | 16445 | 360832 |
| J10_CHRONSINUSITIS | Chronic sinusitis | X Diseases of the respiratory system (J10_) | 16395 | 283342 |
| DM_SEVERAL_COMPLICATIONS | Diabetes, several complications | Diabetes endpoints | 16320 | 271817 |
| K11_OTHFUNC | Other functional intestinal disorders | XI Diseases of the digestive system (K11_) | 16311 | 301931 |
| J10_PNEUMOBACT | Bacterial pneumoniae | X Diseases of the respiratory system (J10_) | 16244 | 314673 |
| K11_OTHDIG | Other diseases of the digestive system | XI Diseases of the digestive system (K11_) | 16222 | 361055 |
| L12_DISORDSKINAPPENDIX | Disorders of skin appendages | XII Diseases of the skin and subcutaneous tissue (L12_) | 16137 | 361140 |
| H7_OCUMUSCLE | Disorders of ocular muscles, binocular movement, accommodation and refraction | VII Diseases of the eye and adnexa (H7_) | 16040 | 361237 |
| ASTHMA_ACUTE_RESPIRATORY_INFECTIONS | Asthma-related acute respiratory infections | Asthma and related endpoints | 16018 | 335114 |
| G6_HEADACHE | Other headache syndromes | VI Diseases of the nervous system (G6_) | 15851 | 287837 |
| AUD | Alcohol use disorder, ICD-based | V Mental and behavioural disorders (F5_) | 15715 | 361562 |

| | | | | |
|---|---|---|---|---|
| H7_VISUDISTURB | Visual disturbances | VII Diseases of the eye and adnexa (H7_) | 15707 | 360091 |
| C3_BREAST_EXALLC | Malignant neoplasm of breast (controls excluding all cancers) | II Neoplasms, from cancer register (ICD-O-3) | 15680 | 167189 |
| F5_KELAMENT | Kela-code for severe mental illness | V Mental and behavioural disorders (F5_) | 15451 | 360114 |
| J10_PLEURA | Other diseases of pleura | X Diseases of the respiratory system (J10_) | 15441 | 361836 |
| J10_BRONCHITIS | Acute bronchitis | X Diseases of the respiratory system (J10_) | 15393 | 356404 |
| ABDOM_HERNIA | Hernia of abdominal wall | XI Diseases of the digestive system (K11_) | 15338 | 361939 |
| E4_ENDOGLAND | Disorders of other endocrine glands | IV Endocrine, nutritional and metabolic diseases (E4_) | 15289 | 361988 |
| H7_VITRBODYGLOBE | Disorders of vitreous body | VII Diseases of the eye and adnexa (H7_) | 15177 | 361552 |
| H7_MACULADEGEN | Degeneration of macula and posterior pole | VII Diseases of the eye and adnexa (H7_) | 15137 | 344569 |
| N14_ENDOMETRIOSIS | Endometriosis | XIV Diseases of the genitourinary system (N14_) | 15088 | 107564 |
| E4_OBESITYCAL | Obesity due to excess calories | IV Endocrine, nutritional and metabolic diseases (E4_) | 15045 | 355902 |
| I9_ATHSCLE | Atherosclerosis, excluding cerebral, coronary and PAD | IX Diseases of the circulatory system (I9_) | 15002 | 349539 |
| M13_ARTHTROSIS_COX_PRIM_HIPSURG | Coxarthrosis, primary, with hip surgery | XIII Diseases of the musculoskeletal system and connective tissue (M13_) | 14990 | 357221 |
| H8_VERTIGO | Disorders of vestibular function (Vertigo) | VIII Diseases of the ear and mastoid process (H8_) | 14918 | 359094 |
| O15_HYPTENSPREG | Pregnancy hypertension | XV Pregnancy, childbirth and the puerperium (O15_) | 14727 | 196143 |
| M13_HALLUXVALGUS | Hallux valgus (acquired) | XIII Diseases of the musculoskeletal system and connective tissue (M13_) | 14726 | 240862 |
| I9_ANGIO | Coronary angioplasty | IX Diseases of the circulatory system (I9_) | 14723 | 313400 |
| H8_MED_SUPP | Suppurative and unspecified otitis media | VIII Diseases of the ear and mastoid process (H8_) | 14406 | 352571 |
| KRA_PSY_DEMENTIA_EXMORE | Any dementia (more control exclusions) | Psychiatric endpoints from Katri R√§ikk√∂nen | 14367 | 277526 |
| H7_EYELIDDIS | Other disorders of eyelid | VII Diseases of the eye and adnexa (H7_) | 14347 | 344684 |
| N14_POLYPFEMGEN | Polyp of the female genital tract | XIV Diseases of the genitourinary system (N14_) | 14324 | 107564 |
| O15_FALSE_LAB | False labour | XV Pregnancy, childbirth and the puerperium (O15_) | 14282 | 168929 |
| AB1_VIRAL_SKIN_MUCOUS_MEMBRANE | Viral infections characterized by skin and mucous membrane lesions | I Certain infectious and parasitic diseases (AB1_) | 14280 | 362997 |
| N14_RENFAIL | Renal failure | XIV Diseases of the genitourinary system (N14_) | 14100 | 363177 |
| M13_DORSALGIANAS | Other/unspecified dorsalgia | XIII Diseases of the musculoskeletal system and connective tissue (M13_) | 14075 | 270964 |
| F5_STRESSOTH | Other reaction to severe stress, and adjustment disorders | V Mental and behavioural disorders (F5_) | 13913 | 337577 |
| D3_ANAEMIA_IRONDEF | Iron deficiency anaemia | III Diseases of the blood and blood-forming organs and certain disorders involving the immune mechanism (D3_) | 13689 | 360528 |
| D3_ANAEMIANAS | Other and unspecified anaemias | III Diseases of the blood and blood-forming organs and certain disorders involving the immune mechanism (D3_) | 13600 | 362319 |
| L12_ATOPIC | Atopic dermatitis | XII Diseases of the skin and subcutaneous tissue (L12_) | 13473 | 336589 |

| | | | | |
|---|---|---|---|---|
| M13_CERVICDISC | Cervical disc disorders | XIII Diseases of the musculoskeletal system and connective tissue (M13_) | 13394 | 270964 |
| G6_AD_WIDE | Alzheimer‚Äôs disease, wide definition | VI Diseases of the nervous system (G6_) | 13393 | 363884 |
| H7_LACRIMALSYSTEM | Disorders of lacrimal system | VII Diseases of the eye and adnexa (H7_) | 13326 | 344684 |
| I9_UAP | Unstable angina pectoris | IX Diseases of the circulatory system (I9_) | 13304 | 333375 |
| TEMPOROMANDIB_INCLAVO | Temporomandibular joint disorders, including avohilmo | XI Diseases of the digestive system (K11_) | 13282 | 363995 |
| C3_PROSTATE_EXALLC | Malignant neoplasm of prostate (controls excluding all cancers) | II Neoplasms, from cancer register (ICD-O-3) | 13216 | 119948 |
| L12_PAPULOSQUAMOUS | Papulosquamous disorders | XII Diseases of the skin and subcutaneous tissue (L12_) | 13206 | 364071 |
| J10_TONSILLITIS | Other and unspecified tonsillitis | X Diseases of the respiratory system (J10_) | 13190 | 308166 |
| N14_FEMALEINFERT | Female infertility | XIV Diseases of the genitourinary system (N14_) | 13142 | 107564 |
| NEURODEGOTH | Other degenerative diseases of the nervous system | Comorbidities of Gastrointestinal endpoints | 13086 | 212179 |
| KRA_PSY_SCHIZODEL_EXMORE | Schizophrenia or delusion (more control exclusions) | Psychiatric endpoints from Katri R√§ikk√∂nen | 13061 | 277526 |
| GEST_DIABETES | Gestational diabetes (for exclusion) | Diabetes endpoints | 13039 | 197831 |
| I9_HEARTFAIL_AND_CHD | Heart failure and coronary heart disease | Cardiometabolic endpoints | 12872 | 333759 |
| I9_VEINSOTH | Other disorders of veins | IX Diseases of the circulatory system (I9_) | 12630 | 324121 |
| O15_DELIV_CAESAR | Single delivery by caesarean section | XV Pregnancy, childbirth and the puerperium (O15_) | 12596 | 92306 |
| M13_RHEUMA | Rheumatoid arthritis | XIII Diseases of the musculoskeletal system and connective tissue (M13_) | 12555 | 240862 |
| CD2_BENIGN_SKIN | Other benign neoplasms of skin | II Neoplasms from hospital discharges (CD2_) | 12341 | 364936 |
| E4_HYPERLIPNAS | Hyperlipidaemia, other/unspecified | IV Endocrine, nutritional and metabolic diseases (E4_) | 12304 | 324150 |
| AB1_OTHER_SEPSIS | Other septicaemia | I Certain infectious and parasitic diseases (AB1_) | 12301 | 332343 |
| O15_OTHER_MATERN_DIS_ELSEWHERE | Other maternal diseases classifiable elsewhere but complicating pregnancy, childbirth and the puerperium | XV Pregnancy, childbirth and the puerperium (O15_) | 12297 | 198180 |
| K11_PULP_PERIAPICAL | Diseases of pulp and periapical tissues | XI Diseases of the digestive system (K11_) | 12078 | 259234 |
| ASTHMA_PNEUMONIA | Asthma-related pneumonia | Asthma and related endpoints | 12044 | 335114 |
| HYPOTHY_REIMB | Hypothyroidism, drug reimbursement | IV Endocrine, nutritional and metabolic diseases (E4_) | 12040 | 88531 |
| H7_VITROTH | Other and unspecified disorders of vitreous body | VII Diseases of the eye and adnexa (H7_) | 12004 | 361552 |
| K11_DIAHER | Diaphragmatic hernia | XI Diseases of the digestive system (K11_) | 11757 | 322586 |
| ST19_FRACT_SHOUL_UPPER_ARM | Fracture of shoulder and upper arm | XIX Injury, poisoning and certain other consequences of external causes (ST19_) | 11750 | 345173 |
| M13_SYSTCONNECT | Systemic connective tissue disorders | XIII Diseases of the musculoskeletal system and connective tissue (M13_) | 11744 | 365533 |

| | | | | |
|---|---|---|---|---|
| G6_EPLEPSY | Epilepsy | VI Diseases of the nervous system (G6_) | 11740 | 287837 |
| O15_CONCEPT_ABNORM | Other abnormal products of conception | XV Pregnancy, childbirth and the puerperium (O15_) | 11716 | 149622 |
| J10_SEPTDEV | Deviated nasal septum | X Diseases of the respiratory system (J10_) | 11680 | 283342 |
| Z21_PROCREATIVE_MANAG | Procreative management | XXI Factors influencing health status and contact with health services (Z21_) | 11666 | 300151 |
| R18_RETEN_URINE | Retention of urine | XVIII Symptoms, signs and abnormal clinical and laboratory findings, not elsewhere classified (R18_) | 11657 | 343006 |
| H7_KERATITIS | Keratitis | VII Diseases of the eye and adnexa (H7_) | 11611 | 357814 |
| N14_POSTMENBLEED | Postmenopausal bleeding | XIV Diseases of the genitourinary system (N14_) | 11582 | 107564 |
| I9_SEQULAE | Sequelae of cerebrovascular disease | IX Diseases of the circulatory system (I9_) | 11408 | 342673 |
| ST19_FRACT_WRIST_HAND_LEVEL | Fracture at wrist and hand level | XIX Injury, poisoning and certain other consequences of external causes (ST19_) | 11394 | 336418 |
| N14_FIOTHNAS | Female infertility, cervigal, vaginal, other or unspecified origin | XIV Diseases of the genitourinary system (N14_) | 11348 | 107564 |
| H7_RETINALDETACH | Retinal detachments and breaks | VII Diseases of the eye and adnexa (H7_) | 11230 | 344569 |
| L12_ACTINKERA | Actinic keratosis | XII Diseases of the skin and subcutaneous tissue (L12_) | 11081 | 364831 |
| RHEU_ARTHRITIS_OTH | Other arthritis (FG) | Rheuma endpoints | 11033 | 285035 |
| E4_OBESITYNAS | Obesity, other/unspecified | IV Endocrine, nutritional and metabolic diseases (E4_) | 11025 | 355902 |
| ALLERG_RHINITIS | Allergic rhinitis | Comorbidities of Asthma | 11009 | 359149 |
| I9_CABG | Coronary artery bypass grafting | IX Diseases of the circulatory system (I9_) | 10938 | 313400 |
| AB1_SEXUAL_TRANSMISSION | Infections with a predominantly sexual mode of transmission | I Certain infectious and parasitic diseases (AB1_) | 10937 | 366340 |
| J10_CHRONRHINITIS | Chronic rhinitis, nasopharyngitis and pharyngitis | X Diseases of the respiratory system (J10_) | 10868 | 283342 |
| K11_LIVER | Diseases of liver | XI Diseases of the digestive system (K11_) | 10827 | 366450 |
| KRA_PSY_ANYMENTAL_SUICID_PREG_NERV_EXMORE | Any mental disorder, or suicide (or attempt), or psychic disorders complicating pregnancy, partum or puerperium or nerve system disorders (more control exclusions) | Psychiatric endpoints from Katri R√§ikk√∂nen | 10811 | 276857 |
| H7_GLAUCSUSP | Glaucoma suspect | VII Diseases of the eye and adnexa (H7_) | 10758 | 358375 |
| K11_OTHDISDIG | Other diseases of the digestive system | XI Diseases of the digestive system (K11_) | 10589 | 361055 |
| M13_ENTESOPATHYOTH | Other enthesopathies | XIII Diseases of the musculoskeletal system and connective tissue (M13_) | 10581 | 275212 |
| DM_RETINOPATHY_EXMORE | Diabetic retinopathy (more control exclusions) | Diabetes endpoints | 10413 | 308633 |
| COPD_LATER | Later onset COPD | COPD and related endpoints | 10404 | 161813 |
| F5_BEHAVE | Behavioural syndromes associated with physiological | V Mental and behavioural disorders (F5_) | 10401 | 366876 |

| | | | | |
|---|---|---|---|---|
| | disturbances and physical factors | | | |
| N14_ABNORMALBLEED | Other abnormal uterine and caginal bleeding | XIV Diseases of the genitourinary system (N14_) | 10319 | 107564 |
| ASTHMA_OBESITY | Obesity related asthma | Comorbidities of Asthma | 10306 | 335114 |
| DENTAL_TMD | TMD related pain | Dental endpoints | 10303 | 366974 |
| O15_PREEC_OR_FETGRO | Pre-eclampsia or poor fetal growth | XV Pregnancy, childbirth and the puerperium (O15_) | 10297 | 200573 |
| H8_EXTERNAL | Diseases of external ear | VIII Diseases of the ear and mastoid process (H8_) | 10272 | 367005 |
| D3_ANAEMIA_IRONDEF_NAS | Other and unspecified iron deficiency | III Diseases of the blood and blood-forming organs and certain disorders involving the immune mechanism (D3_) | 10208 | 360528 |
| L12_URTICARIA | Urticaria | XII Diseases of the skin and subcutaneous tissue (L12_) | 10175 | 364583 |
| KRA_PSY_PERSON_EXMORE | Personality disorders (more control exclusions) | Psychiatric endpoints from Katri R√§ikk√∂nen | 10012 | 277522 |
| I9_CONDUCTIO | Conduction disorders | IX Diseases of the circulatory system (I9_) | 9949 | 286109 |
| K11_OTHANRECT | Other diseases of anus and rectum | XI Diseases of the digestive system (K11_) | 9932 | 301931 |
| I9_NONISCHCARDMYOP | Non-ischemic cardiomyopathy | IX Diseases of the circulatory system (I9_) | 9926 | 303607 |
| F5_ALCOHOL_DEPENDENCE | Alcohol dependence | V Mental and behavioural disorders (F5_) | 9876 | 353355 |
| K11_DENTOFACIAL_ANOMALIES | Dentofacial anomalies [including malocclusion] | XI Diseases of the digestive system (K11_) | 9866 | 259234 |
| H8_SUP_ACUTE | Acute suppurative otitis media | VIII Diseases of the ear and mastoid process (H8_) | 9850 | 352571 |
| G6_XTRAPYR | Extrapyramidal and movement disorders | VI Diseases of the nervous system (G6_) | 9833 | 367444 |
| AB1_OTHER_VIRAL | Other viral diseases | I Certain infectious and parasitic diseases (AB1_) | 9805 | 367472 |
| O15_MATERN_CARE_OTHER | Maternal care for other known or suspected fetal problems | XV Pregnancy, childbirth and the puerperium (O15_) | 9783 | 168929 |
| N14_CALCUKIDUR | Calculus of kidney and ureter | XIV Diseases of the genitourinary system (N14_) | 9713 | 366693 |
| C3_BREAST_ERPLUS_EXALLC | Malignant neoplasm of breast, HER-positive (controls excluding all cancers) | II Neoplasms, from cancer register (ICD-O-3) | 9698 | 167017 |
| E4_FLUIDELECTRO | Other disorders of fluid, electrolyte and acid-base balance | IV Endocrine, nutritional and metabolic diseases (E4_) | 9694 | 324150 |
| M13_DEFORMDORSO | Deforming dorsopathies | XIII Diseases of the musculoskeletal system and connective tissue (M13_) | 9672 | 270964 |
| ALLERG_ASTHMA | Allergic asthma (mode) | Asthma and related endpoints | 9631 | 210122 |
| I9_PAROXTAC | Paroxysmal tachycardia | IX Diseases of the circulatory system (I9_) | 9604 | 191924 |
| K11_CHRONGASTR | Chronic gastritis | XI Diseases of the digestive system (K11_) | 9570 | 320387 |
| DM_RETINA_PROLIF | Proliferative diabetic retinopathy | Diabetes endpoints | 9511 | 362581 |
| E4_NONTOXIC_THYROID | Nontoxic goitre/thyroid nodule | IV Endocrine, nutritional and metabolic diseases (E4_) | 9485 | 367792 |

| | | | | |
|---|---|---|---|---|
| O15_HAEMORRH_EARLY_PREG | Haemorrhage in early pregnancy | XV Pregnancy, childbirth and the puerperium (O15_) | 9433 | 179899 |
| CD2_BENIGN_LIPOMATOUS | Benign lipomatous neoplasm | II Neoplasms from hospital discharges (CD2_) | 9431 | 367846 |
| K11_IBS | Irritable bowel syndrome | XI Diseases of the digestive system (K11_) | 9323 | 301931 |
| G6_ALZHEIMER | Alzheimer disease | VI Diseases of the nervous system (G6_) | 9301 | 367976 |
| L12_PSORIASIS | Psoriasis | XII Diseases of the skin and subcutaneous tissue (L12_) | 9267 | 364071 |
| I9_PULMEMB | Pulmonary embolism | IX Diseases of the circulatory system (I9_) | 9243 | 367108 |
| RHEUMA_SEROPOS_WIDE | Seropositive rheumatoid arthritis, wide | Rheuma endpoints | 9243 | 368029 |
| N14_FEMALE_GENITAL_DYSPLASIA_ALL | All dysplastic lesions of the cervix uteri, vagina or vulva | XIV Diseases of the genitourinary system (N14_) | 9227 | 201643 |
| K11_GASTRODUOULC | Gastroduodenal ulcer | XI Diseases of the digestive system (K11_) | 9216 | 320387 |
| I9_CAVS_OPERATED | Calcific aortic valvular stenosis, operated | IX Diseases of the circulatory system (I9_) | 9153 | 368124 |
| O15_MATERN_CARE_PELVIC_ABNORM | Maternal care for known or suspected abnormality of pelvic organs | XV Pregnancy, childbirth and the puerperium (O15_) | 9149 | 168929 |
| RHEUMA_SEROPOS_OTH | Other/unspecified seropositive rheumatoid arthritis | Rheuma endpoints | 9139 | 368029 |
| I9_PHLETHROMBDVTLOW | DVT of lower extremities | IX Diseases of the circulatory system (I9_) | 9109 | 324121 |
| ST19_FRACT_RIBS_STERNUM_THORACIC_SPINE | Fracture of rib(s), sternum and thoracic spine | XIX Injury, poisoning and certain other consequences of external causes (ST19_) | 9094 | 362260 |
| N14_CHRONKIDNEYDIS | Chronic kidney disease | XIV Diseases of the genitourinary system (N14_) | 9073 | 363177 |
| DENTAL_TMD_FIBRO | TMD muscular pain linked with fibromyalgia | Dental endpoints | 8995 | 368282 |
| VWXY20_SUICI_OTHER_INTENTI_SELF_H | Suicide or other Intentional self-harm | Psychiatric endpoints from Katri R√§ikk√∂nen | 8978 | 368299 |
| T1D_WIDE | Type 1 diabetes, wide definition | Diabetes endpoints | 8967 | 308373 |
| F5_ALCOHOLAC | Acute alcohol intoxication | V Mental and behavioural disorders (F5_) | 8957 | 353355 |
| H7_AMD | Age-related macular degeneration (whether dry or wet) | VII Diseases of the eye and adnexa (H7_) | 8913 | 348936 |
| H7_CONJUNCTIVITISACUNONATOPIC | Conjunctivitis (acute, non atopic) | VII Diseases of the eye and adnexa (H7_) | 8907 | 345122 |
| CD2_BENIGN_MELANOCYTIC | Melanocytic naevi | II Neoplasms from hospital discharges (CD2_) | 8900 | 368377 |
| K11_FISTULA | Colonic or urogenital fistula | XI Diseases of the digestive system (K11_) | 8879 | 368398 |
| K11_FUNCDYSP | Functional dyspepsia | XI Diseases of the digestive system (K11_) | 8875 | 320387 |

| | | | | |
|---|---|---|---|---|
| H8_OTHHEARINGLOSS | Other hearing loss | VIII Diseases of the ear and mastoid process (H8_) | 8833 | 331736 |
| ST19_FRACT_FEMUR | Fracture of femur | XIX Injury, poisoning and certain other consequences of external causes (ST19_) | 8766 | 360928 |
| N14_OTHNONINFUTER | Other noninflammatory disorders of uterus, except cervix | XIV Diseases of the genitourinary system (N14_) | 8752 | 107564 |
| OTHER_SYSTCON_FG | Other systemic involvement of connective tissue (FG) | Rheuma endpoints | 8695 | 285035 |
| M13_IMPINGEMENT | Impingement syndrome of shoulder | XIII Diseases of the musculoskeletal system and connective tissue (M13_) | 8676 | 275212 |
| K11_ILEUS | Paralytic ileus and intestinal obstruction | XI Diseases of the digestive system (K11_) | 8660 | 301931 |
| O15_COMPLIC_PUERP | Complications predominantly related to the puerperium | XV Pregnancy, childbirth and the puerperium (O15_) | 8603 | 202267 |
| PRIM_COXARTHROSIS | Primary coxarthrosis, bilateral | Rheuma endpoints | 8597 | 355023 |
| K11_HAEMORR | Haemmorrhoids and perianal venous thrombosis | XI Diseases of the digestive system (K11_) | 8583 | 301931 |
| J10_INFLUENZA | All influenza | X Diseases of the respiratory system (J10_) | 8555 | 314673 |
| H7_BLEPHAROCHALASIS | Blepharochalasis | VII Diseases of the eye and adnexa (H7_) | 8547 | 344684 |
| K11_UMBHER | Umbilical hernia | XI Diseases of the digestive system (K11_) | 8510 | 322586 |
| O15_PRETERM | Preterm labour and delivery | XV Pregnancy, childbirth and the puerperium (O15_) | 8507 | 162777 |
| O15_GESTAT_HYPERT | Gestational [pregnancy-induced] hypertension | XV Pregnancy, childbirth and the puerperium (O15_) | 8502 | 194266 |
| M13_GOUT | Gout | XIII Diseases of the musculoskeletal system and connective tissue (M13_) | 8489 | 240862 |
| I9_HYPTENSHD | Hypertensive Heart Disease | IX Diseases of the circulatory system (I9_) | 8387 | 265626 |
| M13_EARLY_LUMBAR_PROLAPSE | Early lumbar disc prolapse, operated | XIII Diseases of the musculoskeletal system and connective tissue (M13_) | 8351 | 368926 |
| O15_LABOUR_FETAL_STRESS | Labour and delivery complicated by fetal stress [distress] | XV Pregnancy, childbirth and the puerperium (O15_) | 8324 | 162777 |
| H8_NONSUPPNAS | Nonsuppurative otitis media | VIII Diseases of the ear and mastoid process (H8_) | 8315 | 352571 |
| H8_BPV | Benign paroxysmal vertigo | VIII Diseases of the ear and mastoid process (H8_) | 8280 | 359094 |
| O15_POSTPART_HEAMORRH | Postpartum haemorrhage | XV Pregnancy, childbirth and the puerperium (O15_) | 8249 | 162777 |
| O15_POSTPART_HEAMORRH_ALLW | Postpartum haemorrhage | XV Pregnancy, childbirth and the puerperium (O15_) | 8249 | 202621 |
| O15_POSTPART_HEAMORRH_LB | Postpartum haemorrhage | XV Pregnancy, childbirth and the puerperium (O15_) | 8249 | 146016 |
| M13_SOFTOVERUSE | Soft tissue disorders related to use, overuse and pressure | XIII Diseases of the musculoskeletal system and connective tissue (M13_) | 8236 | 275212 |
| AB1_MYCOSES | Mycoses | I Certain infectious and parasitic diseases (AB1_) | 8179 | 369098 |
| THYROTOXICOSIS | Thyrotoxicosis | Comorbidities of Gastrointestinal endpoints | 8173 | 367407 |

| | | | | |
|---|---|---|---|---|
| L12_ABSCESS_CUT | Cutaneous abscess, furuncle and carbuncle | XII Diseases of the skin and subcutaneous tissue (L12_) | 8157 | 355969 |
| K11_EMBIMPACT_TEETH_INCLAVO | Embedded and impacted teeth , including avohilmo | XI Diseases of the digestive system (K11_) | 8139 | 369138 |
| N14_CERVICAL_DYSPLASIA_ALL | All dysplastic lesions of the cervix uteri | XIV Diseases of the genitourinary system (N14_) | 8025 | 202845 |
| K11_OTHENTERCOL | Other noninfective gastroenteritis and colitis | XI Diseases of the digestive system (K11_) | 7988 | 359927 |
| M13_FIBROBLASTIC | Fibroblastic disorders | XIII Diseases of the musculoskeletal system and connective tissue (M13_) | 7952 | 275212 |
| G6_MIGRAINE_WITH_AURA | Migraine with aura | VI Diseases of the nervous system (G6_) | 7917 | 287837 |
| O15_LABOUR_LONG | Long labour | XV Pregnancy, childbirth and the puerperium (O15_) | 7866 | 162777 |
| DM_KETOACIDOSIS | Diabetic ketoacidosis | Diabetes endpoints | 7841 | 271817 |
| H7_GLAUCOMA_POAG | Primary open-angle glaucoma, strict | VII Diseases of the eye and adnexa (H7_) | 7756 | 358375 |
| H7_REFRAACCOMMODIS | Disorders of refraction and accommodation | VII Diseases of the eye and adnexa (H7_) | 7737 | 361237 |
| K11_CARIES_DENTIN | Dentinal caries | XI Diseases of the digestive system (K11_) | 7680 | 259234 |
| O15_DELIV_FORCEP_VAC | Single delivery by forceps and vacuum extractor | XV Pregnancy, childbirth and the puerperium (O15_) | 7630 | 92306 |
| K11_VENTHER | Ventral hernia | XI Diseases of the digestive system (K11_) | 7628 | 322586 |
| K11_IBD_STRICT | Inflammatory bowel disease, strict (require KELA) | XI Diseases of the digestive system (K11_) | 7625 | 369652 |
| K11_KELAIBD | IBD patients in KELA-register | XI Diseases of the digestive system (K11_) | 7625 | 359927 |
| AB1_VIRAL_OTHER_INTEST_INFECTIONS | Viral and other specified intestinal infections | I Certain infectious and parasitic diseases (AB1_) | 7613 | 336396 |
| ST19_FRACT_FOOT_ANKLE | Fracture of foot, except ankle | XIX Injury, poisoning and certain other consequences of external causes (ST19_) | 7593 | 351392 |
| N14_ENDOMETRIOSIS_ASRM_STAGE3_4 | Endometriosis ASRM stages 3,4 | XIV Diseases of the genitourinary system (N14_) | 7574 | 203296 |
| CD2_PRIMARY_LYMPHOID_HEMATOPOIETIC_EXALLC | Primary_lymphoid and hematopoietic malignant neoplasms (controls excluding all cancers) | II Neoplasms from hospital discharges (CD2_) | 7519 | 299952 |
| K11_OTHGASTR | Other gastritis (incl. Duodenitis) | XI Diseases of the digestive system (K11_) | 7514 | 320387 |
| J10_PERITONSABSC | Peritonsillar abscess | X Diseases of the respiratory system (J10_) | 7510 | 283342 |
| F5_PSYCHOTH | Other and unspecified nonorganic psychotic disorders | V Mental and behavioural disorders (F5_) | 7508 | 364160 |
| E4_DMNAS | Unspecified diabetes | IV Endocrine, nutritional and metabolic diseases (E4_) | 7455 | 308280 |
| O15_LABOUR_MALPOS | Obstructed labour due to malposition and malpresentation of fetus | XV Pregnancy, childbirth and the puerperium (O15_) | 7414 | 162777 |
| I9_AORTANEUR | Aortic aneurysm | IX Diseases of the circulatory system (I9_) | 7395 | 349539 |
| M13_CERVICOBRACHSDR | Cervicobrachial syndrome | XIII Diseases of the musculoskeletal system and connective tissue (M13_) | 7353 | 270964 |

| | | | | |
|---|---|---|---|---|
| DM_HYPOGLYC | Diabetic hypoglycemia | Diabetes endpoints | 7332 | 271817 |
| H7_VISUDISTURBSUB | Subjective visual disturbances | VII Diseases of the eye and adnexa (H7_) | 7313 | 360091 |
| H7_IRIDOCYCLITIS | Iridocyclitis | VII Diseases of the eye and adnexa (H7_) | 7306 | 357814 |
| M13_OSTEOPOROSIS | Osteoporosis | XIII Diseases of the musculoskeletal system and connective tissue (M13_) | 7300 | 358014 |
| M13_GANGLION | Ganglion | XIII Diseases of the musculoskeletal system and connective tissue (M13_) | 7270 | 275212 |
| CPAP | Continuous positive airway pressure | Interstitial lung disease endpoints | 7247 | 164315 |
| O15_PRE_OR_ECLAMPSIA | Pre-eclampsia or eclampsia | XV Pregnancy, childbirth and the puerperium (O15_) | 7212 | 194266 |
| H7_STRABOTH | Other strabismus | VII Diseases of the eye and adnexa (H7_) | 7179 | 361237 |
| H8_TINNITUS | Tinnitus | VIII Diseases of the ear and mastoid process (H8_) | 7175 | 331736 |
| O15_MEMBR_PREMAT_RUPT | Premature rupture of membranes | XV Pregnancy, childbirth and the puerperium (O15_) | 7147 | 168929 |
| NONALLERG_ASTHMA_EXMORE | Non-allergic asthma (mode) (more control exclusions) | Asthma and related endpoints | 7143 | 202399 |
| M13_ENTESOPATHYLOW | Enthesopathies of lower limb, excluding foot | XIII Diseases of the musculoskeletal system and connective tissue (M13_) | 7125 | 270964 |
| COPD_EARLY | Early onset COPD | COPD and related endpoints | 7079 | 359794 |
| I9_PHLETHROM | Phlebitis and thrombophlebitis (not including DVT) | IX Diseases of the circulatory system (I9_) | 7037 | 324121 |
| M13_MYALGIA | Myalgia | XIII Diseases of the musculoskeletal system and connective tissue (M13_) | 7024 | 275212 |
| CD2_BENIGN_COLON_NOS | Benign neoplasm: Colon, unspecified | II Neoplasms from hospital discharges (CD2_) | 7013 | 370264 |
| F5_BIPO | Bipolar affective disorders | V Mental and behavioural disorders (F5_) | 7006 | 329192 |
| N14_FEMGENPAIN | Pain and other conditions associated with female genital organs and menstrual cycle | XIV Diseases of the genitourinary system (N14_) | 6976 | 107564 |
| ST19_FRACT_SKULL_FACIAL_BONES | Fracture of skull and facial bones | XIX Injury, poisoning and certain other consequences of external causes (ST19_) | 6907 | 329318 |
| D3_BLOODOTHER | Other diseases of blood and blood-forming organs | III Diseases of the blood and blood-forming organs and certain disorders involving the immune mechanism (D3_) | 6877 | 370400 |
| N14_PHIMOSIS | Redundant prepuce, phimosis and paraphimosis | XIV Diseases of the genitourinary system (N14_) | 6859 | 119297 |
| K11_DISORD_TEETH | Other disorders of teeth and supporting structures | XI Diseases of the digestive system (K11_) | 6850 | 259234 |
| O15_PREG_MATERN_CARE | Maternal care for other conditions predominantly related to pregnancy | XV Pregnancy, childbirth and the puerperium (O15_) | 6845 | 179899 |
| FE | Focal epilepsy | VI Diseases of the nervous system (G6_) | 6842 | 365537 |
| H7_RETINOPATHYDIAB | Diabetic retinopathy | VII Diseases of the eye and adnexa (H7_) | 6818 | 344569 |
| F5_PERSMOOD | Persistent mood disorders | V Mental and behavioural disorders (F5_) | 6756 | 329192 |

| | | | | |
|---|---|---|---|---|
| APPENDACUT_COMPLIC | Acute appendicitis, with complications | XI Diseases of the digestive system (K11_) | 6747 | 346283 |
| G6_MIGRAINE_NO_AURA | Migraine without aura | VI Diseases of the nervous system (G6_) | 6730 | 287837 |
| TRAUMBRAIN_NONCONCUS | severe traumatic brain injury, does not include concussion | Neurological endpoints | 6687 | 370590 |
| M13_CERVICALGIA | Cervicalgia | XIII Diseases of the musculoskeletal system and connective tissue (M13_) | 6684 | 270964 |
| L12_CELLULITIS | Cellulitis | XII Diseases of the skin and subcutaneous tissue (L12_) | 6683 | 355969 |
| O15_PREECLAMPS | Pre-eclampsia | XV Pregnancy, childbirth and the puerperium (O15_) | 6663 | 194266 |
| F5_SUBSNOALCO | Substance use, excluding alcohol | V Mental and behavioural disorders (F5_) | 6605 | 353355 |
| M13_OTHERBONE | Other disorders of bone | XIII Diseases of the musculoskeletal system and connective tissue (M13_) | 6568 | 358014 |
| H8_EXTOTITIS | Otitis externa | VIII Diseases of the ear and mastoid process (H8_) | 6547 | 367005 |
| H7_IRIDOCYC_ANTER | Anterior Iridocyclitis | VII Diseases of the eye and adnexa (H7_) | 6536 | 370741 |
| I9_INTRACRA | Nontraumatic intracranial haemmorrhage | IX Diseases of the circulatory system (I9_) | 6530 | 342673 |
| F5_SCHZPHR | Schizophrenia | V Mental and behavioural disorders (F5_) | 6515 | 364160 |
| C3_COLORECTAL_EXALLC | Colorectal cancer (controls excluding all cancers) | II Neoplasms, from cancer register (ICD-O-3) | 6509 | 287137 |
| AD_LO_EXMORE | Alzheimer‚Äôs disease (Late onset) (more control exclusions) | Neurological endpoints | 6489 | 170429 |
| K11_PARODON_PERIAPIC_CHRONIC | Chronic periapical parodontitis | XI Diseases of the digestive system (K11_) | 6466 | 259234 |
| RX_INFERTILITY | Medical treatment for female infertility | Drug purchase endpoints | 6446 | 204424 |
| J10_RESPOTHER | Other diseases of the respiratory system | X Diseases of the respiratory system (J10_) | 6381 | 370896 |
| L12_URTICA_NAS | Other and unspecified urticaria | XII Diseases of the skin and subcutaneous tissue (L12_) | 6381 | 364583 |
| E4_PARATHYRO | Disorders of parathyroid gland | IV Endocrine, nutritional and metabolic diseases (E4_) | 6362 | 361988 |
| M13_PATELLAOTH | Other disorders of patella | XIII Diseases of the musculoskeletal system and connective tissue (M13_) | 6333 | 240862 |
| SPONDYLOARTHRITIS | Spondyloarthritis | Comorbidities of Interstitial lung disease endpoints | 6333 | 341455 |
| NIV | Non-invasive ventilation | Interstitial lung disease endpoints | 6315 | 164315 |
| ALCOHOLACUTE10 | Acute alcohol intoxication | Alcohol related diseases | 6289 | 369838 |
| K11_SALIVARY | Diseases of salivary glands | XI Diseases of the digestive system (K11_) | 6269 | 259234 |
| K11_ANOMALI_DENTAL_ARCH_RELATIONS1_INCLAVO | Anomalies of dental arch relationship, including avohilmo | XI Diseases of the digestive system (K11_) | 6260 | 371017 |
| J10_NASALPOLYP | Nasal polyp | X Diseases of the respiratory system (J10_) | 6255 | 283342 |
| E4_DM1NASCOMP | Type 1 diabetes with other specified/multiple/unspecified complications | IV Endocrine, nutritional and metabolic diseases (E4_) | 6234 | 308280 |
| K11_ACUTPANC | Acute pancreatitis | XI Diseases of the digestive system (K11_) | 6223 | 330903 |

| | | | | |
|---|---|---|---|---|
| FE_STRICT | Focal epilepsy, strict definition | VI Diseases of the nervous system (G6_) | 6213 | 365534 |
| ST19_FRACT_LUMBAR_SPINE_PELVIS | Fracture of lumbar spine and pelvis | XIX Injury, poisoning and certain other consequences of external causes (ST19_) | 6209 | 364504 |
| M13_ARTHROSIS_POLY | Polyarthrosis | XIII Diseases of the musculoskeletal system and connective tissue (M13_) | 6185 | 240862 |
| H7_IRIDOACUTE | Acute and subacute iridocyclitis | VII Diseases of the eye and adnexa (H7_) | 6166 | 357814 |
| F5_BEHEMOCHILD | Behavioural and emotional disorders with onset usually occurring in childhood and adolescence | V Mental and behavioural disorders (F5_) | 6160 | 371117 |
| RHEUMA_OTHER_WIDE | Other (seronegative) rheumatoid arthritis, wide | Rheuma endpoints | 6153 | 371124 |
| E4_GOITREMULTINOD | Nontoxic multinodular goitre | IV Endocrine, nutritional and metabolic diseases (E4_) | 6149 | 320703 |
| POLLENALLERG | Pollen allergy | Comorbidities of Asthma | 6118 | 368409 |
| N14_HYPERTROPHYBREAST_BOTH | Hypertrophy of breast in women and men | XIV Diseases of the genitourinary system (N14_) | 6105 | 359487 |
| DRY_AMD | Dry age-related macular degeneration (includes geographic atrophy) | VII Diseases of the eye and adnexa (H7_) | 6065 | 251042 |
| D3_IMMUNEMECHANISM | Certain disorders involving the immune mechanism | III Diseases of the blood and blood-forming organs and certain disorders involving the immune mechanism (D3_) | 6022 | 371255 |
| N14_FEMALE_GENITAL_HSIL | HSIL lesion of the cervix uteri, vagina or vulva | XIV Diseases of the genitourinary system (N14_) | 5996 | 204874 |
| C3_BREAST_ERNEG_EXALLC | Malignant neoplasm of breast, HER-negative (controls excluding all cancers) | II Neoplasms, from cancer register (ICD-O-3) | 5965 | 167017 |
| O15_MAT_CARE_MALPRESENT | Maternal care for known or suspected malpresentation of fetus | XV Pregnancy, childbirth and the puerperium (O15_) | 5957 | 168929 |
| K11_GULC | Gastric ulcer | XI Diseases of the digestive system (K11_) | 5935 | 320387 |
| K11_OTHILEUS | Other or unspecified ileus, impaction or obstruction | XI Diseases of the digestive system (K11_) | 5925 | 301931 |
| N14_ACUTERENFAIL | Acute renal failure | XIV Diseases of the genitourinary system (N14_) | 5908 | 363177 |
| I9_CARDMYO | Cardiomyopathy | IX Diseases of the circulatory system (I9_) | 5874 | 286109 |
| N14_ENDOMETRIOSIS_OVARY | Endometriosis of ovary | XIV Diseases of the genitourinary system (N14_) | 5867 | 107564 |
| ASTHMA_CHILD_EXMORE | Childhood asthma (age<16) (more control exclusions) | Asthma and related endpoints | 5865 | 202399 |
| AB1_VIRAL_NOS | Other viral diseases, not elsewhere classified | I Certain infectious and parasitic diseases (AB1_) | 5864 | 367472 |
| C3_BRONCHUS_LUNG_EXALLC | Malignant neoplasm of bronchus and lung (controls excluding all cancers) | II Neoplasms, from cancer register (ICD-O-3) | 5842 | 287137 |
| L12_SEBORRKERAT | Seborrhoeic keratosis | XII Diseases of the skin and subcutaneous tissue (L12_) | 5835 | 353088 |
| VWXY20_INTENTI_SELF_P_EXPOS_OTHER_UNSPE_CHEMIC_NOXIO_SUBST | Intentional self-poisoning by and exposure to other and unspecified chemicals and noxious substances | XX External causes of morbidity and mortality (VWXY20_) | 5811 | 371466 |

| | | | | |
|---|---|---|---|---|
| K11_ORAL_LICHEN_PLANUS_WIDE | Oral lichen ruber planus, wide definition | XI Diseases of the digestive system (K11_) | 5791 | 371486 |
| ABDOM_HERNIA_POSTOP | Hernia of abdominal wall, postoperative | XI Diseases of the digestive system (K11_) | 5775 | 210314 |
| D3_COAGDEF_PURPUR_HAEMORRHAGIC | Coagulation defects, purpura and other haemorrhagic conditions | III Diseases of the blood and blood-forming organs and certain disorders involving the immune mechanism (D3_) | 5773 | 371504 |
| N14_ENDOMETRIOSIS_ASRM_STAGE1_2 | Endometriosis ASRM stages 1,2 | XIV Diseases of the genitourinary system (N14_) | 5769 | 205101 |
| N14_SALPHOOPH | Salpingitis and oophoritis | XIV Diseases of the genitourinary system (N14_) | 5768 | 188295 |
| M13_SPONDYLOLISTHESIS | Spondylolisthesis/Spondylolysis | XIII Diseases of the musculoskeletal system and connective tissue (M13_) | 5766 | 270964 |
| L12_PSORI_VULG | Psoriasis vulgaris | XII Diseases of the skin and subcutaneous tissue (L12_) | 5759 | 364071 |
| AB1_SEXUAL_TRANSMISSION_NOS | Other predominantly sexually transmitted diseases, not elsewhere classified | I Certain infectious and parasitic diseases (AB1_) | 5753 | 366340 |
| TEMPOROMANDIB | Temporomandibular joint disorders | XI Diseases of the digestive system (K11_) | 5668 | 205355 |
| O15_PREG_ECTOP | Ectopic pregnancy | XV Pregnancy, childbirth and the puerperium (O15_) | 5648 | 149622 |
| N14_GLOMERULAR | Glomerular diseases | XIV Diseases of the genitourinary system (N14_) | 5642 | 371635 |
| AB1_BACT_INTEST_OTH | Other bacterial intestinal infections | I Certain infectious and parasitic diseases (AB1_) | 5631 | 336396 |
| N14_ENDOMETRIOSIS_PELVICPERITONEUM | Endometriosis of pelvic peritoneum | XIV Diseases of the genitourinary system (N14_) | 5628 | 107564 |
| M13_HAMMERTOE | Other hammer toe(s) (acquired) | XIII Diseases of the musculoskeletal system and connective tissue (M13_) | 5625 | 240862 |
| CD2_BENIGN_RECTUM_ANAL | Benign neoplasm: Rectum, anus and anal canal | II Neoplasms from hospital discharges (CD2_) | 5613 | 371664 |
| E4_HYPERPARA | Hyperparathyroidism | IV Endocrine, nutritional and metabolic diseases (E4_) | 5590 | 361988 |
| G6_POLYOTHUNS | Other and unspecified polyneuropathies, also in other diseases | VI Diseases of the nervous system (G6_) | 5553 | 370790 |
| M13_RADICULOPATHY | Radiculopathy | XIII Diseases of the musculoskeletal system and connective tissue (M13_) | 5550 | 270964 |
| M13_ADHCAPSULITIS | Adhesive capsulitis of shoulder | XIII Diseases of the musculoskeletal system and connective tissue (M13_) | 5538 | 275212 |
| I9_AVBLOCK | AV-block | IX Diseases of the circulatory system (I9_) | 5536 | 286109 |
| K11_EROSION_INCLAVO | Erosion of teeth, including avohilmo | XI Diseases of the digestive system (K11_) | 5521 | 371756 |
| H7_CORNULCER | Corneal ulcer | VII Diseases of the eye and adnexa (H7_) | 5476 | 357814 |
| L12_FOLLICULARCYST | Follicular cysts of skin and subcutaneous tissue | XII Diseases of the skin and subcutaneous tissue (L12_) | 5459 | 361140 |
| N14_CERVICAL_HSIL | HSIL lesion of the cervix uteri | XIV Diseases of the genitourinary system (N14_) | 5459 | 205411 |
| K11_IMPACTED_TEET | Impacted teeth , including avohilmo | XI Diseases of the digestive system (K11_) | 5435 | 371842 |

| | | | | |
|---|---|---|---|---|
| H_INCLAVO | | | | |
| F5_ALZHDEMENT | Dementia in Alzheimer disease | V Mental and behavioural disorders (F5_) | 5422 | 356660 |
| C3_COLORECTAL_ADENO_EXALLC | Colorectal adenocarcinoma (controls excluding all cancers) | II Neoplasms, from cancer register (ICD-O-3) | 5378 | 287137 |
| I9_SAHANEUR | Aneurysms, operations, SAH | IX Diseases of the circulatory system (I9_) | 5342 | 342673 |
| CHILDHOOD_ALLERGY | Childhood allergy (age < 16) | Asthma and related endpoints | 5337 | 371940 |
| CD2_BENIGN_OVARY | Benign neoplasm of ovary | II Neoplasms from hospital discharges (CD2_) | 5308 | 205562 |
| G6_ROOTPLEXOTHUNS | Other and unspecified nerve root and plexus disorders, also in other diseases | VI Diseases of the nervous system (G6_) | 5294 | 330377 |
| GOUT_NOS | Gout, unspecified | Rheuma endpoints | 5292 | 368788 |
| KRA_PSY_SLEEP_NONORG_EXMORE | Nonorganic sleeping disorders (more control exclusions) | Psychiatric endpoints from Katri R√§ikk√∂nen | 5254 | 277526 |
| N14_HYDROCELE | Hydrocele | XIV Diseases of the genitourinary system (N14_) | 5221 | 119297 |
| N14_HYPERTROPHYBREAST | Hypertrophy of breast | XIV Diseases of the genitourinary system (N14_) | 5212 | 193973 |
| E4_DM1OPTH | Type 1 diabetes with ophthalmic complications | IV Endocrine, nutritional and metabolic diseases (E4_) | 5202 | 308280 |
| F5_SOMATOFORM | Somatoform disorder | V Mental and behavioural disorders (F5_) | 5164 | 337577 |
| E4_HYPOOSMNAT | Hypo-osmolality and hyponatraemia | IV Endocrine, nutritional and metabolic diseases (E4_) | 5137 | 324150 |
| INFLUENZA | All influenza (not pneumonia) | Asthma and related endpoints | 5136 | 314673 |
| M13_DUPUTRYEN | Palmar fascial fibromatosis [Dupuytren] | XIII Diseases of the musculoskeletal system and connective tissue (M13_) | 5128 | 275212 |
| N14_OTHFEMPELINF | Other female pelvic inflammatory diseases | XIV Diseases of the genitourinary system (N14_) | 5117 | 188295 |
| VOCALCORDDYS | Vocal cord dysfunction | COPD and related endpoints | 5087 | 139795 |
| CD2_BENIGN_OTHERDIGESTIVE | Benign neoplasm of other and ill-defined parts of digestive system | II Neoplasms from hospital discharges (CD2_) | 5062 | 372215 |
| K11_DISLIVOTH | Other diseases of liver | XI Diseases of the digestive system (K11_) | 5052 | 366450 |
| K11_UC_STRICT2 | Ulcerative colitis (strict definition, require KELA, min 2 HDR) | XI Diseases of the digestive system (K11_) | 5034 | 371530 |
| L12_LICHENPLANUS_INCLAVO | Lichen planus, including avohilmo | XII Diseases of the skin and subcutaneous tissue (L12_) | 5015 | 372262 |

#### Supplementary Table 8: Baseline characteristics of the TriNetX database

| | After propensity-score matching | | | | | |
|---|---|---|---|---|---|---|
| | **Hypersomnia** | **Controls** | **Standardized mean difference** | **Insomnia** | **Controls** | **SMD** |
| **Total, n** | 16,371 | 16,371 | | 110,374 | 110,374 | |
| **Age, Mean ± SD** | 41.5 ± 18.9 | 42 ± 19.5 | 0.0226 | 45.6 ± 19.1 | 45.9 ± 19.7 | 0.0135 |
| **Sex (%)** | | | | | | |
| Male | 50.2 | 50.5 | 0.007 | 37.9 | 39.1 | 0.024 |
| Female | 48.3 | 48.7 | 0.007 | 59.3 | 59.3 | 0.0004 |
| **Ethnicity (%)** | | | | | | |
| Hispanic/Latino | 6.0 | 5.9 | 0.008 | 6.9 | 6.5 | 0.015 |
| Not Hispanic/Latino | 71.9 | 72.3 | 0.01 | 70.1 | 70.7 | 0.009 |
| Unknown | 22.1 | 21.8 | 0.006 | 22.5 | 22.8 | 0.007 |
| **Race (%)** | | | | | | |
| White | 71.5 | 71.4 | 0.002 | 74.3 | 74.7 | 0.014 |
| Black | 12.5 | 12.6 | 0.003 | 9.8 | 9.5 | 0.01 |
| Asian | 2.5 | 2.5 | 0.004 | 2.7 | 2.7 | 0.002 |
| **Other risk/conditions (%)** | | | | | | |
| Cerebrovascular diseases | 0.9 | 0.9 | 0.002 | 0.6 | 0.8 | 0.017 |
| Neoplasms | 4.1 | 4.1 | 0.0003 | 6.1 | 6.5 | 0.018 |
| Certain infectious and parasitic diseases | 3.5 | 3.4 | 0.002 | 4.7 | 4.4 | 0.016 |
| **Pre-existing medical procedures (%)** | | | | | | |
| Office or other outpatient services | 25.3 | 25.3 | 0.002 | 23.2 | 24.8 | 0.037 |
| Hospital inpatient and observation care services | 0.9 | 1.1 | 0.02 | 1.4 | 1.5 | 0.006 |

| | | | | | | |
|---|---|---|---|---|---|---|
| **Medication prescriptions (%)** | | | | | | |
| Beta-blockers | 3.3 | 3.3 | <0.0001 | 4.1 | 4.6 | 0.029 |
| ACE inhibitors | 1.9 | 2.1 | 0.014 | 2.5 | 2.7 | 0.014 |
| Angiotensin II inhibitor | 1.9 | 2.1 | 0.014 | 1.4 | 1.3 | 0.013 |
| Calcium channel blockers | 1.6 | 1.6 | <0.0001 | 2.0 | 2.0 | 0.001 |
| Metformin | 1.1 | 0.9 | 0.02 | 1.1 | 1.0 | 0.009 |
| Glucagon-like Peptide 1 Agonist | 0.1 | 0.1 | 0.019 | 0.1 | 0.0 | 0.02 |
| **Laboratory values** | | | | | | |
| Hemoglobin A1C/Hemoglobin total in blood [%] | 6.07 ± 1.55 | 6.95 ± 1.61 | 0.557 | 6.11 ± 1.66 | 6.8 ± 1.63 | 0.420 |
| Cholesterol in serum or plasma [mg/dL] | 182 ± 46.4 | 178 ± 43.4 | 0.094 | 182 ± 48.7 | 178 ± 45.5 | 0.081 |
| Cholesterol in LDL in serum or plasma [mg/dL] | 109 ± 34.9 | 105 ± 34.4 | 0.109 | 106 ± 35.5 | 105 ± 33.2 | 0.047 |
| Cholesterol in HDL in serum or plasma [mg/dL] | 47.7 ± 17.6 | 49.6 ± 15.8 | 0.115 | 53.4 ± 19.9 | 51.5 ± 16.7 | 0.104 |
| Body Mass Index | 30.2 ± 8.06 | 26.8 ± 6.28 | 0.469 | 27.2 ± 6.59 | 27 ± 6.4 | 0.03 |
